# The Th17/IL-17 Axis and Systemic Inflammatory Biomarkers in Sepsis-Associated Encephalopathy and Delirium: A Systematic Review and Meta-Analysis

**DOI:** 10.64898/2026.10.16.26365073

**Authors:** Adriano Chaves-Filho, Edna Fernandes do Nascimento, Luisa Domingos Cancela Goncalves, Francisco Júlio Ribeiro Dias Filho, Tatiana de Queiroz Oliveira, Danielle S. Macedo, Felipe Dal-Pizzol, Maria Elisabete Amaral de Moraes, Tatiana Barichello

## Abstract

**OBJECTIVE:** Sepsis-associated encephalopathy (SAE) and sepsis-associated delirium (SAD) lack confirmatory laboratory tests or biomarkers. We evaluated whether the Th17/interleukin (IL)-17 axis or neuroglial-injury biomarkers reflect acute brain dysfunction beyond systemic inflammation and whether biomarker profiles differ between these phenotypes.

**DATA SOURCES:** EMBASE, MEDLINE/PubMed, and the Cochrane Library from inception to August 1, 2026, supplemented by backward and forward citation searching, following PRISMA 2020.

**STUDY SELECTION:** Observational studies of adults with sepsis classified by SAE or SAD status and reporting circulating biomarker concentrations by neurological group or biomarker-outcome associations. Eighteen studies (2,388 participants; 1,863 with sepsis) were included.

**DATA EXTRACTION:** Two reviewers independently extracted study, phenotype, assay, and outcome data. Risk of bias was assessed with the Newcastle-Ottawa Scale and certainty with GRADE.

**DATA SYNTHESIS:** Random-effects models pooled standardized mean differences (SMDs) separately against non-septic and septic controls and odds ratios (ORs) for biomarker-outcome associations. IL-6 was the only mediator elevated in both comparative frameworks and associated with outcomes (non-septic controls: SMD, 0.54 [0.25–0.82]; septic controls: SMD, 1.05 [0.51–1.59]; OR, 1.85 [1.50–2.29]). S100B, neuron-specific enolase, IL-10, tumor necrosis factor-α, and interferon-γ were elevated versus septic controls, whereas IL-17 was modestly elevated only versus non-septic controls (SMD, 0.37 [0.01–0.73]). The IL-6 septic-control effect was concentrated in SAE (p<0.0001) and remained robust in leave-one-out and risk-of-bias sensitivity analyses. Certainty was low to very low, and all 14 computable prediction intervals included the null.

**CONCLUSIONS:** IL-6 emerges as the leading circulating candidate biomarker of sepsis-associated brain dysfunction, while S100B and NSE capture neuroglial injury beyond sepsis. Current evidence does not establish a discriminating role for the Th17 axis. Future studies require standardized neurological phenotyping and appropriately matched septic controls to establish diagnostic value.

**KEY POINTS:** *Question:* Do the Th17/IL-17 axis or alternative inflammatory and neuroglial-injury biomarkers identify sepsis-associated encephalopathy or delirium beyond the inflammatory signal of sepsis itself?

*Findings:* In this systematic review and meta-analysis of 18 observational studies and 2388 participants, IL-6 was the only biomarker significantly elevated both against non-septic and against septic controls and significantly associated with the sepsis-associated neurological outcome; IL-17 was modestly elevated only against non-septic controls, and every computable prediction interval included the null.

*Meaning:* IL-6, with the neuroglial-injury markers S100B and NSE, is the most promising basis for multimarker models; standardized phenotyping and septic-controlled designs are the next step.

**Summary:** In 18 studies (2388 patients), IL-6 alone was raised against non-septic and septic controls and linked to sepsis-associated encephalopathy and delirium. IL-17 rose only versus non-septic controls and could not be pooled against septic ones. No marker works stand-alone.

## INTRODUCTION

Acute brain dysfunction is among the most frequent organ failures in sepsis, a syndrome implicated in roughly one in five deaths worldwide [1, 2]. Depending on the ascertainment instrument and the population, SAE or SAD is documented in a substantial minority to a majority of septic patients in intensive care [3–5] and is independently associated with prolonged ventilation, higher mortality and persistent cognitive impairment [6–8]. SAE is an acute, diffuse cerebral dysfunction arising in sepsis without direct central nervous system infection or another primary neurological cause; its spectrum ranges from delirium to coma, while SAD represents the delirium phenotype within it [9]. Unlike the other organ failures scored in sepsis [10], SAE has no confirmatory laboratory test and is diagnosed by exclusion with bedside instruments designed for delirium screening [3, 11, 12]; multimodal assessment combining clinical examination, electroencephalography, neuroimaging and brain-injury biomarkers can characterize the syndrome but cannot confirm it in isolation [13].

That gap has motivated two decades of biomarker studies [14, 15], whose candidates should behave differently by class: innate and acute-phase mediators index the inflammatory response common to all sepsis [16, 17] and should separate septic from non-septic patients more sharply than encephalopathic from non-encephalopathic ones, whereas neuroglial-injury markers index structural and barrier damage documented in the septic brain [18–21] and should behave oppositely. Between them sit the T-helper-lineage cytokines, whose relevance is mechanistic rather than merely correlative [22]. IL-17A, produced by Th17 cells under IL-6, IL-1β and IL-23 [23], can disrupt the blood-brain barrier (BBB) integrity by downregulating endothelial tight-junctions [24, 25], and its blockade attenuates BBB leakage and behavioral deficits in murine models of sepsis [26, 27]. If that pathway operates in human sepsis [28], the axis should track the neurological phenotype more closely than generic inflammatory activation markers does.

Existing syntheses have not tested this: they address single markers [29–31] in a single contrast framework and generally pool SAE and SAD without distinguishing them [32–34]. A marker that separates SAE from healthy people may simply be marking sepsis, and if SAE and SAD are distinct phenotypes, pooling them attenuates real effects. We therefore synthesized the full mediator panel, estimating biomarker differences separately in each contrast framework, the association between biomarker level and the neurological outcome, and one explicit priority ordering across the panel, with the Th17 axis prespecified as the hypothesis of interest against Th1, innate, regulatory and neuroglial comparators.

## MATERIALS AND METHODS

The review followed a protocol registered in PROSPERO before extraction (CRD420261371685) and is reported according to PRISMA 2020 [35]; the completed checklist is eAppendix 1 and deviations from the protocol are in eTable 20. We included observational studies of adults with sepsis, septic shock or documented systemic infection meeting the study’s own definition [2], classified by the presence or absence of SAE or SAD by any explicit instrument, and reporting at least one circulating or cerebrospinal fluid biomarker concentration by neurological group or an effect estimate for the biomarker-outcome association; abstracts without a full report, reviews, preclinical studies, studies without a comparison group and studies reporting only composite inflammatory scores were excluded, without language or date restriction. EMBASE, MEDLINE/PubMed and the Cochrane Library were searched from inception to 1 August 2026, with backward and forward citation searching; full strings and per-source yields are in eTable 1 [36]. Records were screened in duplicate on title, abstract and full text, and data were extracted into a piloted form; medians were converted to means by established methods [37–39], standard errors to standard deviations, and values reported only graphically were digitized, and case definitions were recovered post hoc from the source of record on verbatim match (eTable 8). Risk of bias was rated in duplicate with the Newcastle-Ottawa Scale [40], used in preference to QUADAS-2 because no study reported a prespecified threshold, index-test protocol or blinded reference standard [41] (eTable 2).

Two contrast frameworks were prespecified because they answer different questions and are not interchangeable: a non-septic-control framework (cases vs. healthy controls, non-septic hospitalized patients or hospitalized patients without delirium, some of them septic), which cannot separate the contribution of sepsis from that of encephalopathy, and a septic-control framework (cases vs. septic patients without encephalopathy), which is the clinically discriminating contrast and was designated primary. A third association framework pools ORs for the relationship between biomarker level and the neurological outcome.

Continuous comparisons used Hedges’ g, a positive SMD denoting a higher concentration in the SAE or SAD group, and associations were pooled on the log-odds scale, both with random-effects models (DerSimonian-Laird) [42]; frameworks are reported side by side but never pooled. Heterogeneity was quantified with I², τ² and Cochran’s Q [43], and for every comparison with three or more studies we report a 95% prediction interval [44, 45] computed from study-level effect sizes assembled from the arm-level values printed in the forest panels (eTable 9). Each pooled SMD was additionally expressed as the implied area under the receiver-operating-characteristic curve, AUC = Φ(SMD/√2) [46], reported as implied and never as measured accuracy. Prespecified subgroup analyses examined phenotype (SAE vs. SAD) with the Q-test for interaction and are hypothesis-generating [47]; prespecified sensitivity analyses excluded studies at high risk of bias and cerebrospinal fluid measurements and omitted each study in turn. Certainty was rated with GRADE in its prognostic-factor form [48, 49], with downgrading rules prestated and applied mechanically, including risk of bias rated per comparison by weighting each study’s Newcastle-Ottawa band by its random-effects weight in that comparison (eTable 10). We prespecified a biomarker prioritization index (BPI) combining evidence quantity, consistency, effect magnitude and outcome association, rescaled within each framework, with equal base weights and 5 weighting scenarios. Analyses were performed in Review Manager and, for small-study effects, in R (metafor). Full methods are in eAppendix 2.

## RESULTS

### Included Studies

The searches returned 2831 records (727 duplicates removed), of which 2104 were screened, 26 were assessed in full text and 18 studies were included [50–68] (Fig. 1); citation searching added none. They comprised 2388 participants, 1863 with sepsis and 1415 in SAE or SAD arms (Table 1), predominantly from prospective cohorts; one study measured cerebrospinal fluid [64]. Sepsis definitions, ascertainment instruments, from Confusion Assessment Method for the Intensive Care Unit (CAM-ICU) screening to unstructured clinical diagnosis, sampling times and assays differed. Seventeen studies contributed to at least one pooled comparison and one (Kuswardhani et al. 2017 [55]) to none; a case definition could be established for 9 of 18 studies, none electroencephalographic. Of 17 assessed studies, 13 were at low risk of bias, 2 at moderate and 2 at high, with losses concentrated in comparability (eTable 2).

**Figure 1.**
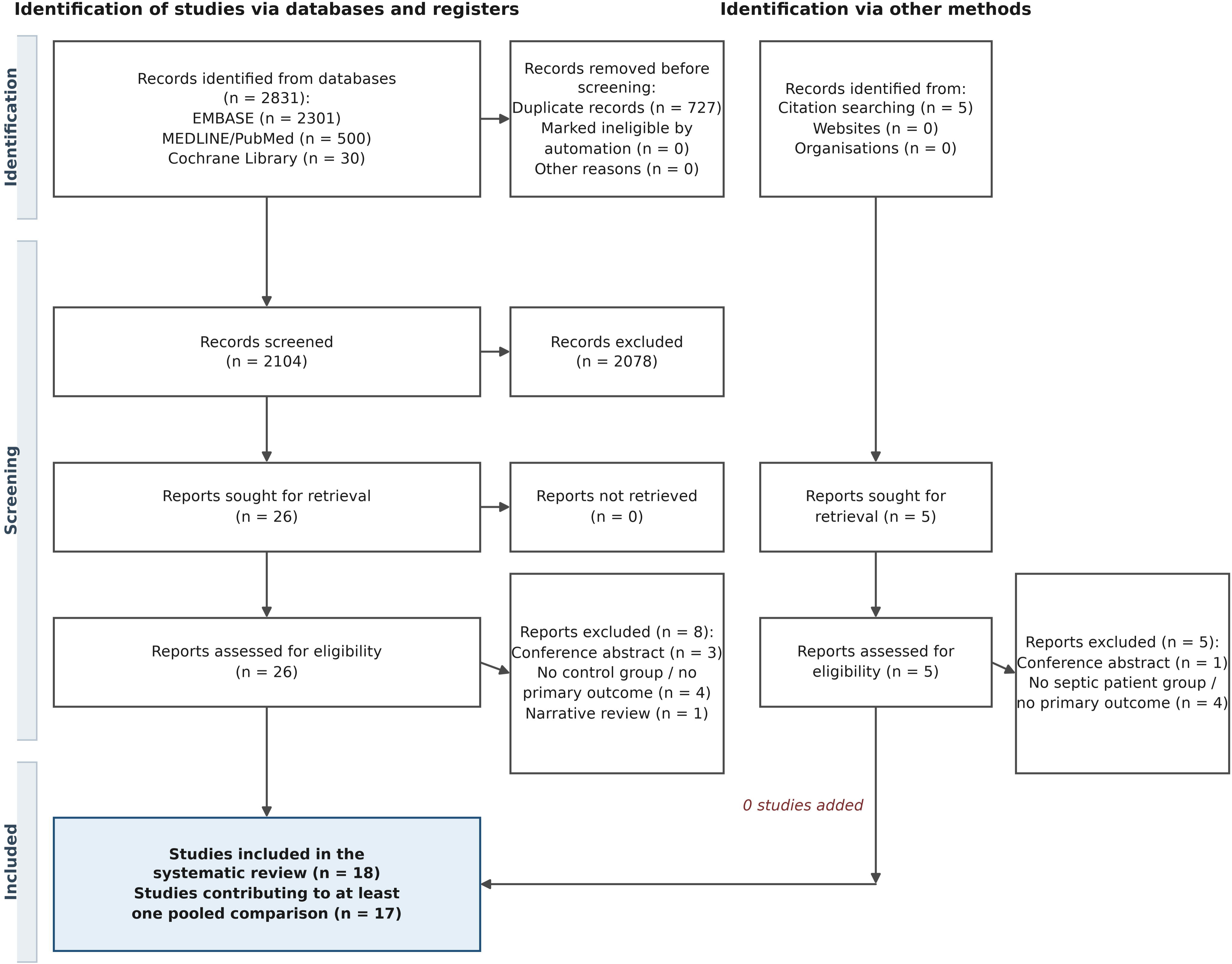
PRISMA 2020 flow diagram. Identification, screening, eligibility assessment and inclusion, shown separately for database searching and for other methods. The citation-searching arm identified 5 reports, all excluded at full text. Of the 18 included studies, 17 contributed to at least one pooled comparison.

**Table 1.** Characteristics of the 18 included studies.

### Biomarkers Versus Non-Septic Controls

Nine biomarkers were pooled against non-septic controls (Fig. 2A, Table 2); C-reactive protein (CRP) was reported against non-septic controls by a single study and was not pooled. Four intervals excluded the null, all of small to moderate magnitude (implied AUCs 0.57 to 0.65): IL-6 (SMD 0.54, 95% CI 0.25 to 0.82; p <0.001; I² = 72%; k = 7; n = 1472), IL-8 (SMD 0.47, 95% CI 0.12 to 0.82; p = 0.009; I² = 75%; k = 4; n = 1268), IL-1β (SMD 0.24, 95% CI 0.03 to 0.45; p = 0.02; I² = 43%; k = 5; n = 1346) and IL-17 (SMD 0.37, 95% CI 0.01 to 0.73; p = 0.047; I² = 48%; k = 3; n = 277). IFN-γ, IL-10, TNF-α, IL-12 and soluble TNF receptor 1 were not elevated.

**Figure 2.**
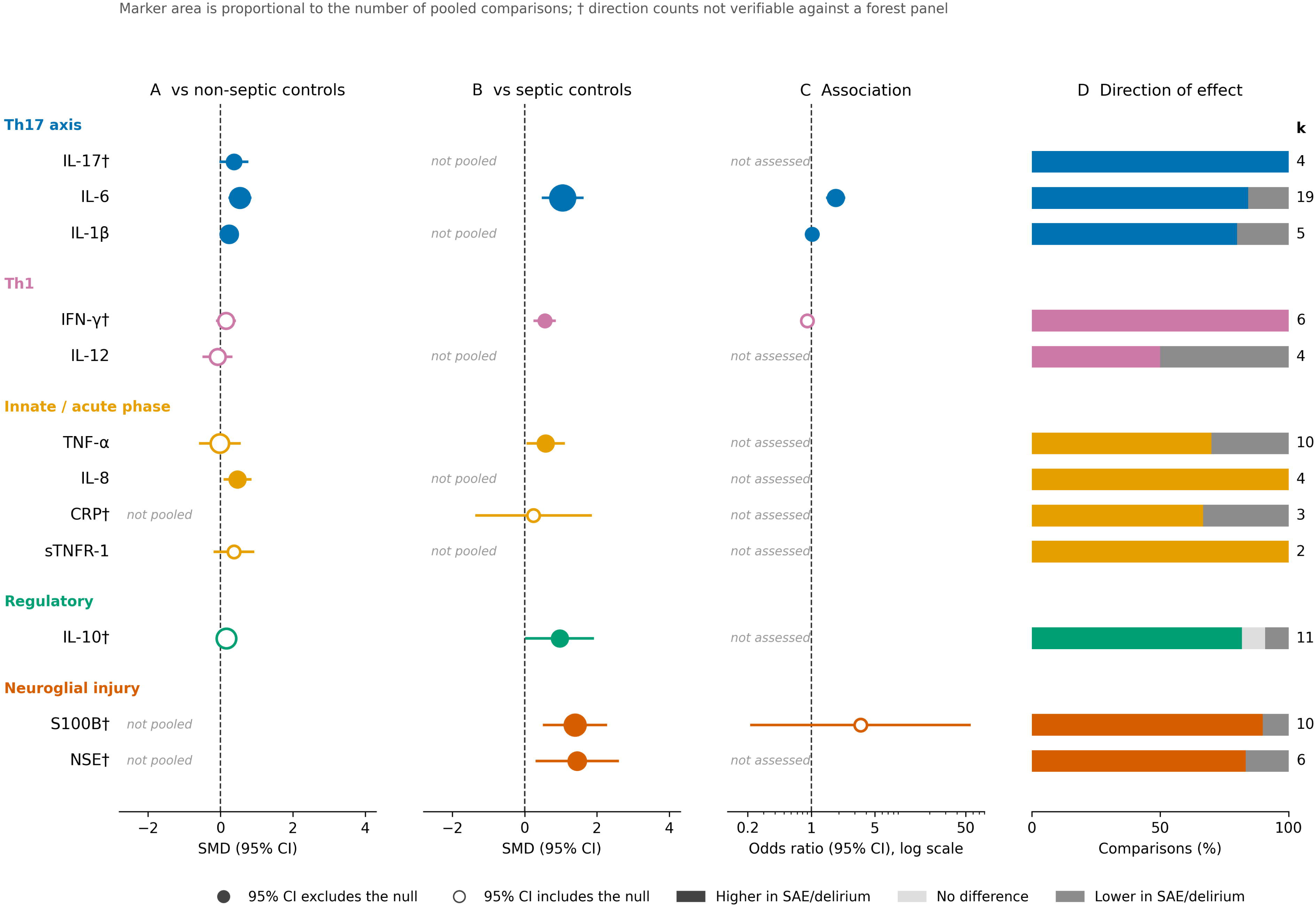
Integrated evidence landscape across the 12 biomarkers, grouped by immunological domain on a common vertical axis. **A**, Pooled standardized mean difference versus non-septic controls. **B**, Versus septic controls without encephalopathy, on the same scale as **A**. **C**, Pooled odds ratio for association with sepsis-associated encephalopathy or delirium, logarithmic scale. **D**, Direction-of-effect vote counts as a proportion of all comparisons, with the number of comparisons annotated at right. Filled markers denote a 95% CI excluding the null, open markers an interval including it; marker area is proportional to the number of pooled comparisons. Rows labeled “not pooled” or “not assessed” indicate that the comparison does not exist in the literature, not that it was null. A dagger marks biomarkers whose direction counts could not be verified against a forest panel. Per-study forest plots are in eFigures 1-3.

**Table 2.** Evidence map: volume of evidence and pooled estimates by biomarker, in both contrast frameworks and in the association framework.

### Biomarkers Versus Septic Controls

Seven biomarkers were pooled in the discriminating framework (Fig. 2B, Table 2). Six were elevated: IL-6 (SMD 1.05, 95% CI 0.51 to 1.59; p <0.001; I² = 92%; k = 12; n = 921), NSE (SMD 1.45, 95% CI 0.33 to 2.57), S100B (SMD 1.39, 95% CI 0.54 to 2.25), IL-10 (SMD 0.97, 95% CI 0.05 to 1.88), TNF-α (SMD 0.58, 95% CI 0.08 to 1.08) and IFN-γ (SMD 0.55, 95% CI 0.28 to 0.83; I² = 0%; k = 2; n = 222); CRP did not discriminate (SMD 0.25, 95% CI -1.33 to 1.82; p = 0.76; I² = 78%; k = 2; n = 38). Conversely, TNF-α and IFN-γ are elevated against septic but not against non-septic controls, which is more consistent with comparator composition than with a biological gradient.

Prediction intervals were computed for 14 of 20 comparisons; all 14 include the null, from -0.35 to 0.83 (IL-1β vs. non-septic controls) to -3.31 to 5.24 (IL-10 vs. septic controls), and -1.03 to 3.14 for IL-6 versus septic controls (eTable 9). Expressed as implied discrimination, the septic-control differences correspond to AUCs of 0.57 to 0.85 (Fig. 3B, eTable 23): none reaches the 0.90 expected of a stand-alone test, every lower limit falls below 0.70, and for IL-6 the range a new cohort should expect (0.23 to 0.99) includes values worse than chance; against non-septic controls no value exceeded 0.65 (Fig. 3A).

**Figure 3.**
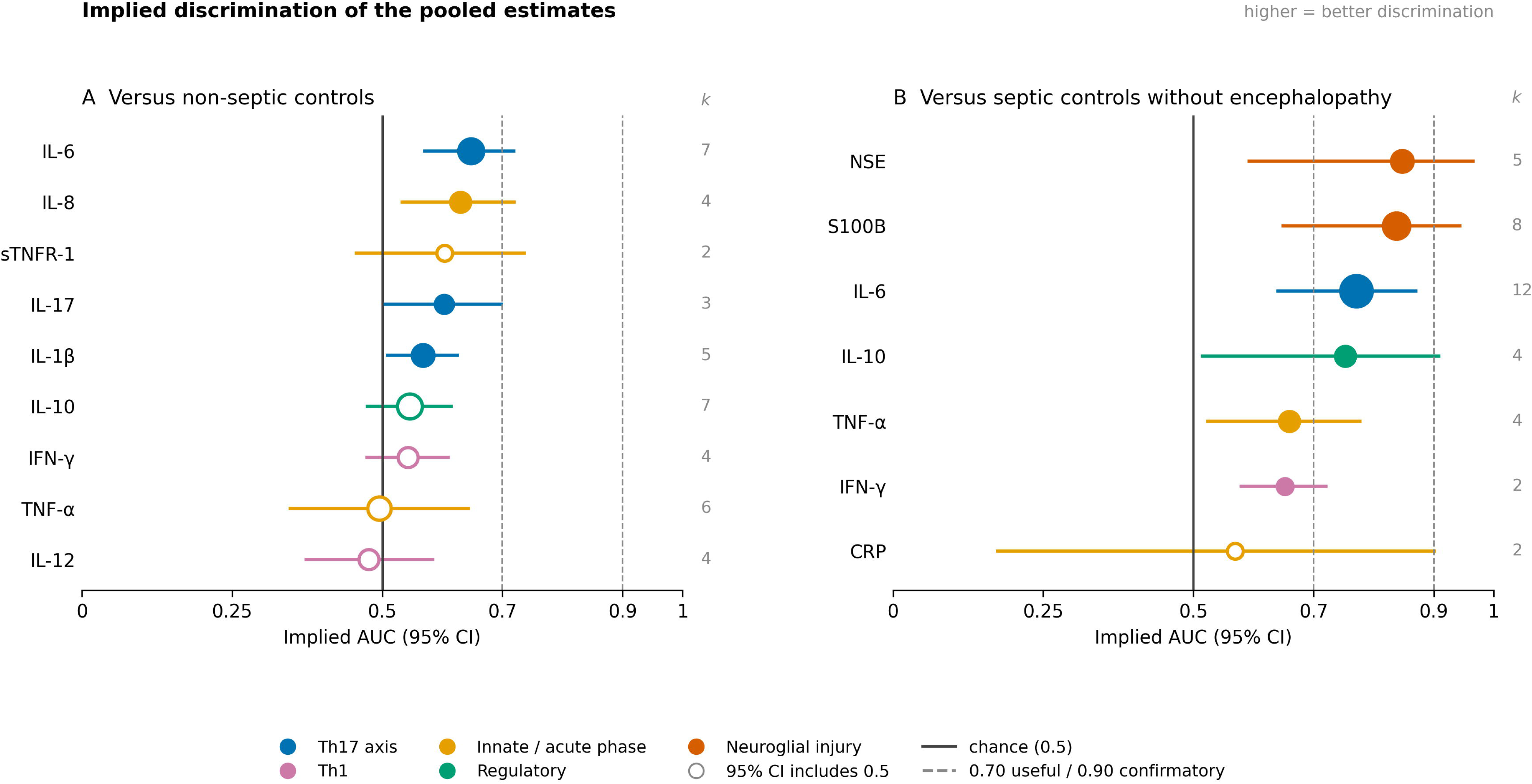
Implied discrimination of every pooled comparison. Each pooled standardized mean difference is shown on the scale of the implied area under the receiver-operating-characteristic curve, AUC = Φ(SMD/√2), applied to the point estimate and both confidence limits. **A**, Versus non-septic controls. **B**, Versus septic controls without encephalopathy. Markers are filled where the interval excludes 0.5; marker area and the printed k are proportional to the number of contributing comparisons; color denotes immunological domain, as in Figures 2 and 4. The solid line marks chance and the dashed lines the conventional thresholds for useful (0.70) and confirmatory (0.90) discrimination. These are implied, not measured, accuracies: no primary study contributed a receiver-operating-characteristic analysis, and each value inherits the heterogeneity of the pool it is transformed from.

### Association With the Neurological Outcome and Direction of Effect

Four biomarkers had pooled ORs for association with the neurological outcome (SAE or delirium) (Fig. 2C, eFigure 3). IL-6 showed OR 1.85, 95% CI 1.50 to 2.29; p <0.001; I² = 34%; k = 4, the only association estimate that is both statistically clear and clinically interpretable. IL-1β showed OR 1.02, 95% CI 1.01 to 1.03; p = 0.002; I² = 0%; k = 2, excluding the null at a magnitude with no plausible bedside value; S100B and IFN-γ were null. Vote counting on direction of effect (Fig. 2D, eTable 21) shows a directionally consistent literature: for 9 of 12 biomarkers at least three quarters of comparisons pointed to higher concentrations in the encephalopathic group.

### Phenotype Subgroups, Certainty and Prioritization

Eight subgroup analyses stratified comparisons into SAE and SAD strata (eTable 5). Four showed statistically clear differences, all with the effect concentrated in the SAE stratum: IL-6 versus septic controls (SAE SMD 1.38, 95% CI 0.83 to 1.94 vs. SAD SMD -0.02, 95% CI -0.37 to 0.33; p <0.0001), S100B (p = 0.02), NSE (p = 0.02) and TNF-α (p = 0.008; SAE stratum a single study). For IL-17 against non-septic controls, the SAE stratum was null (SMD 0.20, 95% CI -0.14 to 0.53; k = 2) and the positive estimate rested on a single SAD comparison (p = 0.07). Egger’s test, possible only for IL-6 versus septic controls, was not significant (p = 0.11; eTable 6). Excluding the two studies at high risk of bias, one of which provided the only cerebrospinal fluid measurements, changed the significance of no pooled estimate (eTable 26). On leave-one-out analysis (eTable 25), IL-6 in both frameworks, S100B and NSE remained significant after omission of any single study, whereas IL-1β and IL-17 against non-septic controls and IL-10 and TNF-α against septic controls each lost significance when one study was omitted. Under the mechanical GRADE rules, 8 of 9 comparisons against non-septic controls and 6 of 7 against septic controls were very low certainty (eTables 3 and 4), the exceptions being IL-1β against non-septic and IFN-γ against septic controls (low). Indirectness was downgraded for every comparison and inconsistency for most, whereas risk of bias downgraded 7 of 9 comparisons against non-septic controls and none against septic controls (eTable 10). On the prioritization index, the highest-ranked biomarkers were IL-6 (88.9), S100B (62.5) and NSE (55.4) against septic controls, with identical ranks in all 5 scenarios (Fig. 4A, eTable 24), and IL-6 (96.4), IL-1β (74.0) and IL-8 (61.0) against non-septic controls (Fig. 4B); IL-6 ranked first in both frameworks and in all scenarios (Fig. 4C).

**Figure 4.**
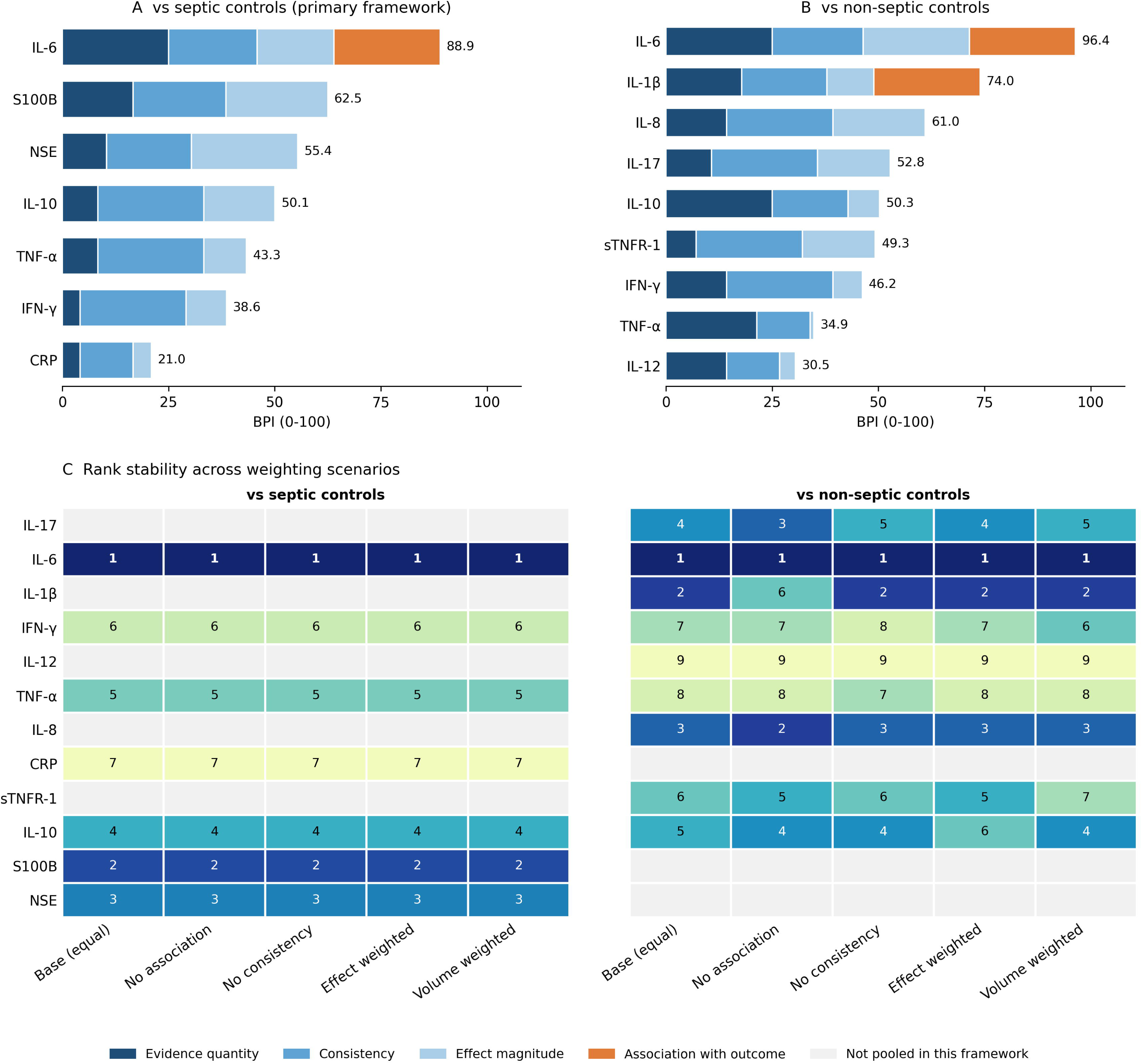
Biomarker prioritization index and its rank stability. **A**, BPI in the septic-control framework and **B**, in the non-septic-control framework, decomposed into the four weighted components, which sum to the value printed at the right of each bar. Components are rescaled within each framework, so scores are comparable within but not between panels. **C**, Rank of each biomarker under the base weighting and the four sensitivity scenarios in both frameworks; cells are shaded by rank within their framework, and gray cells indicate that the biomarker was not pooled in that framework. IL-6 holds rank 1 in all 5 scenarios in both frameworks.

## DISCUSSION

In 18 studies and 2388 participants, one mediator, IL-6, is supported concordantly across all three frameworks, and ranks first on an explicit priority index in both frameworks and under every weighting scenario. Five further markers discriminate encephalopathic from non-encephalopathic septic patients, S100B, NSE, IL-10, TNF-α and IFN-γ, and the two neuroglial markers do so in the pattern a barrier-and-injury account predicts; IL-6, S100B and NSE are also the estimates that survive omission of any single study.

These are claims about direction, not magnitude. Every computable prediction interval spans no difference and both directions, including IL-6 versus septic controls (−1.03 to 3.14), the arithmetic consequence of substantial between-study variance. What replicates is the sign of the difference and its concordance across frameworks, not the size of any pooled estimate [44, 45]. The very low certainty has a likely specific cause: not poor studies, since most are at low risk of bias, but an unstandardized assessed phenotype. Every pooled estimate averages over case definitions from CAM-ICU positivity to electroencephalographic abnormality, and the subgroup analyses show that those definitions do not identify populations with the same biomarker behavior.

The prespecified Th17 hypothesis has not yet received its discriminating test: IL-17 was modestly elevated against non-septic controls (SMD 0.37, 95% CI 0.01 to 0.73), a contrast that cannot separate encephalopathy from sepsis itself, and only 1 study measured it against septic controls, so the discriminating contrast could not be pooled for the axis at all. Three features mean this neither confirms nor refutes the hypothesis. The IL-17 evidence base is the smallest of any biomarker here (3 comparisons, 277 participants) and its prediction interval spans both directions; the axis was measured as circulating IL-17A alone, with no study reporting Th17 cell frequencies, the IL-23/IL-17 axis or the Th17/regulatory T-cell balance, and one reporting cerebrospinal fluid IL-17, while the mechanistic claim concerns a tissue-level process at the barrier [24, 25] that peripheral IL-17A may index poorly; and the pooled signal is not carried by the SAE stratum, which is null (SMD 0.20, 95% CI -0.14 to 0.53), but by a single SAD comparison, and is lost when either of two studies is omitted. The only T-helper-lineage cytokine discriminating against septic controls was IFN-γ, a Th1 signal from two studies. The data support instead that the upstream inducers carry the signal, IL-6 strongly, IL-1β weakly, compatible with a Th17-relevant process read out through its inducers or with unrelated IL-6 pathways, and non-inflammatory contributors such as impaired cerebral perfusion and endothelial activation can produce the same phenotype [69, 70].

Our IL-6, S100B and NSE results are directionally consistent with previous single-marker syntheses [29–31]; separating the frameworks shows that some markers behave differently in each, TNF-α and IFN-γ most clearly, which plausibly explains part of the inconsistency between prior reports, and stratifying on phenotype shows that heterogeneity previously attributed to assay and timing is partly attributable to case definition [12, 71].

For practice, none of these markers is ready to function as a diagnostic test. IL-6 is among the least specific analytes in critical care [17, 72], and the primary literature supplies no threshold, likelihood ratio or discrimination statistic. Implied discrimination sets the ceiling: against septic controls the values run from 0.57 to 0.85, which supports cohort enrichment for trials and triage to formal neurological assessment where electroencephalography cannot be offered to every patient, but not a threshold applied to an individual. The realistic near-term role is a multi-marker model combining an inflammatory index with a neuroglial-injury marker, evaluated against a standardized phenotype and reported with the discrimination and calibration metrics a diagnostic claim requires [73, 74]. For research, four gaps follow: the phenotype must be standardized and SAE and SAD reported separately; the septic-control contrast must become the default; the association framework is nearly empty, with eight of 12 biomarkers never assessed against the outcome; and testing the Th17 hypothesis requires studies that measure the axis rather than one cytokine, in the SAE phenotype, against septic controls.

Neurofilament light chain, the most advanced neuroaxonal marker in sepsis [75], could not be evaluated: none of the included studies measured it, and a search built around the Th17 axis was not designed to find it. No meta-analysis has yet pooled it in SAE or SAD [31, 76, 77], despite consistent associations with delirium in critical illness [78], so it merits a future dedicated, search-focused synthesis [9, 79].

This review covers the full mediator panel, keeps three non-commensurable frameworks separate and applies pre-stated, reproducible rules. Its limitations are substantial. Heterogeneity was high in most comparisons, so the pooled point estimates summarize a heterogeneous literature rather than estimate a common effect, which is what the prediction intervals state directly; many comparisons rest on two to four studies and samples below 400; and a substantial proportion of summaries were medians requiring conversion [37, 38], adding error not propagated into the intervals. The non-septic-control framework mixes healthy, non-septic and non-delirious comparators, many septic in the largest studies; three studies required digitization from figures, standard errors were converted to standard deviations, and one comparison required an imputed SD (eTables 13 and 16) [80]. An interaction test by type of case definition is not estimable, because 13 of the 21 strata reaching two studies are those with no stated instrument (eTable 11); the estimable contrast is the coarser one of eTable 12 and eFigure 4. Follow-up neurocognitive outcomes were outside the extraction, so whether these markers predict cognitive impairment after discharge, remains as a future research topic.

## CONCLUSIONS

To our knowledge, this is the first synthesis to evaluate the inflammatory, T-helper and neuroglial-injury panel in SAE and SAD while keeping the non-septic-control, septic-control and outcome-association frameworks separate, and that separation changes what the literature can be said to show. IL-6 emerges as the lead candidate: it is the only mediator supported concordantly by all three frameworks, its association with the neurological outcome is statistically clear and clinically interpretable, and it ranks first on a prespecified priority index under every weighting scenario. S100B and NSE discriminate encephalopathic from non-encephalopathic septic patients, identifying a neuroglial-injury signal that sepsis alone does not explain and making combined inflammatory and neuroglial panels the rational basis for diagnostic modeling and trial enrichment. The Th17 hypothesis remains open rather than refuted, and this review defines the study that would test it: the IL-23/IL-17 axis measured in SAE against septic controls. Because biomarker behavior differed between SAE and SAD strata, standardized phenotyping and septic-controlled designs are what will turn these signals into potential clinically usable tools.

## Data Availability

All data produced in the present study are available upon reasonable request to the authors

## ACKNOWLEDGMENTS

The authors thank the Brazilian agencies CNPq, CAPES and FUNCAP for the scholarships of the students involved in this study.

## Author contributions

A.C.-F. conceived the study, developed the methodology, supervised the review, performed the analyses, interpreted the findings and drafted the manuscript. E.F.N., L.D.C.G. and F.J.R.D.F. performed literature screening, data extraction and risk-of-bias assessment. L.D.C.G. and T.Q.O. contributed to data verification and interpretation. D.S.M. contributed to the conception of the review, to the immunological interpretation of the mediator panel and to critical revision of the manuscript. F.D.-P. contributed clinical and pathophysiological expertise in sepsis-associated encephalopathy, to the interpretation of the findings and to critical revision of the manuscript. M.E.A.M. and T.B. contributed supervision, interpretation and critical revision. All authors reviewed and approved the final manuscript.

## Data availability

All extracted and analyzed data are contained in the tables and supplemental digital content. The data layer and the table- and figure-generation scripts that produce every reported value are available from the corresponding author on request.

## Ethics

Not applicable: this is a synthesis of published aggregate data, with no individual participant data obtained.

## Registration

PROSPERO CRD420261371685.

## Use of generative artificial intelligence

Generative artificial-intelligence tools (Anthropic Claude) were used during manuscript preparation for language editing, for organization of the scientific text, and to write the document-generation scripts that derive tables and figures from the single data layer. The meta-analyses of record were executed by the authors in Review Manager and in R as described above. All AI-assisted output was independently checked and validated by the authors against the underlying data, the analysis output and the original sources. Generative AI was not used to determine eligibility, extract study data from primary reports, assess risk of bias, generate primary research data or draw scientific conclusions, and no unpublished third-party material or individual participant data were entered into any such tool. The authors are responsible for the content of the study.

## Abbreviations

AUC area under the receiver-operating-characteristic curve

BPI: biomarker prioritization index
CAM-ICU: Confusion Assessment Method for the Intensive Care Unit
CI: confidence interval
CRP: C-reactive protein
GRADE: Grading of Recommendations, Assessment, Development and Evaluations
IFN-γ: interferon-γ
IL: interleukin
NOS: Newcastle-Ottawa Scale
NSE: neuron-specific enolase
OR: odds ratio
PRISMA: Preferred Reporting Items for Systematic Reviews and Meta-Analyses
SAD: sepsis-associated delirium
SAE: sepsis-associated encephalopathy
SMD: standardized mean difference
TNF-α: tumor necrosis factor-α.

## SUPPLEMENTAL DIGITAL CONTENT

Supplemental digital content is merged into a single PDF with a table of contents, comprising eAppendix 1 (PRISMA 2020 checklist), eAppendix 2 (full methods), eTables 1–26 and eFigures 1-4.

