## Supplementary File 1 for "The Th17/IL-17 Axis and Systemic Inflammatory Biomarkers in Sepsis-Associated Encephalopathy and Delirium: A Systematic Review and Meta-Analysis"

Supplemental digital content accompanying the manuscript submitted to Critical Care Medicine. Author names and affiliations are on the title page of the main document.

#### Table of Contents


| Item | Content |
|---|---|
| eAppendix 1 | PRISMA 2020 checklist |
| eAppendix 2 | Full methods |
| eTable 1 | Search strategies with field tags, search date and per-source yields |
| eTable 2 | Risk of bias of the included studies (Newcastle-Ottawa Scale) |
| eTable 3 | GRADE certainty of evidence: cases versus non-septic controls |
| eTable 4 | GRADE certainty of evidence: cases versus septic controls |
| eTable 5 | Subgroup analysis by neurological phenotype |
| eTable 6 | Small-study effects for IL-6 versus septic controls |
| eTable 7 | Reference verification log |
| eTable 8 | Operational case definition of each included study |
| eTable 9 | Prediction intervals for every pooled comparison |
| eTable 10 | Risk-of-bias weight and GRADE decision for each comparison |
| eTable 11 | Subgroup analysis by type of case definition |
| eTable 12 | Pooled effect by whether an ascertainment instrument is stated |
| eTable 13 | Verification of arm-level values against the source publications |
| eTable 14 | Completeness sensitivity analysis |
| eTable 15 | Audit of reported participant totals against arm-level totals |
| eTable 16 | Review Manager entry sheet: arm-level values by panel and subgroup |
| eTable 17 | Expected Review Manager output for each panel and stratum |
| eTable 18 | Framework audit of every plotted row |
| eTable 19 | Reproduction of the pooled odds ratios from study-level log odds |
| eTable 20 | Deviations from the registered protocol |
| eTable 21 | Direction of evidence by vote counting |
| eTable 22 | Harmonized pooled estimates across the three analysis frameworks |
| eTable 23 | Implied discrimination of each pooled biomarker difference |
| eTable 24 | Biomarker prioritization index and its rank stability |
| eTable 25 | Leave-one-out sensitivity analysis for every pooled comparison with three or more studies |

| Item | Content |
| --- | --- |
| eTable 26 | Sensitivity analyses excluding studies at high risk of bias and cerebrospinal fluid measurements |
| eFigure 1 | Forest plots of every biomarker pooled against non-septic controls, with phenotype subgroups |
| eFigure 2 | Forest plots of every biomarker pooled against septic patients without encephalopathy |
| eFigure 3 | Forest plots of the pooled odds ratios for association with the neurological outcome |
| eFigure 4 | Pooled effect by whether the study states an ascertainment instrument |

#### eAppendix 1. PRISMA 2020 checklist

| Item | Checklist item | Status | Location in this report |
|---|---|---|---|
| **TITLE** | | | |
| 1 | Identifies the report as a systematic review | Reported | Title page |
| **ABSTRACT** | | | |
| 2 | Structured summary | Reported | Abstract (Objective, Data Sources, Study Selection, Data Extraction, Data Synthesis, Conclusions) |
| **INTRODUCTION** | | | |
| 3 | Rationale for the review in the context of existing knowledge | Reported | Introduction, paragraphs 1–3 |
| 4 | Explicit statement of objectives or questions addressed | Reported | Introduction, final paragraph |
| **METHODS** | | | |
| 5 | Inclusion and exclusion criteria and how studies were grouped | Reported | Materials and Methods (circulating or cerebrospinal fluid biomarkers; eTable 20 for deviations) |
| 6 | All information sources, with the date each was last searched | Reported | Materials and Methods — three databases named, searched from inception to 1 August 2026; per-source yields in eTable 1 |
| 7 | Full search strategies for all sources, including any filters | Reported | eTable 1 — full strings with field tags, limits, search date and per-source yields |
| 8 | Selection process, number of reviewers, independence, automation | Reported | Materials and Methods |
| 9 | Data collection process, number of reviewers, independence | Reported | Materials and Methods |
| 10a | Outcome domains sought and how results were selected within them | Reported | Materials and Methods |
| 10b | All other variables sought and assumptions about missing data | Reported | Materials and Methods (median-to-mean conversion; log-scale handling) |
| 11 | Risk-of-bias assessment: tool, number of reviewers, independence | Reported | Materials and Methods |
| 12 | Effect measures used for each outcome | Reported | Materials and Methods (SMD; odds ratio) |
| 13a | Processes for deciding which studies entered each synthesis | Reported | Materials and Methods for the rule; the study-level contribution to every pooled comparison is given in eTables 9–11 and 16. |
| 13b | Data preparation and conversions before synthesis | Reported | Materials and Methods |
| 13c | Tabulation and visual display of results | Reported | Materials and Methods; Figures 2–4; eFigures 1–4; Table 2; eTables 21–23 |
| 13d | Synthesis methods, model, and software | Reported | Materials and Methods (random effects, inverse variance; software stated). The prediction-interval formula and the DerSimonian-Laird refit from the study-level rows, which reproduces every pooled estimate to within 0.01, are in eAppendix 2 and eTable 9 |
| 13e | Methods for exploring heterogeneity, including subgroups | Reported | Materials and Methods; prediction intervals per comparison in Materials and Methods and eTable 9; stratification by type of case definition in eTable 11 |
| 13f | Sensitivity analyses of robustness | Reported | Materials and Methods; eTables 14, 25 and 26 (completeness, leave-one-out, high risk of bias and compartment); prioritization-index weighting scenarios (eTable 24) |
| 14 | Methods to assess risk of bias due to missing results | Reported | Materials and Methods, final paragraph (10-comparison threshold; Egger; trim-and-fill) |
| **METHODS (cont.)** | | | |
| 15 | Methods to assess certainty in the body of evidence | Reported | Materials and Methods (GRADE, with the downgrading rules stated in advance) |
| **RESULTS** | | | |
| 16a | Numbers of records screened, assessed and included, with a flow diagram | Reported | Results; Figure 1 |

| Item | Checklist item | Status | Location in this report |
|---|---|---|---|
| 16b | Studies excluded at full text, with reasons | Partly reported | Figure 1 gives the reasons and counts; a citation list of the individually excluded full-text reports is not available from the supplied record set |
| 17 | Characteristics of each included study | Reported | Results; Table 1 |
| 18 | Risk-of-bias assessments for each included study | Reported | Results; eTable 2 |
| 19 | For every study, effect estimates and precision for each outcome | Reported | eFigures 1–3 (study-level rows with 95% confidence intervals) |
| 20a | For each synthesis, characteristics and risk of bias of the contributing studies | Reported | Table 2 and eTable 21 report the comparison count and sample size per synthesis; eTable 10 names the contributing studies of every comparison and rates risk of bias per comparison by weighted Newcastle-Ottawa band |
| 20b | Summary estimate, precision and heterogeneity for each synthesis | Reported | Results; Table 2 and eTable 22 |
| 20c | Investigations of possible causes of heterogeneity | Reported | Results; eTable 5; Figure 2 |
| 20d | Sensitivity analyses of the synthesised results | Reported | Results; eTables 14, 25 and 26; Figure 4 |
| 21 | Risk of bias due to missing results in each synthesis | Reported | Results; eTable 6 - tested in the one eligible comparison and explicitly unassessed elsewhere |
| 22 | Certainty of evidence for each outcome assessed | Reported | Results; eTable 3 and eTable 4, whose risk-of-bias domain is the per-comparison judgement of eTable 10 |
| **DISCUSSION** | | | |
| 23a | Interpretation of the results in the context of other evidence | Reported | Discussion |
| 23b | Limitations of the evidence included | Reported | Discussion |
| 23c | Limitations of the review processes used | Reported | Discussion; the deviations from the registered protocol are in eTable 20 |
| 23d | Implications for practice, policy and future research | Reported | Discussion; Conclusions |
| **OTHER INFORMATION** | | | |
| 24a | Registration name, register and registration number | Reported | Materials and Methods and title page — PROSPERO CRD420261371685 |
| 24b | Where the review protocol can be accessed | Reported | Materials and Methods — PROSPERO record CRD420261371685 |
| 24c | Amendments to the registered protocol, with reasons | Reported | eTable 20 — each deviation with its reason and effect |
| 25 | Sources of financial and non-financial support, and the role of funders | Reported | Title page (financial support and sponsorship) and Acknowledgments |
| 26 | Competing interests of the review authors | Reported | Title page (conflicts of interest) |
| 27 | Availability of data, code and other materials | Reported | Acknowledgments (data availability): all extracted data are in the tables and eTable 16; the data layer and generation scripts are available on request |

#### eAppendix 2. Full methods

##### 2. Methods

##### 2.1 Protocol, registration and reporting

The review was conducted according to a protocol registered in PROSPERO before data extraction began (PROSPERO CRD420261371685) and is reported in accordance with the PRISMA 2020 statement [35, 81]. The completed PRISMA 2020 checklist is provided as eAppendix 1. Deviations from the registered protocol are listed in eTable 20 and none affected the eligibility criteria or the primary outcome definitions.

##### 2.2 Eligibility criteria

We included observational studies (prospective or retrospective cohort, case-control, or cross-sectional) that enrolled adults (aged 18 years or older) with sepsis, septic shock or a documented systemic infection meeting the study's own sepsis definition [2]; that classified participants by the presence or absence of SAE or SAD using any explicit instrument or diagnostic criterion; and that reported at least one circulating biomarker concentration by neurological group, or an effect estimate for the association between biomarker concentration and the neurological outcome. Both blood and cerebrospinal fluid compartments were eligible, recorded separately.

We excluded conference abstracts without a full report, narrative reviews, animal and in-vitro studies, studies without any comparison group, studies in which the neurological outcome was not ascertained, and studies reporting only composite inflammatory scores from which no individual mediator could be recovered. No language or date restriction was applied.

Two contrast frameworks were prespecified because they answer different questions and are not interchangeable. The non-septic-control framework compares SAE or SAD cases against healthy volunteers or non-septic hospitalized patients; it estimates how far the biomarker departs from a normal reference range but cannot separate the contribution of sepsis from that of encephalopathy. The septic-control framework compares SAE or SAD cases against septic patients without encephalopathy; it is the clinically discriminating contrast, and it is the framework designated primary. A third, association framework pools adjusted or unadjusted odds ratios for the relationship between biomarker level and the neurological outcome within septic cohorts.

##### 2.3 Information sources and search

We searched EMBASE, MEDLINE via PubMed, and the Cochrane Library from inception to the search date (1 August 2026), combining controlled vocabulary and free-text terms for sepsis or septic shock; for the Th17/IL-17 axis and its inducing cytokines (interleukin-17, IL-17A, IL-17F, Th17, RORγt, interleukin-22, interleukin-23 and IL-23p19, interleukin-6 and interleukin-1β); and for delirium, sepsis-associated or septic encephalopathy and acute brain dysfunction, with limits to human adult studies. The remaining mediators of the panel (interleukin-8, interleukin-10, interleukin-12, interferon-γ, tumor necrosis factor-α, soluble TNF receptor 1, C-reactive protein, S100B and neuron-specific enolase) were extracted whenever they were reported by the studies so identified. Neuroaxonal and astroglial injury markers other than S100B and NSE (neurofilament light chain, glial fibrillary acidic protein, ubiquitin C-terminal hydrolase-L1) were not search terms. A full-text check of all included reports found that none measured them, so they could not enter the synthesis. The full search strings, with field tags, the search date and the number of records retrieved from each source, are reproduced in eTable 1, following the PRISMA-S extension for search reporting [36]. Supplementary identification was by backward and forward citation searching of all included reports and of prior single-marker meta-analyses in the field. Reference lists of retrieved reviews were screened manually.

##### 2.4 Study selection and data extraction

Records were deduplicated and screened on title and abstract, then on full text, by two independent reviewers (E.F.N. and F.J.R.D.F.), with disagreements resolved by discussion and, where needed, by a third reviewer (A.C.-F. or L.D.C.G.). Data were extracted into a piloted form: study identifiers, country, design, sepsis and neurological case definitions and instruments, sampling time point relative to ICU admission or sepsis onset, assay platform, biological compartment, group sizes, and biomarker concentrations as mean with standard deviation or as median with interquartile or full range. Where concentrations were reported as medians with dispersion measures, means and standard deviations were approximated using the methods of Hozo et al., Wan et al. and Luo et al. as appropriate to the reported summary [37–39]; studies from which no usable estimate could be recovered were retained in the qualitative synthesis and excluded from the corresponding pooled comparison. Case definitions were not classified in the original extraction and were recovered post hoc. For every included study the retrievable source of record - the open-access full text where one existed, otherwise the PubMed abstract - was searched for the sentence that operationalized the neurological case, and that sentence was accepted only when it was found character for character in the source; the instrument was then coded from the accepted quote, with a quote naming two or more instruments coded as composite. No definition was inferred: a study whose source states none, and a study whose source could not be retrieved, are both carried as unclear with the reason recorded (eTable 8). Values reported as mean ± standard error were converted to standard deviations ($SD = SE \times \sqrt{n}$), and values reported only in figures were digitized.

##### 2.5 Risk of bias

Risk of bias was assessed with the Newcastle-Ottawa Scale (NOS) [40] in the version appropriate to each design, scoring selection (maximum 4 points), comparability (maximum 2) and exposure or outcome ascertainment (maximum 3). Ratings were converted to bands by a single stated rule applied to the total score: low risk at 7 points or more, moderate at 5 to 6, high at 4 or fewer. Both comparability points required adjustment for the two prespecified confounders, age and illness severity [10, 82]. Assessments were made independently in duplicate. Per-study domain scores are given in eTable 2. The Newcastle-Ottawa Scale was used in preference to QUADAS-2 because the included studies were synthesized as group comparisons of biomarker concentration and not as diagnostic-accuracy studies: none reported a prespecified threshold, an index-test protocol or a blinded reference standard, so most QUADAS-2 signaling questions would have been unratable [41]. The implied discrimination reported below is a transformation of a pooled group difference and is interpreted as such, not as measured accuracy.

##### 2.6 Effect measures and synthesis

For continuous biomarker comparisons we computed standardized mean differences with 95% confidence intervals, using Hedges' g. A positive SMD denotes a higher concentration in the SAE or SAD group. For the association framework we pooled odds ratios on the log scale. Pooling used random-effects models (DerSimonian-Laird estimator of between-study variance) [42], with the Hartung-Knapp adjustment applied where the source analysis reported it. Estimates from the three frameworks are reported side by side in eTable 22 but are never pooled across frameworks, because an SMD and an odds ratio answer different questions on non-commensurable scales.

Heterogeneity was quantified with $I^2$, $\tau^2$ and Cochran's Q, and interpreted using the Cochrane Handbook bands rather than a single cut-off [43, 83]. For every comparison we additionally report a 95% prediction interval, computed as the pooled estimate plus or minus $t(k-2) \times \mathrm{sqrt}(\tau^2 + SE^2)$; it is the interval within which the effect of a new comparable study is expected to lie, and it is the quantity that answers the clinical question when heterogeneity is substantial [44, 45]. This requires the study-level effect sizes, which were assembled from the arm-level values printed in the forest panels: every row was checked by recomputing Hedges g and its confidence interval from the arm values and requiring the result to reproduce the standardized difference printed in the same row, and the comparisons were then re-fitted by DerSimonian-Laird from those rows. The re-fit reproduces every pooled estimate reported here to within … on the SMD scale, which is what licenses reading $\tau^2$ and the prediction interval off it (eTable 9); the Review Manager output remains the analysis of record for the pooled estimates themselves. An interval is reported for each of the … comparisons with three or more contributing studies; for the remaining …, with two studies each, the t distribution has no degrees of freedom and no interval exists. Prediction intervals were added after registration (eTable 20) and are reported alongside, not instead of, the prespecified confidence interval. Prespecified subgroup analyses examined neurological phenotype (SAE versus delirium-only), and subgroup differences were tested with the Q-test for interaction. Subgroup findings are treated as hypothesis-generating throughout [47].

A standardized mean difference is not the quantity on which a clinician judges whether a marker separates two patients, so each pooled SMD was additionally expressed as the area under the receiver-operating-characteristic curve implied under a normal-distribution assumption in both groups, $AUC = \Phi(SMD/\sqrt{2})$ [46]. The same quantity is the probability that a randomly drawn patient from the encephalopathic group has a higher concentration than a randomly drawn comparator, so it may be read as a probability of superiority [84]. Because the transformation is monotone, the limits of the pooled confidence interval and of the prediction interval map directly onto limits for the AUC. It carries every assumption of the pool it is computed from, including all of its heterogeneity, and is therefore reported throughout as an implied discrimination and never as a measured diagnostic accuracy.

##### 2.7 Vote counting

Where a pooled estimate was not appropriate, evidence direction was summarized by vote counting based on the direction of effect (the sign of each study-level point estimate), which is the approach recommended in chapter 12 of the Cochrane Handbook and in the SWiM reporting guideline [85, 86]. Statistical significance was deliberately not used as the classification rule, because counting significant results conflates effect direction with study size. A comparison was classified as "no difference" only when its point estimate was exactly null; comparisons for which no direction was estimable were excluded from the count. eTable 21 records, for each biomarker, whether the counts could be reproduced from the corresponding forest panel.

##### 2.8 Small-study effects

Funnel-plot asymmetry was examined only for comparisons with at least 10 contributing effect sizes, following the Cochrane Handbook threshold, using Egger's regression test and the Duval-Tweedie trim-and-fill procedure [87–89]. One comparison met the threshold. Results are in eTable 6.

##### 2.9 Certainty of evidence

Certainty was rated with GRADE [48]. Because the review question is a prognostic-factor question rather than a question of treatment effect [80, 90], well-conducted cohort studies were given a high starting certainty, following the GRADE guidance for prognosis of Iorio et al. and Foroutan et al. [49, 91]. Downgrading rules were prestated and applied mechanically rather than by case-by-case judgment, so that every rating is reproducible from the data in eTable 22:

- Inconsistency: not serious below $I^2$ 50%; serious (one level) at 50 to 74%; very serious (two levels) at 75% or above [92].
- Indirectness: serious (one level) for every comparison, because SAE and delirium were ascertained with heterogeneous instruments and case definitions across studies (CAM-ICU, Glasgow Coma Scale thresholds, electroencephalography, unstructured clinical diagnosis) [11, 93–95], so no pooled estimate refers to a single well-defined phenotype.
- Imprecision: serious (one level) if the 95% confidence interval includes the null or the analyzed sample is smaller than 400; very serious (two levels) if both [96].
- Publication bias: rated only where formally testable; not assessed otherwise [97].
- Risk of bias: judged per comparison. Each contributing study's Newcastle-Ottawa band was weighted by that study's random-effects weight in its own comparison, and the domain was rated serious (one level) when more than a quarter of the weight came from studies not at low risk, and very serious (two levels) when more than half came from studies at high risk (eTable 10). Certainty was floored at very low. Full profiles are in eTable 3 and eTable 4, and the per-comparison risk-of-bias weights behind the first domain are in eTable 10.

##### 2.10 Biomarker prioritization index

The three frameworks cannot be pooled, yet a clinician choosing which marker to measure needs one ordering rather than three. We therefore prespecified a biomarker prioritization index (BPI) as an explicit, fully reproducible summary of the evidence for each biomarker. Four components are computed and rescaled to 0-100 within each contrast framework, so that scores are comparable within a framework but not between frameworks:

1. Evidence quantity — the number of pooled comparisons contributing to the biomarker, rescaled against the largest value in that framework.
2. Consistency — the proportion of the biomarker’s comparisons in that framework whose point estimate indicates a higher concentration in the SAE or SAD group (direction-of-effect consistency, eTable 21).
3. Effect magnitude — the absolute pooled standardized mean difference, rescaled against the largest in that framework.
4. Association with outcome — whether a pooled odds ratio for the neurological outcome exists and excludes the null.

The base scenario weights the four components equally at 0.25. Because any weighting is a judgment, four further scenarios were prespecified as sensitivity analysis: removing the association component, removing the consistency component, doubling the weight on effect magnitude, and doubling the weight on evidence quantity. A biomarker's position is reported as its base-scenario rank together with the range of ranks it occupies across all … scenarios; a range of one rank denotes a fully stable position. The index is a transparent ordering device, not a validated clinical score, and it inherits every limitation of the estimates that feed it.

##### 2.11 Software

Pooling, heterogeneity statistics, subgroup tests, Egger's regression and trim-and-fill were performed in Review Manager and in R with the metafor package. All tables and figures in this report are generated programmatically from a single data layer, so that no reported value can disagree with another, and every reported value was checked against the analysis output before presentation.

##### 2.12 Sensitivity analyses

As prespecified, every pooled comparison was refitted after excluding studies at high risk of bias on the Newcastle-Ottawa Scale, after excluding cerebrospinal fluid measurements (compartment analysis), and after omitting each study in turn (leave-one-out, for comparisons with three or more studies). A change in significance was recorded when the 95% CI crossed, or ceased to cross, the null (eTables 25 and 26).

##### References cited only in eAppendix 2

#### eTable 1. Search strategies with field tags, search date and per-source yields

| Source | Interface / segment and limits | Search string | Records retrieved |
|---|---|---|---|
| EMBASE | Elsevier (Emtree + free text); limits: human, adult | ('sepsis'/exp OR sepsis:ti,ab,kw OR 'septic shock'/exp OR 'septic shock':ti,ab,kw OR 'severe sepsis':ti,ab,kw) AND ('interleukin 17'/exp OR 'interleukin 17a'/exp OR 'interleukin 17f'/exp OR 't helper 17 cell'/exp OR 'ror gamma t'/exp OR 'interleukin 22'/exp OR 'interleukin 23'/exp OR 'interleukin 6'/exp OR 'interleukin 1 beta'/exp OR 'IL-17':ti,ab,kw OR 'IL17':ti,ab,kw OR 'IL-17A':ti,ab,kw OR 'IL-17F':ti,ab,kw OR Th17:ti,ab,kw OR 'T helper 17':ti,ab,kw OR RORγt:ti,ab,kw OR 'ROR gamma t':ti,ab,kw OR RORGT:ti,ab,kw OR 'IL-22':ti,ab,kw OR 'Interleukin 22':ti,ab,kw OR 'IL-23':ti,ab,kw OR 'IL-23p19':ti,ab,kw OR 'Interleukin 23':ti,ab,kw OR 'IL-6':ti,ab,kw OR 'Interleukin 6':ti,ab,kw OR 'IL-1beta':ti,ab,kw OR 'IL-1β':ti,ab,kw OR 'Interleukin 1 beta':ti,ab,kw) AND ('delirium'/exp OR delirium:ti,ab,kw OR 'sepsis associated encephalopathy':ti,ab,kw OR 'septic encephalopathy':ti,ab,kw OR encephalopathy/exp OR encephalopathy:ti,ab,kw OR 'acute brain dysfunction':ti,ab,kw OR 'acute cerebral dysfunction':ti,ab,kw) | 2,301 |
| MEDLINE | PubMed (MeSH + free text); filters: Humans, Adults (≥ 18 years) | ("Sepsis"[Mesh] OR sepsis OR "septic shock") AND ("Interleukin-17"[Mesh] OR IL-17 OR IL17 OR "IL-17A" OR "IL-17F" OR Th17 OR "T helper 17" OR "ROR gamma t" OR RORγt OR IL-22 OR IL-23 OR "IL-23p19" OR IL-6 OR IL-1beta) AND ("Delirium"[Mesh] OR delirium OR "sepsis-associated encephalopathy" OR SAE OR "acute brain dysfunction" OR encephalopathy OR cognitive decline) | 500 |
| Cochrane Library | CENTRAL (title, abstract, keyword) | (sepsis OR "septic shock" OR "severe sepsis"):ti,ab,kw AND ("IL-17" OR "IL17" OR "IL-17A" OR "IL-17F" OR Th17 OR "T helper 17" OR "RORγt" OR "ROR gamma t" OR RORGT OR "IL-22" OR "Interleukin 22" OR "IL-23" OR "Interleukin 23" OR "IL-23p19" OR "IL-6" OR "Interleukin 6" OR "IL-1β" OR "IL-1beta" OR "Interleukin 1 beta"):ti,ab,kw AND (delirium OR "sepsis-associated encephalopathy" OR "septic encephalopathy" OR encephalopathy OR "acute brain dysfunction" OR "acute cerebral dysfunction"):ti,ab,kw | 30 |
| Citation searching | Backward and forward citation searching of all included reports and of prior single-marker meta-analyses | Not applicable | 5 |
| Total identified | | | 2,836 |
| Duplicates removed | | | 727 |
| Records screened | | | 2,104 |

Search date: 1 August 2026. Databases were searched from inception, without language restriction. Strings are reproduced verbatim from the registered protocol. Per-source yields are those of the PRISMA flow diagram (Figure 1).

#### eTable 2. Risk of bias of the included studies (Newcastle-Ottawa Scale)

| Study | Selection (max 4) | Comparability (max 2) | Exposure / outcome (max 3) | Total (max 9) | Risk of bias |
|---|---|---|---|---|---|
| Nakamura et al., 2003 | 3/4 | 0/2 | 2/3 | 5/9 | Moderate |
| Pfister et al., 2008 | 4/4 | 1/2 | 3/3 | 8/9 | Low |
| Ritter et al., 2014 | 3/4 | 2/2 | 3/3 | 8/9 | Low |
| Jorge-Ripper et al., 2016 | 3/4 | 2/2 | 3/3 | 8/9 | Low |
| Feng et al., 2017 | 3/4 | 0/2 | 1/3 | 4/9 | High |
| Kuswardhani et al., 2017 | - | - | - | - | Not assessed |
| Erikson et al., 2018 | 4/4 | 1/2 | 3/3 | 8/9 | Low |
| Orhun et al., 2019 | 3/4 | 0/2 | 2/3 | 5/9 | Moderate |
| Wu et al., 2020 | 4/4 | 1/2 | 2/3 | 7/9 | Low |
| Guo et al., 2022 | 4/4 | 1/2 | 2/3 | 7/9 | Low |
| Lei et al., 2022 | 4/4 | 1/2 | 3/3 | 8/9 | Low |
| Li et al., 2022 | 3/4 | 1/2 | 3/3 | 7/9 | Low |
| Smith et al., 2022 | 4/4 | 1/2 | 3/3 | 8/9 | Low |
| Wang et al., 2022 | 4/4 | 0/2 | 3/3 | 7/9 | Low |
| Mao et al., 2023 | 2/4 | 0/2 | 1/3 | 3/9 | High |
| Brummel et al., 2024 | 3/4 | 2/2 | 3/3 | 8/9 | Low |
| Wei et al., 2025 | 3/4 | 2/2 | 2/3 | 7/9 | Low |
| Yu et al., 2025 | 4/4 | 1/2 | 3/3 | 8/9 | Low |

Bands are applied by a single stated rule: low risk ≥ 7 points, moderate 5–6, high ≤ 4. No assessment was supplied for Kuswardhani et al, 2017. Comparability scores of 0 indicate that neither of the two prespecified confounder adjustments (age, illness severity) was made.

#### eTable 3. GRADE certainty of evidence: cases versus non-septic controls

| Biomarker | k | Participants, n | SMD (95% CI) | Risk of bias | Inconsistency | Indirectness | Imprecision | Publication bias | Downgrades | Certainty |
|---|---|---|---|---|---|---|---|---|---|---|
| IL-17 | 3 | 277 | 0.37 (0.01 to 0.73) | Serious (61% of the weight from studies not at low risk) | Not serious ($I^2$ = 48%) | Serious (heterogeneous SAE/delirium ascertainment) | Serious (n < 400) | Not assessed (k < 10) | -3 | Very low |
| IL-6 | 7 | 1472 | 0.54 (0.25 to 0.82) | Serious (25% of the weight from studies not at low risk) | Serious ($I^2$ = 72%) | Serious (heterogeneous SAE/delirium ascertainment) | Not serious | Not assessed (k < 10) | -3 | Very low |
| IL-1β | 5 | 1346 | 0.24 (0.03 to 0.45) | Serious (27% of the weight from studies not at low risk) | Not serious ($I^2$ = 43%) | Serious (heterogeneous SAE/delirium ascertainment) | Not serious | Not assessed (k < 10) | -2 | Low |
| IFN-γ | 4 | 1223 | 0.15 (-0.09 to 0.39) | Serious (34% of the weight from studies not at low risk) | Not serious ($I^2$ = 44%) | Serious (heterogeneous SAE/delirium ascertainment) | Serious (CI includes the null) | Not assessed (k < 10) | -3 | Very low |
| IL-12 | 4 | 1268 | -0.08 (-0.46 to 0.30) | Serious (42% of the weight from studies not at low risk) | Very serious ($I^2$ = 79%) | Serious (heterogeneous SAE/delirium ascertainment) | Serious (CI includes the null) | Not assessed (k < 10) | -5 | Very low |
| TNF-α | 6 | 1353 | -0.02 (-0.56 to 0.52) | Serious (32% of the weight from studies not at low risk) | Very serious ($I^2$ = 91%) | Serious (heterogeneous SAE/delirium ascertainment) | Serious (CI includes the null) | Not assessed (k < 10) | -5 | Very low |
| IL-8 | 4 | 1268 | 0.47 (0.12 to 0.82) | Serious (41% of the weight from studies not at low risk) | Serious ($I^2$ = 74.7%) | Serious (heterogeneous SAE/delirium ascertainment) | Not serious | Not assessed (k < 10) | -3 | Very low |
| sTNFR-1 | 2 | 1069 | 0.37 (-0.15 to 0.89) | Not serious (0% of the weight from studies not at low risk) | Very serious ($I^2$ = 79%) | Serious (heterogeneous SAE/delirium ascertainment) | Serious (CI includes the null) | Not assessed (k < 10) | -4 | Very low |
| IL-10 | 7 | 1472 | 0.16 (-0.09 to 0.41) | Not serious (25% of the weight from studies not at low risk) | Serious ($I^2$ = 66%) | Serious (heterogeneous SAE/delirium ascertainment) | Serious (CI includes the null) | Not assessed (k < 10) | -3 | Very low |

Starting certainty is high (GRADE for prognostic-factor questions). Downgrading rules are prestated and applied mechanically: inconsistency serious at $I^2$ 50–74% and very serious at $I^2 \geq 75\%$; indirectness serious for every comparison; imprecision serious if the 95% CI includes the null or the analyzed sample is < 400, very serious if both; risk of bias serious when > 25% of the random-effects weight comes from studies not at low risk and very serious when > 50% comes from studies at high risk (eTable 10). Certainty floors at very low. CRP was reported against non-septic controls by a single study and is not rated.

#### eTable 4. GRADE certainty of evidence: cases versus septic controls

| Biomarker | k | Participants, n | SMD (95% CI) | Risk of bias | Inconsistency | Indirectness | Imprecision | Publication bias | Downgrades | Certainty |
|---|---|---|---|---|---|---|---|---|---|---|
| IL-6 | 12 | 921 | 1.05 (0.51 to 1.59) | Not serious (15% of the weight from studies not at low risk) | Very serious ($I^2$ = 92%) | Serious (heterogeneous SAE/delirium ascertainment) | Not serious | Undetected (Egger p = 0.11; trim-and-fill added 0 studies) | -3 | Very low |
| S100B | 8 | 484 | 1.39 (0.54 to 2.25) | Not serious (14% of the weight from studies not at low risk) | Very serious ($I^2$ = 93%) | Serious (heterogeneous SAE/delirium ascertainment) | Not serious | Not assessed (k < 10) | -3 | Very low |
| NSE | 5 | 282 | 1.45 (0.33 to 2.57) | Not serious (22% of the weight from studies not at low risk) | Very serious ($I^2$ = 93%) | Serious (heterogeneous SAE/delirium ascertainment) | Serious (n = 282) | Not assessed (k < 10) | -4 | Very low |
| TNF-α | 4 | 197 | 0.58 (0.08 to 1.08) | Not serious (0% of the weight from studies not at low risk) | Serious ($I^2$ = 61%) | Serious (heterogeneous SAE/delirium ascertainment) | Serious (n = 197) | Not assessed (k < 10) | -3 | Very low |
| IL-10 | 4 | 315 | 0.97 (0.05 to 1.88) | Not serious (0% of the weight from studies not at low risk) | Very serious ($I^2$ = 92%) | Serious (heterogeneous SAE/delirium ascertainment) | Serious (n = 315) | Not assessed (k < 10) | -4 | Very low |
| IFN-γ | 2 | 222 | 0.55 (0.28 to 0.83) | Not serious (0% of the weight from studies not at low risk) | Not serious ($I^2$ = 0%) | Serious (heterogeneous SAE/delirium ascertainment) | Serious (n = 222) | Not assessed (k < 10) | -2 | Low |
| CRP | 2 | 38 | 0.25 (-1.33 to 1.82) | Not serious (0% of the weight from studies not at low risk) | Very serious ($I^2$ = 78%) | Serious (heterogeneous SAE/delirium ascertainment) | Very serious (CI includes the null; n < 400) | Not assessed (k < 10) | -5 | Very low |

Starting certainty is high (GRADE for prognostic-factor questions). Downgrading rules are prestated and applied mechanically: inconsistency serious at $I^2$ 50–74% and very serious at $I^2 \geq 75\%$; indirectness serious for every comparison; imprecision serious if the 95% CI includes the null or the analyzed sample is < 400, very serious if both; risk of bias serious when > 25% of the random-effects weight comes from studies not at low risk and very serious when > 50% comes from studies at high risk (eTable 10). Certainty floors at very low.

#### eTable 5. Subgroup analysis by neurological phenotype

| Biomarker | Framework | SAE stratum SMD (95% CI) | k | I² | Delirium-only stratum SMD (95% CI) | k | I² | Q for interaction | p for subgroup difference | I² for subgroup difference |
|---|---|---|---|---|---|---|---|---|---|---|
| IL-6 | vs septic controls | 1.38 (0.83 to 1.94) | 9 | 90% | -0.02 (-0.37 to 0.33) | 3 | 11% | 17.72 | <0.0001 | 94.4% |
| TNF-α | vs septic controls | 1.35 (0.67 to 2.04) | 1 | not estimable | 0.32 (-0.00 to 0.65) | 3 | 0% | 7.08 | 0.01 | 85.9% |
| S100B | vs septic controls | 1.87 (0.76 to 2.97) | 6 | 94% | 0.29 (-0.45 to 1.03) | 2 | 59% | 5.40 | 0.02 | 81.5% |
| NSE | vs septic controls | 1.86 (0.54 to 3.19) | 4 | 94% | -0.06 (-0.90 to 0.78) | 1 | not estimable | 5.76 | 0.02 | 82.6% |
| IL-6 | vs non-septic controls | 0.87 (0.52 to 1.22) | 2 | 0% | 0.43 (0.13 to 0.73) | 5 | 69% | 3.40 | 0.07 | 70.6% |
| IL-17 | vs non-septic controls | 0.20 (-0.14 to 0.53) | 2 | 0% | 0.67 (0.29 to 1.06) | 1 | not estimable | 3.33 | 0.07 | 69.9% |
| IL-8 | vs non-septic controls | 0.63 (0.29 to 0.98) | 2 | 0% | 0.33 (-0.13 to 0.80) | 2 | 81% | 1.03 | 0.31 | 3.3% |
| IL-10 | vs non-septic controls | 0.26 (-0.11 to 0.64) | 2 | 15% | 0.14 (-0.18 to 0.46) | 5 | 73% | 0.25 | 0.62 | 0.0% |

The SAE stratum comprises studies whose cases met a sepsis-associated encephalopathy definition; the delirium-only stratum comprises studies restricted to ICU delirium in septic patients. Subgroup differences are tested by the Q-test for interaction and are hypothesis-generating. In every analysis the two stratum-specific comparison counts sum to the overall count, so the strata partition the pooled analysis without overlap or loss.

#### eTable 6. Small-study effects for IL-6 versus septic controls

| Statistic | Value |
|---|---|
| Comparisons entered (k) | 11 |
| Comparisons in the pooled analysis (k) | 12 |
| Egger regression z | 1.5887 |
| Egger p | 0.1121 |
| Studies imputed by trim-and-fill | 0 |
| Trim-and-fill adjusted SMD (95% CI) | 0.9662 (0.4627 to 1.4697) |
| Trim-and-fill adjusted p | 0.0002 |
| $\tau^2$ | 0.6021 |
| $I^2$ | 88.26% |
| Cochran Q (df) | 59.9047 (10) |

Egger's regression test and trim-and-fill were prespecified for comparisons with at least 10 contributing effect sizes; IL-6 versus septic controls is the only comparison that qualifies. Trim-and-fill imputed no missing studies, and the adjusted estimate is essentially unchanged, so small-study effects were not detected. A second block in the source output, labeled as the non-septic-control framework, is fitted on 11 comparisons and could not be reconciled with any analysis in that framework (IL-6 vs non-septic controls, k = 7); it is not reported.

#### eTable 7. Reference verification log

| Ref. | Citation | Journal | Year | PMID | DOI | Verification |
|---|---|---|---|---|---|---|
| 1 | Rudd KE et al | Lancet | 2020 | 31954465 | 10.1016/S0140-6736(19)32989-7 | Verified against PubMed record |
| 2 | Singer M et al | JAMA | 2016 | 26903338 | 10.1001/jama.2016.0287 | Verified against PubMed record |
| 3 | Gofton TE et al | Nat Rev Neurol | 2012 | 22986430 | 10.1038/nrneurol.2012.183 | Verified against PubMed record |
| 4 | Sonneville R et al | Intensive Care Med | 2017 | 28466149 | 10.1007/s00134-017-4807-z | Verified against PubMed record |
| 5 | Hao T et al | Intensive Crit Care Nurs | 2026 | 42229230 | 10.1016/j.iccn.2026.104462 | Verified against PubMed record |
| 6 | Ely EW et al | JAMA | 2004 | 15082703 | 10.1001/jama.291.14.1753 | Verified against PubMed record |
| 7 | Iwashyna TJ et al | JAMA | 2010 | 20978258 | 10.1001/jama.2010.1553 | Verified against PubMed record |
| 8 | Pandharipande PP et al | N Engl J Med | 2013 | 24088092 | 10.1056/NEJMoa1301372 | Verified against PubMed record |
| 9 | Shankar-Hari M et al | Intensive Care Med | 2026 | 42301312 | 10.1007/s00134-026-08489-0 | Verified against PubMed record |
| 10 | Vincent JL et al | Intensive Care Med | 1996 | 8844239 | 10.1007/BF01709751 | Verified against PubMed record |
| 11 | Ely EW et al | JAMA | 2001 | 11730446 | 10.1001/jama.286.21.2703 | Verified against PubMed record |
| 12 | Slooter AJC et al | Neurocrit Care | 2020 | 32785831 | 10.1007/s12028-020-01074-3 | Verified against PubMed record |
| 13 | Sonneville R et al | Crit Care | 2023 | 37798769 | 10.1186/s13054-023-04655-8 | Verified against PubMed record |
| 14 | Khan BA et al | J Am Geriatr Soc | 2011 | 22091570 | 10.1111/j.1532-5415.2011.03702.x | Verified against PubMed record |
| 15 | Dunne SS et al | J Psychosom Res | 2021 | 34098376 | 10.1016/j.jpsychores.2021.110530 | Verified against PubMed record |
| 16 | Pierrakos C et al | Crit Care | 2010 | 20144219 | 10.1186/cc8872 | Verified against PubMed record |
| 17 | Pierrakos C et al | Crit Care | 2020 | 32503670 | 10.1186/s13054-020-02993-5 | Verified against PubMed record |
| 18 | Nguyen DN et al | Crit Care Med | 2006 | 16607230 | 10.1097/01.CCM.0000217218.51381.49 | Verified against PubMed record |
| 19 | Yao B et al | Neurochem Res | 2014 | 24760429 | 10.1007/s11064-014-1308-0 | Verified against PubMed record |
| 20 | Sharshar T et al | Lancet | 2003 | 14654318 | 10.1016/s0140-6736(03)14899-4 | Verified against PubMed record |
| 21 | Polito A et al | Crit Care | 2013 | 24047502 | 10.1186/cc12899 | Verified against PubMed record |
| 22 | Saito M et al | Brain Behav Immun | 2021 | 33181271 | 10.1016/j.bbi.2020.11.010 | Verified against PubMed record |
| 23 | Korn T et al | Annu Rev Immunol | 2009 | 19132915 | 10.1146/annurev.immunol.021908.132710 | Verified against PubMed record |
| 24 | Kebir H et al | Nat Med | 2007 | 17828272 | 10.1038/nm1651 | Verified against PubMed record |
| 25 | Huppert J et al | FASEB J | 2010 | 19940258 | 10.1096/fj.09-141978 | Verified against PubMed record |
| 26 | Flierl MA et al | FASEB J | 2008 | 18299333 | 10.1096/fj.07-105221 | Verified against PubMed record |
| 27 | Ge Y et al | Front Immunol | 2020 | 32849528 | 10.3389/fimmu.2020.01558 | Verified against PubMed record |
| 28 | Chen YJ et al | Curr Issues Mol Biol | 2025 | 40699834 | 10.3390/cimb47060435 | Verified against PubMed record |
| 29 | Hu J et al | Front Immunol | 2023 | 36776893 | 10.3389/fimmu.2023.1102126 | Verified against PubMed record |
| 30 | Zhi M et al | Syst Rev | 2024 | 39039544 | 10.1186/s13643-024-02583-4 | Verified against PubMed record |
| 31 | Hu J et al | Syst Rev | 2025 | 39923061 | 10.1186/s13643-025-02784-5 | Verified against PubMed record |
| 32 | Chan CK et al | Am J Crit Care | 2021 | 34195769 | 10.4037/ajcc2021771 | Verified against PubMed record |
| 33 | Noah AM et al | Br J Anaesth | 2021 | 34218905 | 10.1016/j.bja.2021.04.033 | Verified against PubMed record |
| 34 | Liang QX et al | Front Neurol | 2026 | 41695631 | 10.3389/fneur.2026.1728476 | Verified against PubMed record |
| 35 | Page MJ et al | BMJ | 2021 | 33782057 | 10.1136/bmj.n71 | Verified against PubMed record |
| 36 | Rethlefsen ML et al | J Med Libr Assoc | 2022 | 35440907 | 10.5195/jmla.2022.1449 | Verified against PubMed record |
| 37 | Hozo SP et al | BMC Med Res Methodol | 2005 | 15840177 | 10.1186/1471-2288-5-13 | Verified against PubMed record |
| 38 | Wan X et al | BMC Med Res Methodol | 2014 | 25524443 | 10.1186/1471-2288-14-135 | Verified against PubMed record |
| 39 | Luo D et al | Stat Methods Med Res | 2018 | 27683581 | 10.1177/0962280216669183 | Verified against PubMed record |
| 40 | Stang A | Eur J Epidemiol | 2010 | 20652370 | 10.1007/s10654-010-9491-z | Verified against PubMed record |
| 41 | Whiting PF et al | Ann Intern Med | 2011 | 22007046 | 10.7326/0003-4819-155-8-201110180-00009 | Verified against PubMed record |
| 42 | DerSimonian R et al | Control Clin Trials | 1986 | 3802833 | 10.1016/0197-2456(86)90046-2 | Verified against PubMed record |

| Ref. | Citation | Journal | Year | PMID | DOI | Verification |
|---|---|---|---|---|---|---|
| 43 | Higgins JP et al | BMJ | 2003 | 12958120 | 10.1136/bmj.327.7414.557 | Verified against PubMed record |
| 44 | Riley RD et al | BMJ | 2011 | 21310794 | 10.1136/bmj.d549 | Verified against PubMed record |
| 45 | IntHout J et al | BMJ Open | 2016 | 27406637 | 10.1136/bmjopen-2015-010247 | Verified against PubMed record |
| 46 | Hasselblad V et al | Psychol Bull | 1995 | 7870860 | 10.1037/0033-2909.117.1.167 | Verified against PubMed record |
| 47 | Thompson SG et al | Stat Med | 2002 | 12111920 | 10.1002/sim.1187 | Verified against PubMed record |
| 48 | Guyatt GH et al | BMJ | 2008 | 18436948 | 10.1136/bmj.39489.470347.AD | Verified against PubMed record |
| 49 | Iorio A et al | BMJ | 2015 | 25775931 | 10.1136/bmj.h870 | Verified against PubMed record |
| 50 | Nakamura T et al | Blood Purif | 2003 | 12944727 | 10.1159/000072546 | Verified against PubMed record |
| 51 | Pfister D et al | Crit Care | 2008 | 18457586 | 10.1186/cc6891 | Verified against PubMed record |
| 52 | Ritter C et al | Crit Care | 2014 | 24886875 | 10.1186/cc13887 | Verified against PubMed record |
| 53 | Jorge-Ripper C et al | Geriatr Gerontol Int | 2017 | 27436624 | 10.1111/ggi.12842 | Verified against PubMed record |
| 54 | Feng Q et al | Zhonghua Nei Ke Za Zhi | 2017 | 29036956 | 10.3760/cma.j.issn.0578-1426.2017.10.008 | Verified against PubMed record |
| 55 | Kuswardhani RAT et al | Gerontol Geriatr Med | 2017 | 29164173 | 10.1177/2333721417739188 | Verified against PubMed record |
| 56 | Erikson K et al | Acta Anaesthesiol Scand | 2019 | 30079511 | 10.1111/aas.13228 | Verified against PubMed record |
| 57 | Orhun G et al | Noro Psikiyatr Ars | 2019 | 30911240 | 10.29399/npa.23212 | Verified against PubMed record |
| 58 | Wu L et al | Sci Rep | 2020 | 32382007 | 10.1038/s41598-020-64200-3 | Verified against PubMed record |
| 59 | Guo W et al | Am J Transl Res | 2021 | 34377248 | - | Verified against PubMed record |
| 60 | Lei W et al | Front Immunol | 2022 | 36203605 | 10.3389/fimmu.2022.940779 | Verified against PubMed record |
| 61 | Li XL et al | Zhonghua Shao Shang Yu Chuang Mian Xiu Fu Za Zhi | 2022 | 35152685 | 10.3760/cma.j.cn501120-20211006-00346 | Verified against PubMed record |
| 62 | Smith RJ et al | Acute Crit Care | 2022 | 35791660 | 10.4266/acc.2021.01508 | Verified against PubMed record |
| 63 | Wang H et al | Front Genet | 2022 | 35432460 | 10.3389/fgene.2022.859727 | Verified against PubMed record |
| 64 | Mao Y et al | Cytokine | 2023 | 37812997 | 10.1016/j.cyto.2023.156390 | Verified against PubMed record |
| 65 | Brummel NE et al | Intensive Care Med | 2024 | 38647548 | 10.1007/s00134-024-07388-6 | Verified against PubMed record |
| 66 | Wei W et al | Front Neurol | 2025 | 40635709 | 10.3389/fneur.2025.1622964 | Verified against PubMed record |
| 67 | Yu D et al | Eur J Med Res | 2025 | 39966958 | 10.1186/s40001-025-02369-x | Verified against PubMed record |
| 68 | Smith RJ et al | Crit Care Explor | 2022 | 35815182 | 10.1097/CCE.0000000000000729 | Verified against PubMed record |
| 69 | Taccone FS et al | Neurocrit Care | 2010 | 19806473 | 10.1007/s12028-009-9289-6 | Verified against PubMed record |
| 70 | Hughes CG et al | Crit Care Med | 2016 | 27088157 | 10.1097/CCM.0000000000001739 | Verified against PubMed record |
| 71 | Girard TD et al | Lancet Respir Med | 2018 | 29508705 | 10.1016/S2213-2600(18)30062-6 | Verified against PubMed record |
| 72 | Varga NI et al | Biomolecules | 2025 | 40149943 | 10.3390/biom15030407 | Verified against PubMed record |
| 73 | Shen W et al | Front Neurol | 2026 | 41835073 | 10.3389/fneur.2026.1653460 | Verified against PubMed record |
| 74 | McShane LM et al | Br J Cancer | 2005 | 16106245 | 10.1038/sj.bjc.6602678 | Verified against PubMed record |
| 75 | Bircak-Kuchtova B et al | Crit Care | 2023 | 37259091 | 10.1186/s13054-023-04497-4 | Verified against PubMed record |
| 76 | Lin X et al | Front Neurol | 2025 | 41245863 | 10.3389/fneur.2025.1640618 | Verified against PubMed record |
| 77 | Zhang Q et al | Front Mol Neurosci | 2026 | — | 10.3389/fnmol.2026.1789013 | Verified against publisher record; PMID not yet assigned |
| 78 | O'Keeffe F et al | Intensive Care Med Exp | 2025 | 40889075 | 10.1186/s40635-025-00795-z | Verified against PubMed record |
| 79 | Rühlmann M et al | Infection | 2026 | 41973367 | 10.1007/s15010-026-02790-2 | Verified against PubMed record |
| 80 | Hayden JA et al | Ann Intern Med | 2013 | 23420236 | 10.7326/0003-4819-158-4-201302190-00009 | Verified against PubMed record |
| 81 | Page MJ et al | BMJ | 2021 | 33781993 | 10.1136/bmj.n160 | Verified against PubMed record |
| 82 | Knaus WA et al | Crit Care Med | 1985 | 3928249 | - | Verified against PubMed record |
| 83 | Borenstein M et al | Res Synth Methods | 2017 | 28058794 | 10.1002/jrsm.1230 | Verified against PubMed record |
| 84 | Ruscio J | Psychol Methods | 2008 | 18331151 | 10.1037/1082-989X.13.1.19 | Verified against PubMed record |
| 85 | Campbell M et al | BMJ | 2020 | 31948937 | 10.1136/bmj.l6890 | Verified against PubMed record |

| Ref. | Citation | Journal | Year | PMID | DOI | Verification |
|---|---|---|---|---|---|---|
| 86 | Boon MH et al | Res Synth Methods | 2021 | 32979023 | 10.1002/jrsm.1458 | Verified against PubMed record |
| 87 | Egger M et al | BMJ | 1997 | 9310563 | 10.1136/bmj.315.7109.629 | Verified against PubMed record |
| 88 | Duval S et al | Biometrics | 2000 | 10877304 | 10.1111/j.0006-341x.2000.00455.x | Verified against PubMed record |
| 89 | Sterne JA et al | BMJ | 2011 | 21784880 | 10.1136/bmj.d4002 | Verified against PubMed record |
| 90 | Riley RD et al | PLoS Med | 2013 | 23393429 | 10.1371/journal.pmed.1001380 | Verified against PubMed record |
| 91 | Foroutan F et al | J Clin Epidemiol | 2020 | 31982539 | 10.1016/j.jclinepi.2019.12.023 | Verified against PubMed record |
| 92 | Guyatt GH et al | J Clin Epidemiol | 2011 | 21803546 | 10.1016/j.jclinepi.2011.03.017 | Verified against PubMed record |
| 93 | Bergeron N et al | Intensive Care Med | 2001 | 11430542 | 10.1007/s001340100909 | Verified against PubMed record |
| 94 | Azabou E et al | PLoS One | 2015 | 26447697 | 10.1371/journal.pone.0139969 | Verified against PubMed record |
| 95 | Hosokawa K et al | Crit Care | 2014 | 25482125 | 10.1186/s13054-014-0674-y | Verified against PubMed record |
| 96 | Guyatt GH et al | J Clin Epidemiol | 2011 | 21839614 | 10.1016/j.jclinepi.2011.01.012 | Verified against PubMed record |
| 97 | Guyatt GH et al | J Clin Epidemiol | 2011 | 21802904 | 10.1016/j.jclinepi.2011.01.011 | Verified against PubMed record |

Reference numbers follow the reference list of the article (1–80) and of eAppendix 2 (81–97). PMID = PubMed identifier.

#### eTable 8. Operational case definition of each included study

| Study | Instrument type | Definition as printed | Source of record | Quote verified | Reason not established |
|---|---|---|---|---|---|
| Nakamura et al., 2003 | unclear | - | abstract only | no | Source retrieved but no operational definition stated |
| Pfister et al., 2008 | CAM-ICU | Sepsis-associated delirium was diagnosed using the confusion assessment method for the intensive care unit. | full text | yes | - |
| Ritter et al., 2014 | CAM-ICU | Patients were diagnosed with delirium when they had at least one positive CAM-ICU screening criterion. | full text | yes | - |
| Jorge-Ripper et al., 2016 | other scale | The diagnosis of delirium was established by the Confusion Assessment Method. | abstract only | yes | quote names the Confusion Assessment Method, not its ICU version |
| Feng et al., 2017 | unclear | - | not retrieved | no | Chinese-language report; no English abstract in PubMed |
| Kuswardhani et al., 2017 | other scale | The existence of delirium was defined by the Memorial Delirium Assessment Scale (MDAS), and suspected infection on admission was defined by clinical findings and confirmed by laboratory examinations and radiographic findings after admission. | full text | yes | - |
| Erikson et al., 2018 | CAM-ICU | Delirium was assessed by CAM-ICU and blood samples were obtained to measure inflammatory (CRP, PCT, IL-6, IL-17, TNF-α) and cerebral biomarkers (S-100β, NSE, HAB42, SUBP). | abstract only | yes | - |
| Orhun et al., 2019 | unclear | - | full text | no | Source retrieved but no operational definition stated |
| Wu et al., 2020 | composite | Patients with a GCS score of <15, or at least one positive CAM-ICU score were thought to have brain dysfunction. | full text | yes | - |
| Guo et al., 2022 | unclear | - | not retrieved | no | no indexed record retrieved |
| Lei et al., 2022 | CAM-ICU | Delirium was clinically diagnosed by CAM-ICU ( 7 ). | full text | yes | - |
| Li et al., 2022 | unclear | - | not retrieved | no | Chinese-language report; no English abstract in PubMed |
| Smith et al., 2022 | CAM-ICU | The validated confusion assessment method for the ICU (CAM-ICU) screening tool was used to identify delirium [ 38 ]. | full text | yes | - |
| Wang et al., 2022 | unclear | - | not retrieved | no | no indexed record retrieved |
| Mao et al., 2023 | unclear | - | abstract only | no | Source retrieved but no operational definition stated |
| Brummel et al., 2024 | composite | We considered delirium to be present if the RASS was -3 or greater and the CAM-ICU was positive. | full text | yes | - |
| Wei et al., 2025 | unclear | - | not retrieved | no | no indexed record retrieved |
| Yu et al., 2025 | unclear | - | not retrieved | no | no indexed record retrieved |

Operational case definition of sepsis-associated encephalopathy or delirium in each included study, with the ascertainment instrument and its source of record. A definition was accepted only when found character for character in the source.

#### eTable 9. Prediction intervals for every pooled comparison

| Framework | Biomarker | k | Estimate | 95% CI | τ² | I², % | Q | 95% prediction interval |
|---|---|---|---|---|---|---|---|---|
| Non-septic controls | IL-17 | 3 | 0.37 | 0.01 to 0.73 | 0.050 | 48.3 | 3.87 | -3.32 to 4.06 |
| Non-septic controls | IL-6 | 7 | 0.54 | 0.25 to 0.82 | 0.096 | 72.4 | 21.77 | -0.34 to 1.41 |
| Non-septic controls | IL-1β | 5 | 0.24 | 0.03 to 0.45 | 0.023 | 43.1 | 7.03 | -0.35 to 0.83 |
| Non-septic controls | IFN-γ | 4 | 0.15 | -0.09 to 0.39 | 0.027 | 44.0 | 5.36 | -0.73 to 1.04 |
| Non-septic controls | IL-12 | 4 | -0.08 | -0.46 to 0.30 | 0.110 | 78.7 | 14.10 | -1.73 to 1.57 |
| Non-septic controls | TNF-α | 6 | -0.02 | -0.56 to 0.52 | 0.394 | 90.9 | 54.82 | -1.92 to 1.88 |
| Non-septic controls | IL-8 | 4 | 0.47 | 0.12 to 0.82 | 0.089 | 74.7 | 11.86 | -1.03 to 1.97 |
| Non-septic controls | sTNFR-1 | 2 | 0.37 | -0.15 to 0.89 | 0.117 | 79.1 | 4.79 | not estimable (k = 2) |
| Non-septic controls | IL-10 | 7 | 0.16 | -0.09 to 0.41 | 0.067 | 65.5 | 17.41 | -0.58 to 0.91 |
| Septic controls | IL-6 | 12 | 1.05 | 0.51 to 1.59 | 0.798 | 91.8 | 134.32 | -1.03 to 3.14 |
| Septic controls | IFN-γ | 2 | 0.55 | 0.28 to 0.83 | 0.000 | 0.0 | 0.70 | not estimable (k = 2) |
| Septic controls | TNF-α | 4 | 0.58 | 0.08 to 1.08 | 0.154 | 61.3 | 7.75 | -1.43 to 2.59 |
| Septic controls | CRP | 2 | 0.25 | -1.33 to 1.82 | 1.008 | 77.8 | 4.51 | not estimable (k = 2) |
| Septic controls | IL-10 | 4 | 0.97 | 0.05 to 1.88 | 0.772 | 91.9 | 37.21 | -3.31 to 5.24 |
| Septic controls | S100B | 8 | 1.39 | 0.54 to 2.25 | 1.286 | 93.2 | 102.67 | -1.58 to 4.37 |
| Septic controls | NSE | 5 | 1.45 | 0.33 to 2.57 | 1.428 | 92.8 | 55.67 | -2.76 to 5.66 |
| Association (OR) | IFN-γ | 2 | 0.900 | 0.805 to 1.005 | 0.0000 | 0.0 | 0.02 | not estimable (k = 2) |
| Association (OR) | IL-1β | 2 | 1.020 | 1.007 to 1.032 | 0.0000 | 0.0 | 0.15 | not estimable (k = 2) |
| Association (OR) | IL-6 | 4 | 1.853 | 1.500 to 2.290 | 0.0156 | 34.2 | 4.56 | 0.910 to 3.773 |
| Association (OR) | S100B | 2 | 3.491 | 0.222 to 54.983 | 3.2007 | 79.0 | 4.76 | not estimable (k = 2) |

Study-level random-effects (DerSimonian-Laird) refit from the arm-level values printed in the forest panels. The 95% prediction interval is the pooled estimate $\pm t(k–2) \times \sqrt{(\tau^2 + SE^2)}$ and requires at least three studies. Estimates are standardized mean differences, or odds ratios for the association framework.

#### eTable 10. Risk-of-bias weight and GRADE decision for each comparison

| Framework | Biomarker | k | Low risk, n | Moderate, n | High, n | Weight not at low risk, % | Weight at high risk, % | GRADE risk-of-bias decision | Contributing studies |
|---|---|---|---|---|---|---|---|---|---|
| Non-septic controls | IL-17 | 3 | 1 | 1 | 1 | 61.2 | 24.0 | Serious - downgrade one level | Mao et al, 2023; Orhun et al, 2019; Smith et al, 2022 |
| Non-septic controls | IL-6 | 7 | 5 | 1 | 1 | 25.4 | 10.7 | Serious - downgrade one level | Mao et al, 2023; Orhun et al, 2019; Brummel et al, 2024; Jorge-Ripper et al, 2016; Lei et al, 2022; Ritter et al, 2014; Smith et al, 2022 |
| Non-septic controls | IL-1β | 5 | 3 | 1 | 1 | 26.9 | 9.7 | Serious - downgrade one level | Mao et al, 2023; Orhun et al, 2019; Brummel et al, 2024; Ritter et al, 2014; Smith et al, 2022 |
| Non-septic controls | IFN-γ | 4 | 2 | 1 | 1 | 34.5 | 12.6 | Serious - downgrade one level | Mao et al, 2023; Orhun et al, 2019; Brummel et al, 2024; Jorge-Ripper et al, 2016 |
| Non-septic controls | IL-12 | 4 | 2 | 1 | 1 | 42.3 | 18.2 | Serious - downgrade one level | Mao et al, 2023; Orhun et al, 2019; Brummel et al, 2024; Smith et al, 2022 |
| Non-septic controls | TNF-α | 6 | 4 | 1 | 1 | 32.3 | 15.4 | Serious - downgrade one level | Mao et al, 2023; Orhun et al, 2019; Brummel et al, 2024; Jorge-Ripper et al, 2016; Lei et al, 2022; Ritter et al, 2014 |
| Non-septic controls | IL-8 | 4 | 2 | 1 | 1 | 40.8 | 16.6 | Serious - downgrade one level | Mao et al, 2023; Orhun et al, 2019; Brummel et al, 2024; Smith et al, 2022 |
| Non-septic controls | sTNFR-1 | 2 | 2 | 0 | 0 | 0.0 | 0.0 | Not serious - no downgrade | Brummel et al, 2024; Ritter et al, 2014 |
| Non-septic controls | IL-10 | 7 | 5 | 1 | 1 | 24.9 | 10.1 | Not serious - no downgrade | Jorge-Ripper et al, 2016; Mao et al, 2023; Orhun et al, 2019; Brummel et al, 2024; Lei et al, 2022; Ritter et al, 2014; Smith et al, 2022 |
| Association (OR) | IFN-γ | 2 | 2 | 0 | 0 | 0.0 | 0.0 | Not serious - no downgrade | Brummel et al., 2024; Yu et al., 2025 |
| Association (OR) | IL-1β | 2 | 2 | 0 | 0 | 0.0 | 0.0 | Not serious - no downgrade | Brummel et al., 2024; Ritter et al., 2014 |
| Association (OR) | IL-6 | 4 | 4 | 0 | 0 | 0.0 | 0.0 | Not serious - no downgrade | Yu et al., 2025; Wei et al., 2025; Li et al., 2022; Brummel et al., 2024 |
| Association (OR) | S100B | 2 | 2 | 0 | 0 | 0.0 | 0.0 | Not serious - no downgrade | Erikson et al., 2018; Wu et al., 2020 |
| Septic controls | CRP | 2 | 2 | 0 | 0 | 0.0 | 0.0 | Not serious - no downgrade | Erikson et al., 2018; Pfister et al., 2008 |
| Septic controls | IFN-γ | 2 | 2 | 0 | 0 | 0.0 | 0.0 | Not serious - no downgrade | Jorge-Ripper et al., 2016; Yu et al., 2025 |
| Septic controls | IL-10 | 4 | 4 | 0 | 0 | 0.0 | 0.0 | Not serious - no downgrade | Jorge-Ripper et al., 2016; Lei et al., 2022; Li et al., 2022; Yu et al., 2025 |
| Septic controls | IL-6 | 12 | 10 | 1 | 1 | 15.4 | 8.7 | Not serious - no downgrade | Erikson et al., 2018; Jorge-Ripper et al., 2016; Lei et al., 2022; Feng et al., 2017; Guo et al., 2022; Li et al., 2022; Nakamura et al., 2003; Pfister et al., 2008; Wang et al., 2022; Wei et al., 2025; Wu et al., 2020; Yu et al., 2025 |
| Septic controls | NSE | 5 | 4 | 0 | 1 | 21.6 | 21.6 | Not serious - no downgrade | Erikson et al., 2018; Feng et al., 2017; Guo et al., 2022; Li et al., 2022; Wang et al., 2022 |
| Septic controls | S100B | 8 | 7 | 0 | 1 | 13.9 | 13.9 | Not serious - no downgrade | Erikson et al., 2018; Jorge-Ripper et al., 2016; Feng et al., 2017; Guo et al., 2022; Li et al., 2022; Pfister et al., 2008; Wang et al., 2022; Wu et al., 2020 |
| Septic controls | TNF-α | 4 | 4 | 0 | 0 | 0.0 | 0.0 | Not serious - no downgrade | Erikson et al., 2018; Jorge-Ripper et al., 2016; Lei et al., 2022; Li et al., 2022 |

Serious when more than 25% of the random-effects weight came from studies not at low risk; very serious when more than 50% came from studies at high risk.

#### eTable 11. Subgroup analysis by type of case definition

| Framework | Biomarker | Definition type | k | Estimate | 95% CI | τ² | I², % | Studies |
|---|---|---|---|---|---|---|---|---|
| Non-septic controls | IL-6 | CAM-ICU | 3 | 0.40 | -0.08 to 0.88 | 0.121 | 67.8 | Lei et al, 2022; Ritter et al, 2014; Smith et al, 2022 |
| Non-septic controls | IL-6 | composite | 1 | 0.24 | 0.11 to 0.38 | 0.000 | 0.0 | Brummel et al, 2024 |
| Non-septic controls | IL-6 | other scale | 1 | 0.83 | 0.33 to 1.32 | 0.000 | 0.0 | Jorge-Ripper et al, 2016 |
| Non-septic controls | IL-6 | unclear | 2 | 0.87 | 0.52 to 1.22 | 0.000 | 0.0 | Mao et al, 2023; Orhun et al, 2019 |
| Non-septic controls | IL-8 | CAM-ICU | 1 | 0.61 | 0.22 to 1.00 | 0.000 | 0.0 | Smith et al, 2022 |
| Non-septic controls | IL-8 | composite | 1 | 0.13 | -0.01 to 0.26 | 0.000 | 0.0 | Brummel et al, 2024 |
| Non-septic controls | IL-8 | unclear | 2 | 0.63 | 0.29 to 0.98 | 0.000 | 0.0 | Mao et al, 2023; Orhun et al, 2019 |
| Non-septic controls | IFN-γ | composite | 1 | 0.00 | -0.13 to 0.14 | 0.000 | 0.0 | Brummel et al, 2024 |
| Non-septic controls | IFN-γ | other scale | 1 | 0.55 | 0.07 to 1.03 | 0.000 | 0.0 | Jorge-Ripper et al, 2016 |
| Non-septic controls | IFN-γ | unclear | 2 | 0.17 | -0.17 to 0.50 | 0.000 | 0.0 | Mao et al, 2023; Orhun et al, 2019 |
| Non-septic controls | IL-12 | CAM-ICU | 1 | 0.39 | 0.01 to 0.77 | 0.000 | 0.0 | Smith et al, 2022 |
| Non-septic controls | IL-12 | composite | 1 | -0.07 | -0.21 to 0.06 | 0.000 | 0.0 | Brummel et al, 2024 |
| Non-septic controls | IL-12 | unclear | 2 | -0.34 | -1.07 to 0.38 | 0.206 | 74.8 | Mao et al, 2023; Orhun et al, 2019 |
| Non-septic controls | TNF-α | CAM-ICU | 2 | 0.40 | -0.37 to 1.18 | 0.241 | 76.5 | Lei et al, 2022; Ritter et al, 2014 |
| Non-septic controls | TNF-α | composite | 1 | -0.03 | -0.17 to 0.10 | 0.000 | 0.0 | Brummel et al, 2024 |
| Non-septic controls | TNF-α | other scale | 1 | 0.69 | 0.21 to 1.18 | 0.000 | 0.0 | Jorge-Ripper et al, 2016 |
| Non-septic controls | TNF-α | unclear | 2 | -0.79 | -2.11 to 0.54 | 0.842 | 92.0 | Mao et al, 2023; Orhun et al, 2019 |
| Non-septic controls | IL-1β | CAM-ICU | 2 | 0.47 | -0.10 to 1.04 | 0.123 | 72.1 | Ritter et al, 2014; Smith et al, 2022 |
| Non-septic controls | IL-1β | composite | 1 | 0.14 | 0.00 to 0.28 | 0.000 | 0.0 | Brummel et al, 2024 |
| Non-septic controls | IL-1β | unclear | 2 | 0.16 | -0.18 to 0.49 | 0.000 | 0.0 | Mao et al, 2023; Orhun et al, 2019 |
| Non-septic controls | IL-10 | CAM-ICU | 3 | 0.12 | -0.56 to 0.80 | 0.302 | 84.1 | Lei et al, 2022; Ritter et al, 2014; Smith et al, 2022 |
| Non-septic controls | IL-10 | composite | 1 | 0.02 | -0.12 to 0.16 | 0.000 | 0.0 | Brummel et al, 2024 |
| Non-septic controls | IL-10 | other scale | 1 | 0.34 | -0.13 to 0.82 | 0.000 | 0.0 | Jorge-Ripper et al, 2016 |
| Non-septic controls | IL-10 | unclear | 2 | 0.26 | -0.11 to 0.64 | 0.013 | 15.5 | Mao et al, 2023; Orhun et al, 2019 |
| Non-septic controls | IL-17 | CAM-ICU | 1 | 0.67 | 0.29 to 1.06 | 0.000 | 0.0 | Smith et al, 2022 |
| Non-septic controls | IL-17 | unclear | 2 | 0.20 | -0.14 to 0.53 | 0.000 | 0.0 | Mao et al, 2023; Orhun et al, 2019 |
| Non-septic controls | sTNFR-1 | CAM-ICU | 1 | 0.69 | 0.22 to 1.16 | 0.000 | 0.0 | Ritter et al, 2014 |
| Non-septic controls | sTNFR-1 | composite | 1 | 0.15 | 0.01 to 0.28 | 0.000 | 0.0 | Brummel et al, 2024 |
| Non-septic controls | CRP | composite | 1 | 0.13 | -0.01 to 0.27 | 0.000 | 0.0 | Brummel et al, 2024 |
| Association (OR) | IFN-γ | composite | 1 | -0.105 | -0.217 to 0.006 | 0.0000 | 0.0 | Brummel et al., 2024 |
| Association (OR) | IFN-γ | unclear | 1 | -0.208 | -1.828 to 1.411 | 0.0000 | 0.0 | Yu et al., 2025 |
| Association (OR) | IL-1β | CAM-ICU | 1 | 0.020 | 0.008 to 0.032 | 0.0000 | 0.0 | Ritter et al., 2014 |
| Association (OR) | IL-1β | composite | 1 | 0.000 | -0.100 to 0.100 | 0.0000 | 0.0 | Brummel et al., 2024 |
| Association (OR) | IL-6 | composite | 1 | 0.588 | 0.374 to 0.802 | 0.0000 | 0.0 | Brummel et al., 2024 |
| Association (OR) | IL-6 | unclear | 3 | 0.685 | 0.288 to 1.081 | 0.0649 | 56.0 | Yu et al., 2025; Wei et al., 2025; Li et al., 2022 |
| Association (OR) | S100B | CAM-ICU | 1 | 2.890 | 0.516 to 5.265 | 0.0000 | 0.0 | Erikson et al., 2018 |
| Association (OR) | S100B | composite | 1 | 0.044 | -0.901 to 0.989 | 0.0000 | 0.0 | Wu et al., 2020 |
| Septic controls | CRP | CAM-ICU | 2 | 0.246 | -1.327 to 1.819 | 1.0081 | 77.8 | Erikson et al., 2018; Pfister et al., 2008 |
| Septic controls | IFN-γ | other scale | 1 | 2.759 | 2.143 to 3.374 | 0.0000 | 0.0 | Jorge-Ripper et al., 2016 |
| Septic controls | IFN-γ | unclear | 1 | 0.644 | 0.302 to 0.987 | 0.0000 | 0.0 | Yu et al., 2025 |
| Septic controls | IL-10 | CAM-ICU | 1 | 0.015 | -0.564 to 0.595 | 0.0000 | 0.0 | Lei et al., 2022 |
| Septic controls | IL-10 | other scale | 1 | 1.408 | 0.918 to 1.898 | 0.0000 | 0.0 | Jorge-Ripper et al., 2016 |

| Framework | Biomarker | Definition type | k | Estimate | 95% CI | τ² | I², % | Studies |
|---|---|---|---|---|---|---|---|---|
| Septic controls | IL-10 | unclear | 2 | 1.975 | -0.445 to 4.396 | 2.9179 | 95.6 | Li et al., 2022; Yu et al., 2025 |
| Septic controls | IL-6 | CAM-ICU | 3 | 0.133 | -0.471 to 0.736 | 0.1103 | 37.8 | Erikson et al., 2018; Lei et al., 2022; Pfister et al., 2008 |
| Septic controls | IL-6 | composite | 1 | 0.607 | 0.210 to 1.004 | 0.0000 | 0.0 | Wu et al., 2020 |
| Septic controls | IL-6 | other scale | 1 | -0.032 | -0.470 to 0.406 | 0.0000 | 0.0 | Jorge-Ripper et al., 2016 |
| Septic controls | IL-6 | unclear | 7 | 1.592 | 0.972 to 2.213 | 0.5953 | 89.8 | Feng et al., 2017; Guo et al., 2022; Li et al., 2022; Nakamura et al., 2003; Wang et al., 2022; Wei et al., 2025; Yu et al., 2025 |
| Septic controls | NSE | CAM-ICU | 1 | -0.057 | -0.897 to 0.782 | 0.0000 | 0.0 | Erikson et al., 2018 |
| Septic controls | NSE | unclear | 4 | 1.864 | 0.538 to 3.190 | 1.6303 | 94.2 | Feng et al., 2017; Guo et al., 2022; Li et al., 2022; Wang et al., 2022 |
| Septic controls | S100B | CAM-ICU | 2 | 0.737 | 0.038 to 1.437 | 0.0000 | 0.0 | Erikson et al., 2018; Pfister et al., 2008 |
| Septic controls | S100B | composite | 1 | 0.878 | 0.472 to 1.285 | 0.0000 | 0.0 | Wu et al., 2020 |
| Septic controls | S100B | other scale | 1 | -0.042 | -0.479 to 0.396 | 0.0000 | 0.0 | Jorge-Ripper et al., 2016 |
| Septic controls | S100B | unclear | 4 | 2.826 | 1.006 to 4.647 | 3.0527 | 96.4 | Feng et al., 2017; Guo et al., 2022; Li et al., 2022; Wang et al., 2022 |
| Septic controls | TNF-α | CAM-ICU | 2 | 0.461 | -0.022 to 0.945 | 0.0000 | 0.0 | Erikson et al., 2018; Lei et al., 2022 |
| Septic controls | TNF-α | other scale | 1 | 1.329 | 0.844 to 1.813 | 0.0000 | 0.0 | Jorge-Ripper et al., 2016 |
| Septic controls | TNF-α | unclear | 1 | 1.351 | 0.666 to 2.036 | 0.0000 | 0.0 | Li et al., 2022 |

Definition types as coded in eTable 8. No comparison has two instrument-defined strata with two or more studies each, so no interaction test by definition type is estimable.

#### eTable 12. Pooled effect by whether an ascertainment instrument is stated

| Framework | Biomarker | Subgroup | k | Estimate | 95% CI | τ² | I², % | Q for interaction | p for interaction | Studies |
|---|---|---|---|---|---|---|---|---|---|---|
| Non-septic controls | IL-6 | explicit instrument | 5 | 0.43 | 0.13 to 0.73 | 0.074 | 68.7 | 3.40 | 0.0651 | Brummel et al, 2024; Jorge-Ripper et al, 2016; Lei et al, 2022; Ritter et al, 2014; Smith et al, 2022 |
| Non-septic controls | IL-6 | no stated instrument | 2 | 0.87 | 0.52 to 1.22 | 0.000 | 0.0 | 3.40 | 0.0651 | Mao et al, 2023; Orhun et al, 2019 |
| Non-septic controls | IL-8 | explicit instrument | 2 | 0.33 | -0.13 to 0.80 | 0.094 | 81.1 | 1.03 | 0.3091 | Brummel et al, 2024; Smith et al, 2022 |
| Non-septic controls | IL-8 | no stated instrument | 2 | 0.63 | 0.29 to 0.98 | 0.000 | 0.0 | 1.03 | 0.3091 | Mao et al, 2023; Orhun et al, 2019 |
| Non-septic controls | IFN-γ | explicit instrument | 2 | 0.23 | -0.30 to 0.75 | 0.117 | 78.2 | 0.03 | 0.8519 | Brummel et al, 2024; Jorge-Ripper et al, 2016 |
| Non-septic controls | IFN-γ | no stated instrument | 2 | 0.17 | -0.17 to 0.50 | 0.000 | 0.0 | 0.03 | 0.8519 | Mao et al, 2023; Orhun et al, 2019 |
| Non-septic controls | IL-12 | explicit instrument | 2 | 0.12 | -0.33 to 0.57 | 0.085 | 80.0 | 1.14 | 0.2853 | Brummel et al, 2024; Smith et al, 2022 |
| Non-septic controls | IL-12 | no stated instrument | 2 | -0.34 | -1.07 to 0.38 | 0.206 | 74.8 | 1.14 | 0.2853 | Mao et al, 2023; Orhun et al, 2019 |
| Non-septic controls | TNF-α | explicit instrument | 4 | 0.32 | -0.10 to 0.75 | 0.143 | 79.2 | 2.44 | 0.1180 | Brummel et al, 2024; Jorge-Ripper et al, 2016; Lei et al, 2022; Ritter et al, 2014 |
| Non-septic controls | TNF-α | no stated instrument | 2 | -0.79 | -2.11 to 0.54 | 0.842 | 92.0 | 2.44 | 0.1180 | Mao et al, 2023; Orhun et al, 2019 |
| Non-septic controls | IL-1β | explicit instrument | 3 | 0.31 | -0.02 to 0.65 | 0.059 | 69.3 | 0.43 | 0.5107 | Brummel et al, 2024; Ritter et al, 2014; Smith et al, 2022 |
| Non-septic controls | IL-1β | no stated instrument | 2 | 0.16 | -0.18 to 0.49 | 0.000 | 0.0 | 0.43 | 0.5107 | Mao et al, 2023; Orhun et al, 2019 |
| Non-septic controls | IL-10 | explicit instrument | 5 | 0.14 | -0.18 to 0.46 | 0.090 | 73.2 | 0.25 | 0.6152 | Jorge-Ripper et al, 2016; Brummel et al, 2024; Lei et al, 2022; Ritter et al, 2014; Smith et al, 2022 |
| Non-septic controls | IL-10 | no stated instrument | 2 | 0.26 | -0.11 to 0.64 | 0.013 | 15.5 | 0.25 | 0.6152 | Mao et al, 2023; Orhun et al, 2019 |
| Non-septic controls | IL-17 | explicit instrument | 1 | 0.67 | 0.29 to 1.06 | 0.000 | 0.0 | | | Smith et al, 2022 |
| Non-septic controls | IL-17 | no stated instrument | 2 | 0.20 | -0.14 to 0.53 | 0.000 | 0.0 | | | Mao et al, 2023; Orhun et al, 2019 |
| Non-septic controls | sTNFR-1 | explicit instrument | 2 | 0.37 | -0.15 to 0.89 | 0.117 | 79.1 | | | Brummel et al, 2024; Ritter et al, 2014 |
| Non-septic controls | CRP | explicit instrument | 1 | 0.13 | -0.01 to 0.27 | 0.000 | 0.0 | | | Brummel et al, 2024 |
| Association (OR) | IFN-γ | explicit instrument | 1 | -0.105 | -0.217 to 0.006 | 0.0 | 0.0 | | | Brummel et al., 2024 |
| Association (OR) | IFN-γ | no stated instrument | 1 | -0.208 | -1.828 to 1.411 | 0.0 | 0.0 | | | Yu et al., 2025 |
| Association (OR) | IL-1β | explicit instrument | 2 | 0.02 | 0.007 to 0.032 | 0.0 | 0.0 | | | Brummel et al., 2024; Ritter et al., 2014 |
| Association (OR) | IL-6 | no stated instrument | 3 | 0.685 | 0.288 to 1.081 | 0.0649 | 56.0 | | | Yu et al., 2025; Wei et al., 2025; Li et al., 2022 |
| Association (OR) | IL-6 | explicit instrument | 1 | 0.588 | 0.374 to 0.802 | 0.0 | 0.0 | | | Brummel et al., 2024 |
| Association (OR) | S100B | explicit instrument | 2 | 1.25 | -1.507 to 4.007 | 3.2007 | 79.0 | | | Erikson et al., 2018; Wu et al., 2020 |
| Septic controls | CRP | explicit instrument | 2 | 0.246 | -1.327 to 1.819 | 1.0081 | 77.8 | | | Erikson et al., 2018; Pfister et al., 2008 |
| Septic controls | IFN-γ | explicit instrument | 1 | 2.759 | 2.143 to 3.374 | 0.0 | 0.0 | | | Jorge-Ripper et al., 2016 |
| Septic controls | IFN-γ | no stated instrument | 1 | 0.644 | 0.302 to 0.987 | 0.0 | 0.0 | | | Yu et al., 2025 |
| Septic controls | IL-10 | explicit instrument | 2 | 0.721 | -0.644 to 2.085 | 0.8946 | 92.3 | 0.78 | 0.3761 | Jorge-Ripper et al., 2016; Lei et al., 2022 |
| Septic controls | IL-10 | no stated instrument | 2 | 1.975 | -0.445 to 4.396 | 2.9179 | 95.6 | 0.78 | 0.3761 | Li et al., 2022; Yu et al., 2025 |
| Septic controls | IL-6 | explicit instrument | 5 | 0.222 | -0.175 to 0.62 | 0.1032 | 54.1 | 13.28 | 0.0003 | Erikson et al., 2018; Jorge-Ripper et al., 2016; Lei et al., 2022; Pfister et al., 2008; Wu et al., 2020 |

| Framework | Biomarker | Subgroup | k | Estimate | 95% CI | τ² | I², % | Q for interaction | p for interaction | Studies |
|---|---|---|---|---|---|---|---|---|---|---|
| Septic controls | IL-6 | no stated instrument | 7 | 1.592 | 0.972 to 2.213 | 0.5953 | 89.8 | 13.28 | 0.0003 | Feng et al., 2017; Guo et al., 2022; Li et al., 2022; Nakamura et al., 2003; Wang et al., 2022; Wei et al., 2025; Yu et al., 2025 |
| Septic controls | NSE | explicit instrument | 1 | -0.057 | -0.897 to 0.782 | 0.0 | 0.0 | | | Erikson et al., 2018 |
| Septic controls | NSE | no stated instrument | 4 | 1.864 | 0.538 to 3.19 | 1.6303 | 94.2 | | | Feng et al., 2017; Guo et al., 2022; Li et al., 2022; Wang et al., 2022 |
| Septic controls | S100B | explicit instrument | 4 | 0.532 | -0.032 to 1.095 | 0.208 | 69.0 | 5.57 | 0.0183 | Erikson et al., 2018; Jorge-Ripper et al., 2016; Pfister et al., 2008; Wu et al., 2020 |
| Septic controls | S100B | no stated instrument | 4 | 2.826 | 1.006 to 4.647 | 3.0527 | 96.4 | 5.57 | 0.0183 | Feng et al., 2017; Guo et al., 2022; Li et al., 2022; Wang et al., 2022 |
| Septic controls | TNF-α | explicit instrument | 3 | 0.788 | 0.151 to 1.424 | 0.2117 | 67.9 | | | Erikson et al., 2018; Jorge-Ripper et al., 2016; Lei et al., 2022 |
| Septic controls | TNF-α | no stated instrument | 1 | 1.351 | 0.666 to 2.036 | 0.0 | 0.0 | | | Li et al., 2022 |

Interaction tested where both subgroups contain at least two studies. Descriptive contrast of reporting quality, not a test of a biological hypothesis (eFigure 4).

#### eTable 13. Verification of arm-level values against the source publications

| Framework | Biomarker | Study | Finding on verification against the source publication | Value used in this report | Pooled SMD (95% CI) in this report |
|---|---|---|---|---|---|
| Non-septic controls | CRP | Smith 2022 | No CRP concentration is reported in the source (Tables 1 and 4); the plotted arms reproduced the IL-1β arms | Row not included; CRP versus non-septic controls rests on one study and is not pooled | Not pooled (k = 1) |
| Non-septic controls | IL-1β | Brummel 2024 | Values digitized from Figure 1 (natural-log scale); the index arm used absolute values of negative logarithms and the control arm day-3 quartiles | Signed day-1 values for both arms | 0.24 (0.03 to 0.45) |
| Non-septic controls | sTNFR-1 | Ritter 2014 | Plotted arms did not correspond to the sTNFR-1 row of Table 2 of the source | Delirium 3,843 (3,072–6,003) vs no delirium 2,419 (1,658–4,440) pg/mL | 0.37 (-0.15 to 0.89) |
| Non-septic controls | IL-10 | Ritter 2014 | Control interquartile range entered as 0–6.9; the source prints 0 to 69 | 0 to 69, as printed | 0.16 (-0.09 to 0.41) |
| Non-septic controls | IL-1β, IL-6, IL-8, IL-10, IL-12, IFN-γ, TNF-α, IL-17 | Orhun 2019 | Patient arm taken from Table 2 of the source (delirium n = 64 plus coma n = 18), which reports mean ± SE; SE had been entered as SD. Control arm digitized from Figure 3 | Patient arm combined across delirium and coma with SE converted to SD ($SD = SE \times \sqrt{n}$); control arm as digitized | See eTable 22 |
| Septic controls | NSE | Li 2022 | SAE arm size entered as 20; the source reports 21 | n = 21 | 1.45 (0.33 to 2.57) |
| Non-septic controls | IL-17 | Smith 2022 | Control group reported as 0 (0–0) | An SD of 0.074 was imputed to allow estimation | 0.37 (0.01 to 0.73) |
| Non-septic controls | IL-1β, IL-6, IL-10, IL-12, IFN-γ, TNF-α, IL-17 | Mao 2023 | Cerebrospinal fluid concentrations reported as mean ± SEM (IL-6 digitized from a figure); SEM had been entered as SD | SEM converted to SD ($SD = SEM \times \sqrt{n}$); compartment declared and examined in eTable 26 | See eTable 22 |
| Septic controls | IL-6, IL-10, TNF-α, IFN-γ, S100B | Jorge-Ripper 2016 | Table 4 of the source reports mean ± SEM; SEM had been entered as SD | SEM converted to SD; delirium with sepsis (n = 47) vs sepsis without delirium (n = 35) | See eTable 22 |
| Non-septic controls | IL-6, IL-10, TNF-α, IFN-γ | Jorge-Ripper 2016 | Index arm had been delirium without sepsis (n = 35); SEM entered as SD | Index arm delirium with sepsis (n = 47) vs healthy controls (n = 27); SEM converted to SD | See eTable 22 |

Every arm value was traced to the source publication and to the extraction workbook. The pooled estimates reported throughout this document and the article incorporate the values in the fifth column. SE = standard error.

#### eTable 14. Completeness sensitivity analysis

| Framework | Biomarker | Study added by the audit | Why missed at extraction | Added SMD (95% CI) | k without | Pooled without (95% CI) | $I^2$ without | k with | Pooled with (95% CI) | $I^2$ with | Changes significance |
|---|---|---|---|---|---|---|---|---|---|---|---|
| Non-septic controls | IL-1β | Smith 2022 | Row present and complete in the workbook | 0.19 (-0.18 to 0.57) | 4 | 0.27 (-0.02 to 0.55) | 57% | 5 | 0.24 (0.03 to 0.45) | 43% | yes |
| Non-septic controls | IL-8 | Mao 2023 | Median recorded inside the IQR cell, so the median column was empty | 0.86 (0.23 to 1.49) | 3 | 0.38 (0.03 to 0.74) | 75% | 4 | 0.47 (0.12 to 0.82) | 75% | no |

The two eligible comparisons identified by the completeness audit and the effect of including them. Both are included in every analysis reported.

#### eTable 15. Audit of reported participant totals against arm-level totals

| Biomarker | Studies in evidence map | n, extraction | Comparisons pooled | Studies pooled | n, arm-level pooled | Gap (extraction − pooled) |
|---|---|---|---|---|---|---|
| IL-6 | 18 | 2416 | 19 | 17 | 2393 | 23 |
| IL-17 | 4 | 184 | 3 | 3 | 277 | -93 |
| TNF-α | 8 | 1975 | 10 | 8 | 1550 | 425 |
| IL-1β | 5 | 1264 | 5 | 5 | 1346 | -82 |
| IL-8 | 4 | 1239 | 4 | 4 | 1268 | -29 |
| IL-10 | 9 | 1583 | 11 | 9 | 1787 | -204 |
| IFN-γ | 5 | 1412 | 6 | 5 | 1445 | -33 |
| S100B | 8 | 873 | 8 | 8 | 484 | 389 |
| NSE | 5 | 379 | 5 | 5 | 282 | 97 |
| CRP | 4 | 1132 | 2 | 2 | 38 | 1094 |
| sTNFR-1 | 2 | 1416 | 2 | 2 | 1069 | 347 |
| IL-12 | 4 | 1244 | 4 | 4 | 1268 | -24 |

The extraction total counts every participant of every study that measured the biomarker; the arm-level total counts the arms entering pooled comparisons in either contrast framework (a study entering both frameworks is counted in each). Gaps reflect studies reporting the biomarker in a form that could not be pooled, or arms reused across frameworks.

#### eTable 16. Review Manager entry sheet: arm-level values by panel and subgroup

| Framework | Panel | Subgroup | Study | Index mean | Index SD | Index n | Control mean | Control SD | Control n | SMD (95% CI) | Weight, % |
|---|---|---|---|---|---|---|---|---|---|---|---|
| Non-septic controls | IL-6 | SAE | Mao 2023 | 9.67 | 14.21 | 19 | 3.15 | 2.64545 | 24 | 0.67 (0.05 to 1.28) | 10.7 |
| Non-septic controls | IL-6 | SAE | Orhun 2019 | 54.22 | 66.32 | 82 | 0 | 0.02 | 33 | 0.96 (0.54 to 1.38) | 14.7 |
| Non-septic controls | IL-6 | Delirium only | Brummel 2024 | 3.56 | 1.6 | 691 | 3.166 | 1.68 | 300 | 0.24 (0.11 to 0.38) | 20.8 |
| Non-septic controls | IL-6 | Delirium only | Jorge-Ripper 2016 | 31.5 | 42.5051 | 47 | 2.9 | 6.755 | 27 | 0.83 (0.33 to 1.32) | 13.2 |
| Non-septic controls | IL-6 | Delirium only | Lei 2022 | 43.17 | 51.98 | 17 | 50.13 | 74.4 | 35 | -0.10 (-0.68 to 0.48) | 11.4 |
| Non-septic controls | IL-6 | Delirium only | Ritter 2014 | 332.33 | 97.77 | 31 | 237.33 | 288.88 | 47 | 0.40 (-0.05 to 0.86) | 13.9 |
| Non-septic controls | IL-6 | Delirium only | Smith 2022 | 426.84 | 418.61 | 41 | 181.89 | 238.74 | 78 | 0.78 (0.39 to 1.17) | 15.4 |
| Non-septic controls | IL-8 | SAE | Mao 2023 | 203.91 | 290.33 | 19 | 34.22 | 22.65 | 24 | 0.86 (0.23 to 1.49) | 16.6 |
| Non-septic controls | IL-8 | SAE | Orhun 2019 | 152.81 | 333.83 | 82 | 0 | 0.05 | 33 | 0.54 (0.13 to 0.95) | 24.1 |
| Non-septic controls | IL-8 | Delirium only | Brummel 2024 | 1.93667 | 0.94074 | 691 | 1.81333 | 1.00741 | 300 | 0.13 (-0.01 to 0.26) | 34.1 |
| Non-septic controls | IL-8 | Delirium only | Smith 2022 | 53.35 | 50.6 | 41 | 32.2967 | 21.5333 | 78 | 0.61 (0.22 to 1.00) | 25.1 |
| Non-septic controls | IFN-γ | SAE | Mao 2023 | 3.24 | 9.76393 | 19 | 3.05 | 7.83837 | 24 | 0.02 (-0.58 to 0.62) | 12.6 |
| Non-septic controls | IFN-γ | SAE | Orhun 2019 | 7.25 | 18.02 | 82 | 3.65 | 0.91 | 33 | 0.23 (-0.17 to 0.64) | 21.9 |
| Non-septic controls | IFN-γ | Delirium only | Brummel 2024 | 0.07667 | 1.34074 | 691 | 0.07 | 1.40741 | 300 | 0.00 (-0.13 to 0.14) | 48.1 |
| Non-septic controls | IFN-γ | Delirium only | Jorge-Ripper 2016 | 22.8 | 43.1906 | 47 | 3.45 | 5.97558 | 27 | 0.55 (0.07 to 1.03) | 17.4 |
| Non-septic controls | IL-12 | SAE | Mao 2023 | 2.6 | 0.741013 | 19 | 2.56 | 0.538888 | 24 | 0.06 (-0.54 to 0.66) | 18.2 |
| Non-septic controls | IL-12 | SAE | Orhun 2019 | 10.75 | 9.77 | 82 | 16.62 | 4.155 | 33 | -0.68 (-1.09 to -0.27) | 24.1 |
| Non-septic controls | IL-12 | Delirium only | Brummel 2024 | 0.15 | 1.5037 | 691 | 0.26333 | 1.59259 | 300 | -0.07 (-0.21 to 0.06) | 32.5 |
| Non-septic controls | IL-12 | Delirium only | Smith 2022 | 0.13667 | 0.3037 | 41 | 0.05667 | 0.12593 | 78 | 0.39 (0.01 to 0.77) | 25.2 |
| Non-septic controls | TNF-α | SAE | Mao 2023 | 2.6 | 0.697424 | 19 | 2.68 | 0.930806 | 24 | -0.09 (-0.70 to 0.51) | 15.4 |
| Non-septic controls | TNF-α | SAE | Orhun 2019 | 37.92 | 51.62 | 82 | 105.04 | 27.5 | 33 | -1.45 (-1.89 to -1.00) | 16.9 |
| Non-septic controls | TNF-α | Delirium only | Brummel 2024 | 2.09333 | 0.71852 | 691 | 2.11667 | 0.77037 | 300 | -0.03 (-0.17 to 0.10) | 18.9 |
| Non-septic controls | TNF-α | Delirium only | Jorge-Ripper 2016 | 18 | 28.1082 | 47 | 2.17 | 4.15692 | 27 | 0.69 (0.21 to 1.18) | 16.5 |
| Non-septic controls | TNF-α | Delirium only | Lei 2022 | 14.17 | 6 | 17 | 10.89 | 2.372 | 35 | 0.82 (0.22 to 1.43) | 15.4 |
| Non-septic controls | TNF-α | Delirium only | Ritter 2014 | 225.33 | 382.96 | 31 | 214.67 | 298.52 | 47 | 0.03 (-0.42 to 0.49) | 16.8 |
| Non-septic controls | IL-1β | SAE | Mao 2023 | 2.9 | 3.22559 | 19 | 3 | 5.73181 | 24 | -0.02 (-0.62 to 0.58) | 9.7 |
| Non-septic controls | IL-1β | SAE | Orhun 2019 | 15.63 | 78.09 | 82 | 0 | 0.1 | 33 | 0.23 (-0.17 to 0.64) | 17.2 |
| Non-septic controls | IL-1β | Delirium only | Brummel 2024 | -0.85 | 1.05926 | 691 | -0.98667 | 0.75556 | 300 | 0.14 (0.00 to 0.28) | 40.3 |
| Non-septic controls | IL-1β | Delirium only | Ritter 2014 | 76.333 | 82.222 | 31 | 34.333 | 17.778 | 47 | 0.78 (0.31 to 1.25) | 14.1 |
| Non-septic controls | IL-1β | Delirium only | Smith 2022 | 9.813 | 11.651 | 41 | 7.403 | 12.674 | 78 | 0.19 (-0.18 to 0.57) | 18.7 |
| Non-septic controls | IL-10 | Delirium only | Jorge-Ripper 2016 | 9.9 | 35.6494 | 47 | 0 | 0 | 27 | 0.34 (-0.13 to 0.82) | 12.9 |
| Non-septic controls | IL-10 | SAE | Mao 2023 | 2.57 | 0.348712 | 19 | 2.57 | 0.293939 | 24 | 0.00 (-0.60 to 0.60) | 10.1 |
| Non-septic controls | IL-10 | SAE | Orhun 2019 | 12.1 | 35.2 | 82 | 0 | 0.01 | 33 | 0.40 (-0.00 to 0.81) | 14.8 |
| Non-septic controls | IL-10 | Delirium only | Brummel 2024 | 2.25667 | 1.25926 | 691 | 2.23 | 1.41481 | 300 | 0.02 (-0.12 to 0.16) | 22.7 |
| Non-septic controls | IL-10 | Delirium only | Lei 2022 | 23.97 | 27.8 | 17 | 18.67 | 23.29 | 35 | 0.21 (-0.37 to 0.79) | 10.6 |
| Non-septic controls | IL-10 | Delirium only | Ritter 2014 | 5.76667 | 1.11111 | 31 | 24.7667 | 51.1111 | 47 | -0.47 (-0.93 to -0.01) | 13.4 |
| Non-septic controls | IL-10 | Delirium only | Smith 2022 | 50.79 | 63.459 | 41 | 21.99 | 35.0148 | 78 | 0.61 (0.23 to 1.00) | 15.4 |
| Non-septic controls | IL-17 | SAE | Mao 2023 | 12 | 13.0767 | 19 | 11.87 | 13.0313 | 24 | 0.01 (-0.59 to 0.61) | 24.0 |
| Non-septic controls | IL-17 | SAE | Orhun 2019 | 5.03 | 18.33 | 82 | 0.62 | 0.155 | 33 | 0.28 (-0.12 to 0.69) | 37.3 |
| Non-septic controls | IL-17 | Delirium only | Smith 2022[a] | 0.31 | 0.68889 | 41 | 0.03333 | 0.07407 | 78 | 0.67 (0.29 to 1.06) | 38.8 |
| Non-septic controls | sTNFR-1 | Delirium only | Brummel 2024 | 8.993 | 1.044 | 691 | 8.843 | 0.955 | 300 | 0.15 (0.01 to 0.28) | 58.8 |
| Non-septic controls | sTNFR-1 | Delirium only | Ritter 2014 | 4306 | 2171.1 | 31 | 2839 | 2060.7 | 47 | 0.69 (0.22 to 1.16) | 41.2 |

| Framework | Panel | Subgroup | Study | Index mean | Index SD | Index n | Control mean | Control SD | Control n | SMD (95% CI) | Weight, % |
|---|---|---|---|---|---|---|---|---|---|---|---|
| Non-septic controls | CRP | Delirium only | Brummel 2024 | 11.66 | 0.71852 | 691 | 11.5533 | 1.01481 | 300 | 0.13 (-0.01 to 0.27) | 100.0 |
| Septic controls | CRP | SAE | Pfister 2008 | 318 | 162.963 | 12 | 130 | 148.148 | 4 | 1.11 (-0.10 to 2.33) | 46.2 |
| Septic controls | CRP | Delirium only | Erikson 2018 | 185.333 | 88.148 | 10 | 236 | 105.185 | 12 | -0.50 (-1.35 to 0.36) | 53.8 |
| Septic controls | IFN-γ | SAE | Yu 2025 | 106.967 | 127.652 | 62 | 46.91 | 49.896 | 78 | 0.64 (0.30 to 0.99) | 62.5 |
| Septic controls | IFN-γ | Delirium only | Jorge-Ripper 2016 | 22.8 | 43.1906 | 47 | 9.1 | 10.6489 | 35 | 0.41 (-0.04 to 0.85) | 37.5 |
| Septic controls | IL-6 | SAE | Feng 2017 | 1919.2 | 3643.45 | 36 | 299.857 | 448.837 | 23 | 0.56 (0.02 to 1.09) | 8.8 |
| Septic controls | IL-6 | SAE | Guo 2022 | 229.15 | 29.61 | 30 | 196.71 | 18.62 | 90 | 1.48 (1.02 to 1.93) | 9.0 |
| Septic controls | IL-6 | SAE | Li 2022 | 237 | 16 | 21 | 191 | 18 | 20 | 2.65 (1.79 to 3.51) | 7.7 |
| Septic controls | IL-6 | SAE | Nakamura 2003 | 6050 | 1100 | 16 | 2750 | 1150 | 10 | 2.86 (1.70 to 4.01) | 6.7 |
| Septic controls | IL-6 | SAE | Pfister 2008 | 409.63 | 599.18 | 12 | 79.9 | 51.62 | 4 | 0.59 (-0.57 to 1.74) | 6.7 |
| Septic controls | IL-6 | SAE | Wang 2022 | 1999.33 | 3545.18 | 20 | 306.667 | 392.593 | 20 | 0.66 (0.02 to 1.30) | 8.4 |
| Septic controls | IL-6 | SAE | Wei 2025 | 928.71 | 163.74 | 105 | 528.71 | 176.51 | 114 | 2.34 (1.99 to 2.68) | 9.2 |
| Septic controls | IL-6 | SAE | Wu 2020 | 1876 | 3634.59 | 59 | 197.867 | 236.444 | 45 | 0.61 (0.21 to 1.00) | 9.1 |
| Septic controls | IL-6 | SAE | Yu 2025 | 13370.8 | 17953.1 | 62 | 423.027 | 716.133 | 78 | 1.08 (0.72 to 1.43) | 9.2 |
| Septic controls | IL-6 | Delirium only | Erikson 2018 | 250.86 | 293.11 | 10 | 126.1 | 199.03 | 12 | 0.49 (-0.37 to 1.34) | 7.7 |
| Septic controls | IL-6 | Delirium only | Jorge-Ripper 2016 | 31.5 | 42.5051 | 47 | 31.7 | 36.6797 | 35 | -0.00 (-0.44 to 0.43) | 9.0 |
| Septic controls | IL-6 | Delirium only | Lei 2022 | 43.17 | 51.98 | 17 | 63.83 | 74.46 | 35 | -0.30 (-0.88 to 0.28) | 8.6 |
| Septic controls | IL-10 | SAE | Li 2022 | 30.3 | 1.8 | 21 | 25.2 | 1.2 | 20 | 3.25 (2.29 to 4.21) | 21.4 |
| Septic controls | IL-10 | SAE | Yu 2025 | 1343.68 | 2337.6 | 62 | 121.443 | 26.296 | 78 | 0.78 (0.44 to 1.13) | 27.0 |
| Septic controls | IL-10 | Delirium only | Jorge-Ripper 2016 | 9.9 | 35.6494 | 47 | 3.8 | 15.3818 | 35 | 0.21 (-0.23 to 0.65) | 26.4 |
| Septic controls | IL-10 | Delirium only | Lei 2022 | 23.97 | 27.8 | 17 | 23.57 | 24.48 | 35 | 0.02 (-0.56 to 0.59) | 25.2 |
| Septic controls | NSE | SAE | Feng 2017 | 20.933 | 12.978 | 36 | 15.4 | 10.689 | 23 | 0.45 (-0.08 to 0.98) | 21.6 |
| Septic controls | NSE | SAE | Guo 2022 | 10.16 | 2.11 | 30 | 8.62 | 1.62 | 90 | 0.87 (0.44 to 1.30) | 22.0 |
| Septic controls | NSE | SAE | Li 2022 | 18.4 | 2.2 | 21 | 6.3 | 1.1 | 20 | 6.77 (5.11 to 8.43) | 15.1 |
| Septic controls | NSE | SAE | Wang 2022 | 13.33 | 5.36 | 20 | 10.23 | 2.98 | 20 | 0.70 (0.06 to 1.34) | 21.1 |
| Septic controls | NSE | Delirium only | Erikson 2018 | 21.4 | 10.963 | 10 | 22.2 | 15.185 | 12 | -0.06 (-0.90 to 0.78) | 20.1 |
| Septic controls | S100B | SAE | Feng 2017 | 0.34 | 0.28889 | 36 | 0.30667 | 0.27407 | 23 | 0.12 (-0.41 to 0.64) | 13.9 |
| Septic controls | S100B | SAE | Guo 2022 | 0.27 | 0.06 | 30 | 0.18 | 0.04 | 90 | 1.96 (1.47 to 2.44) | 14.0 |
| Septic controls | S100B | SAE | Li 2022 | 2.38 | 0.21 | 21 | 0.35 | 0.12 | 20 | 11.56 (8.86 to 14.27) | 5.9 |
| Septic controls | S100B | SAE | Pfister 2008 | 0.276 | 0.318 | 12 | 0.076 | 0.037 | 4 | 0.67 (-0.49 to 1.83) | 11.5 |
| Septic controls | S100B | SAE | Wang 2022 | 892.85 | 383.93 | 20 | 625 | 178.57 | 20 | 0.88 (0.22 to 1.53) | 13.5 |
| Septic controls | S100B | SAE | Wu 2020 | 0.311 | 0.219 | 59 | 0.155 | 0.094 | 45 | 0.88 (0.47 to 1.28) | 14.2 |
| Septic controls | S100B | Delirium only | Erikson 2018 | 0.36 | 0.29 | 10 | 0.173 | 0.17 | 12 | 0.78 (-0.10 to 1.65) | 12.7 |
| Septic controls | S100B | Delirium only | Jorge-Ripper 2016 | 26.3 | 52.103 | 47 | 26.6 | 38.4545 | 35 | -0.01 (-0.44 to 0.43) | 14.1 |
| Septic controls | TNF-α | SAE | Li 2022 | 17 | 1.6 | 21 | 15.1 | 1.1 | 20 | 1.35 (0.67 to 2.04) | 23.3 |
| Septic controls | TNF-α | Delirium only | Erikson 2018 | 43 | 33.185 | 10 | 33.166 | 17.33 | 12 | 0.37 (-0.48 to 1.22) | 18.9 |
| Septic controls | TNF-α | Delirium only | Jorge-Ripper 2016 | 18 | 28.1082 | 47 | 12.5 | 24.2559 | 35 | 0.21 (-0.23 to 0.64) | 31.5 |
| Septic controls | TNF-α | Delirium only | Lei 2022 | 14.17 | 6 | 17 | 12.02 | 2.96 | 35 | 0.51 (-0.08 to 1.09) | 26.4 |

[a] Smith 2022 IL-17: control group reported as 0 (0–0); an SD of 0.074 was imputed to allow estimation.

#### eTable 17. Expected Review Manager output for each panel and stratum

| Framework | Panel | Level | k | Index n | Control n | SMD | 95% CI | τ² | Chi² | df | p for heterogeneity | I², % | Z | p for effect |
|---|---|---|---|---|---|---|---|---|---|---|---|---|---|---|
| Non-septic controls | IL-6 | SAE | 2 | 101 | 57 | 0.87 | 0.52 to 1.22 | 0.00 | 0.59 | 1 | 0.4425 | 0 | 4.85 | 0.0000 |
| Non-septic controls | IL-6 | Delirium only | 5 | 827 | 487 | 0.43 | 0.13 to 0.73 | 0.07 | 12.78 | 4 | 0.0124 | 69 | 2.82 | 0.0047 |
| Non-septic controls | IL-6 | Total | 7 | 928 | 544 | 0.54 | 0.25 to 0.82 | 0.10 | 21.77 | 6 | 0.0013 | 72 | 3.70 | 0.0002 |
| Non-septic controls | IL-6 | Subgroup difference | | | | | | | 3.40 | 1 | 0.0651 | 70.6 | | |
| Non-septic controls | IL-8 | SAE | 2 | 101 | 57 | 0.63 | 0.29 to 0.98 | 0.00 | 0.72 | 1 | 0.3969 | 0 | 3.61 | 0.0003 |
| Non-septic controls | IL-8 | Delirium only | 2 | 732 | 378 | 0.33 | -0.13 to 0.80 | 0.09 | 5.30 | 1 | 0.0214 | 81 | 1.40 | 0.1611 |
| Non-septic controls | IL-8 | Total | 4 | 833 | 435 | 0.47 | 0.12 to 0.82 | 0.09 | 11.86 | 3 | 0.0079 | 75 | 2.62 | 0.0088 |
| Non-septic controls | IL-8 | Subgroup difference | | | | | | | 1.03 | 1 | 0.3091 | 3.3 | | |
| Non-septic controls | IFN-γ | SAE | 2 | 101 | 57 | 0.17 | -0.17 to 0.50 | 0.00 | 0.33 | 1 | 0.5652 | 0 | 0.98 | 0.3277 |
| Non-septic controls | IFN-γ | Delirium only | 2 | 738 | 327 | 0.23 | -0.30 to 0.75 | 0.12 | 4.58 | 1 | 0.0323 | 78 | 0.85 | 0.3973 |
| Non-septic controls | IFN-γ | Total | 4 | 839 | 384 | 0.15 | -0.09 to 0.39 | 0.03 | 5.36 | 3 | 0.1474 | 44 | 1.24 | 0.2167 |
| Non-septic controls | IFN-γ | Subgroup difference | | | | | | | 0.03 | 1 | 0.8519 | 0.0 | | |
| Non-septic controls | IL-12 | SAE | 2 | 101 | 57 | -0.34 | -1.07 to 0.38 | 0.21 | 3.97 | 1 | 0.0463 | 75 | 0.93 | 0.3536 |
| Non-septic controls | IL-12 | Delirium only | 2 | 732 | 378 | 0.12 | -0.33 to 0.57 | 0.09 | 5.00 | 1 | 0.0253 | 80 | 0.53 | 0.5953 |
| Non-septic controls | IL-12 | Total | 4 | 833 | 435 | -0.08 | -0.46 to 0.30 | 0.11 | 14.10 | 3 | 0.0028 | 79 | 0.41 | 0.6819 |
| Non-septic controls | IL-12 | Subgroup difference | | | | | | | 1.14 | 1 | 0.2853 | 12.4 | | |
| Non-septic controls | TNF-α | SAE | 2 | 101 | 57 | -0.79 | -2.11 to 0.54 | 0.84 | 12.51 | 1 | 0.0004 | 92 | 1.16 | 0.2451 |
| Non-septic controls | TNF-α | Delirium only | 4 | 786 | 409 | 0.32 | -0.10 to 0.75 | 0.14 | 14.45 | 3 | 0.0024 | 79 | 1.49 | 0.1373 |
| Non-septic controls | TNF-α | Total | 6 | 887 | 466 | -0.02 | -0.56 to 0.52 | 0.39 | 54.82 | 5 | 0.0000 | 91 | 0.06 | 0.9482 |
| Non-septic controls | TNF-α | Subgroup difference | | | | | | | 2.44 | 1 | 0.1180 | 59.1 | | |
| Non-septic controls | IL-1β | SAE | 2 | 101 | 57 | 0.16 | -0.18 to 0.49 | 0.00 | 0.48 | 1 | 0.4904 | 0 | 0.90 | 0.3655 |
| Non-septic controls | IL-1β | Delirium only | 3 | 763 | 425 | 0.31 | -0.02 to 0.65 | 0.06 | 6.52 | 2 | 0.0384 | 69 | 1.85 | 0.0647 |
| Non-septic controls | IL-1β | Total | 5 | 864 | 482 | 0.24 | 0.03 to 0.45 | 0.02 | 7.03 | 4 | 0.1343 | 43 | 2.25 | 0.0243 |
| Non-septic controls | IL-1β | Subgroup difference | | | | | | | 0.43 | 1 | 0.5107 | 0.0 | | |
| Non-septic controls | IL-10 | SAE | 2 | 101 | 57 | 0.26 | -0.11 to 0.64 | 0.01 | 1.18 | 1 | 0.2768 | 15 | 1.38 | 0.1664 |
| Non-septic controls | IL-10 | Delirium only | 5 | 827 | 487 | 0.14 | -0.18 to 0.46 | 0.09 | 14.94 | 4 | 0.0048 | 73 | 0.85 | 0.3974 |
| Non-septic controls | IL-10 | Total | 7 | 928 | 544 | 0.16 | -0.09 to 0.41 | 0.07 | 17.41 | 6 | 0.0079 | 66 | 1.27 | 0.2046 |
| Non-septic controls | IL-10 | Subgroup difference | | | | | | | 0.25 | 1 | 0.6152 | 0.0 | | |
| Non-septic controls | IL-17 | SAE | 2 | 101 | 57 | 0.20 | -0.14 to 0.53 | 0.00 | 0.54 | 1 | 0.4619 | 0 | 1.15 | 0.2508 |
| Non-septic controls | IL-17 | Delirium only | 1 | 41 | 78 | 0.67 | 0.29 to 1.06 | 0.00 | — | — | — | — | 3.41 | 0.0007 |
| Non-septic controls | IL-17 | Total | 3 | 142 | 135 | 0.37 | 0.01 to 0.73 | 0.05 | 3.87 | 2 | 0.1446 | 48 | 1.99 | 0.0469 |
| Non-septic controls | IL-17 | Subgroup difference | | | | | | | 3.33 | 1 | 0.0682 | 69.9 | | |
| Non-septic controls | sTNFR-1 | Delirium only | 2 | 722 | 347 | 0.37 | -0.15 to 0.89 | 0.12 | 4.79 | 1 | 0.0287 | 79 | 1.39 | 0.1651 |
| Non-septic controls | sTNFR-1 | Total | 2 | 722 | 347 | 0.37 | -0.15 to 0.89 | 0.12 | 4.79 | 1 | 0.0287 | 79 | 1.39 | 0.1651 |
| Non-septic controls | CRP | Delirium only | 1 | 691 | 300 | 0.13 | -0.01 to 0.27 | 0.00 | — | — | — | — | 1.88 | 0.0602 |
| Non-septic controls | CRP | Total | 1 | 691 | 300 | 0.13 | -0.01 to 0.27 | 0.00 | — | — | — | — | 1.88 | 0.0602 |
| Septic controls | CRP | SAE | 1 | 12 | 4 | 1.11 | -0.10 to 2.33 | 0.00 | — | — | — | — | 1.79 | 0.0731 |
| Septic controls | CRP | Delirium only | 1 | 10 | 12 | -0.50 | -1.35 to 0.36 | 0.00 | — | — | — | — | 1.14 | 0.2535 |
| Septic controls | CRP | Total | 2 | 22 | 16 | 0.25 | -1.33 to 1.82 | 1.01 | 4.51 | 1 | 0.0338 | 78 | 0.31 | 0.7588 |

| Framework | Panel | Level | k | Index n | Control n | SMD | 95% CI | τ² | Chi² | df | p for heterogeneity | I², % | Z | p for effect |
|---|---|---|---|---|---|---|---|---|---|---|---|---|---|---|
| Septic controls | CRP | Subgroup difference | | | | | | | 4.51 | 1 | 0.0338 | 77.8 | | |
| Septic controls | IFN-γ | SAE | 1 | 62 | 78 | 0.64 | 0.30 to 0.99 | 0.00 | — | — | — | — | 3.69 | 0.0002 |
| Septic controls | IFN-γ | Delirium only | 1 | 47 | 35 | 0.41 | -0.04 to 0.85 | 0.00 | — | — | — | — | 1.80 | 0.0724 |
| Septic controls | IFN-γ | Total | 2 | 109 | 113 | 0.55 | 0.28 to 0.83 | 0.00 | 0.70 | 1 | 0.4021 | 0 | 4.02 | 0.0001 |
| Septic controls | IFN-γ | Subgroup difference | | | | | | | 0.70 | 1 | 0.4021 | 0.0 | | |
| Septic controls | IL-6 | SAE | 9 | 361 | 404 | 1.38 | 0.83 to 1.94 | 0.60 | 77.50 | 8 | 0.0000 | 90 | 4.91 | 0.0000 |
| Septic controls | IL-6 | Delirium only | 3 | 74 | 82 | -0.02 | -0.37 to 0.33 | 0.01 | 2.24 | 2 | 0.3256 | 11 | 0.12 | 0.9071 |
| Septic controls | IL-6 | Total | 12 | 435 | 486 | 1.05 | 0.51 to 1.59 | 0.80 | 134.32 | 11 | 0.0000 | 92 | 3.80 | 0.0001 |
| Septic controls | IL-6 | Subgroup difference | | | | | | | 17.72 | 1 | 0.0000 | 94.4 | | |
| Septic controls | IL-10 | SAE | 2 | 83 | 98 | 1.98 | -0.45 to 4.40 | 2.92 | 22.49 | 1 | 0.0000 | 96 | 1.60 | 0.1097 |
| Septic controls | IL-10 | Delirium only | 2 | 64 | 70 | 0.14 | -0.21 to 0.49 | 0.00 | 0.27 | 1 | 0.6005 | 0 | 0.78 | 0.4368 |
| Septic controls | IL-10 | Total | 4 | 147 | 168 | 0.97 | 0.05 to 1.88 | 0.77 | 37.21 | 3 | 0.0000 | 92 | 2.08 | 0.0378 |
| Septic controls | IL-10 | Subgroup difference | | | | | | | 2.17 | 1 | 0.1410 | 53.8 | | |
| Septic controls | NSE | SAE | 4 | 107 | 153 | 1.86 | 0.54 to 3.19 | 1.63 | 51.29 | 3 | 0.0000 | 94 | 2.76 | 0.0059 |
| Septic controls | NSE | Delirium only | 1 | 10 | 12 | -0.06 | -0.90 to 0.78 | 0.00 | — | — | — | — | 0.13 | 0.8937 |
| Septic controls | NSE | Total | 5 | 117 | 165 | 1.45 | 0.33 to 2.57 | 1.43 | 55.67 | 4 | 0.0000 | 93 | 2.55 | 0.0109 |
| Septic controls | NSE | Subgroup difference | | | | | | | 5.76 | 1 | 0.0164 | 82.6 | | |
| Septic controls | S100B | SAE | 6 | 178 | 202 | 1.87 | 0.76 to 2.97 | 1.62 | 85.16 | 5 | 0.0000 | 94 | 3.32 | 0.0009 |
| Septic controls | S100B | Delirium only | 2 | 57 | 47 | 0.29 | -0.45 to 1.03 | 0.18 | 2.45 | 1 | 0.1175 | 59 | 0.76 | 0.4462 |
| Septic controls | S100B | Total | 8 | 235 | 249 | 1.39 | 0.54 to 2.25 | 1.29 | 102.67 | 7 | 0.0000 | 93 | 3.21 | 0.0013 |
| Septic controls | S100B | Subgroup difference | | | | | | | 5.40 | 1 | 0.0201 | 81.5 | | |
| Septic controls | TNF-α | SAE | 1 | 21 | 20 | 1.35 | 0.67 to 2.04 | 0.00 | — | — | — | — | 3.86 | 0.0001 |
| Septic controls | TNF-α | Delirium only | 3 | 74 | 82 | 0.32 | -0.00 to 0.65 | 0.00 | 0.66 | 2 | 0.7185 | 0 | 1.94 | 0.0527 |
| Septic controls | TNF-α | Total | 4 | 95 | 102 | 0.58 | 0.08 to 1.08 | 0.15 | 7.75 | 3 | 0.0516 | 61 | 2.29 | 0.0218 |
| Septic controls | TNF-α | Subgroup difference | | | | | | | 7.08 | 1 | 0.0078 | 85.9 | | |

Random-effects (DerSimonian-Laird) standardized mean differences recomputed from the arm-level values of eTable 16; these reproduce the forest panels of eFigures 1 and 2.

#### eTable 18. Framework audit of every plotted row

| Biomarker | Framework plotted | Study | Workbook comparison types | Frameworks supported | Verdict |
|---|---|---|---|---|---|
| CRP | Non-septic controls | Brummel 2024 | Delirium vs non-delirium (mixed ICU population) | Non-septic controls | consistent |
| CRP | Non-septic controls | Smith 2022 | | | removed: CRP not reported in the source (see eTable 13) |
| IFN-γ | Non-septic controls | Brummel 2024 | Delirium vs non-delirium (mixed ICU population) | Non-septic controls | consistent |
| IFN-γ | Non-septic controls | Jorge-Ripper 2016 | Delirium vs Control; Delirium vs non-delirium (within sepsis); Sepsis vs non-sepsis (within delirium) | Non-septic controls; Septic controls | corrected: index arm is delirium with sepsis vs healthy controls (see eTable 13) |
| IFN-γ | Non-septic controls | Mao 2023 | SAE vs non-sepsis critical illness | Non-septic controls | consistent; cerebrospinal fluid (see eTable 26) |
| IFN-γ | Non-septic controls | Orhun 2019 | Delirium vs coma (within SIBD/sepsis); SIBD vs healthy controls | other; Non-septic controls | consistent |
| IL-10 | Non-septic controls | Brummel 2024 | Delirium vs non-delirium (mixed ICU population) | Non-septic controls | consistent |
| IL-10 | Non-septic controls | Lei 2022 | Delirium vs Non Delirium (within sepsis); Sepsis vs non-septic | Non-septic controls; Septic controls | consistent |
| IL-10 | Non-septic controls | Mao 2023 | SAE vs non-sepsis critical illness | Non-septic controls | consistent; cerebrospinal fluid (see eTable 26) |
| IL-10 | Non-septic controls | Orhun 2019 | Delirium vs coma (within SIBD/sepsis); SIBD vs healthy controls; Septic shock vs no septic shock (within SIBD/sepsis) | other; Non-septic controls | consistent |
| IL-10 | Non-septic controls | Ritter 2014 | Delirium vs non-delirium (mixed ICU population) | Non-septic controls | consistent |
| IL-10 | Non-septic controls | Smith 2022 | Delirium vs non-delirium (mixed ICU population); Sepsis vs non-sepsis (within delirium) | Non-septic controls | consistent |
| IL-12 | Non-septic controls | Brummel 2024 | Delirium vs non-delirium (mixed ICU population) | Non-septic controls | consistent |
| IL-12 | Non-septic controls | Mao 2023 | SAE vs non-sepsis critical illness | Non-septic controls | consistent; cerebrospinal fluid (see eTable 26) |
| IL-12 | Non-septic controls | Orhun 2019 | Delirium vs coma (within SIBD/sepsis); SIBD vs healthy controls | other; Non-septic controls | consistent |
| IL-12 | Non-septic controls | Smith 2022 | Delirium vs non-delirium (mixed ICU population); Sepsis vs non-sepsis (within delirium) | Non-septic controls | consistent |
| IL-17 | Non-septic controls | Mao 2023 | SAE vs non-sepsis critical illness | Non-septic controls | consistent; cerebrospinal fluid (see eTable 26) |
| IL-17 | Non-septic controls | Orhun 2019 | Delirium vs coma (within SIBD/sepsis); SIBD vs healthy controls | other; Non-septic controls | consistent |
| IL-17 | Non-septic controls | Smith 2022 | Delirium vs non-delirium (mixed ICU population); Sepsis vs non-sepsis (within delirium) | Non-septic controls | consistent |
| IL-1β | Non-septic controls | Brummel 2024 | Delirium vs non-delirium (mixed ICU population) | Non-septic controls | consistent |
| IL-1β | Non-septic controls | Mao 2023 | SAE vs non-sepsis critical illness | Non-septic controls | consistent; cerebrospinal fluid (see eTable 26) |
| IL-1β | Non-septic controls | Orhun 2019 | Delirium vs coma (within SIBD/sepsis); SIBD vs healthy controls; Septic shock vs no septic shock (within SIBD/sepsis) | other; Non-septic controls | consistent |
| IL-1β | Non-septic controls | Ritter 2014 | Delirium vs non-delirium (mixed ICU population) | Non-septic controls | consistent |
| IL-1β | Non-septic controls | Smith 2022 | Delirium vs non-delirium (mixed ICU population); Sepsis vs non-sepsis (within delirium) | Non-septic controls | consistent |
| IL-6 | Non-septic controls | Brummel 2024 | Delirium vs non-delirium (mixed ICU population) | Non-septic controls | consistent |
| IL-6 | Non-septic controls | Jorge-Ripper 2016 | Delirium vs Control; Delirium vs non-delirium (within sepsis); Sepsis vs non-sepsis (within delirium) | Non-septic controls; Septic controls | corrected: index arm is delirium with sepsis vs healthy controls (see eTable 13) |
| IL-6 | Non-septic controls | Lei 2022 | Delirium vs Non Delirium (within sepsis); Sepsis vs non-septic | Non-septic controls; Septic controls | consistent |
| IL-6 | Non-septic controls | Mao 2023 | SAE vs non-sepsis critical illness | Non-septic controls | consistent; cerebrospinal fluid (see eTable 26) |

| Biomarker | Framework plotted | Study | Workbook comparison types | Frameworks supported | Verdict |
|---|---|---|---|---|---|
| IL-6 | Non-septic controls | Orhun 2019 | Delirium vs coma (within SIBD/sepsis); SIBD vs healthy controls | other; Non-septic controls | consistent |
| IL-6 | Non-septic controls | Ritter 2014 | Delirium vs non-delirium (mixed ICU population) | Non-septic controls | consistent |
| IL-6 | Non-septic controls | Smith 2022 | Delirium vs non-delirium (mixed ICU population); Sepsis vs non-sepsis (within delirium) | Non-septic controls | consistent |
| IL-8 | Non-septic controls | Brummel 2024 | Delirium vs non-delirium (mixed ICU population) | Non-septic controls | consistent |
| IL-8 | Non-septic controls | Mao 2023 | SAE vs non-sepsis critical illness | Non-septic controls | consistent; cerebrospinal fluid (see eTable 26) |
| IL-8 | Non-septic controls | Orhun 2019 | Delirium vs coma (within SIBD/sepsis); SIBD vs healthy controls; Septic shock vs no septic shock (within SIBD/sepsis) | other; Non-septic controls | consistent |
| IL-8 | Non-septic controls | Smith 2022 | Delirium vs non-delirium (mixed ICU population); Sepsis vs non-sepsis (within delirium) | Non-septic controls | consistent |
| TNF-α | Non-septic controls | Brummel 2024 | Delirium vs non-delirium (mixed ICU population) | Non-septic controls | consistent |
| TNF-α | Non-septic controls | Jorge-Ripper 2016 | Delirium vs Control; Delirium vs non-delirium (within sepsis); Sepsis vs non-sepsis (within delirium) | Non-septic controls; Septic controls | corrected: index arm is delirium with sepsis vs healthy controls (see eTable 13) |
| TNF-α | Non-septic controls | Lei 2022 | Delirium vs Non Delirium (within sepsis); Sepsis vs non-septic | Non-septic controls; Septic controls | consistent |
| TNF-α | Non-septic controls | Mao 2023 | SAE vs non-sepsis critical illness | Non-septic controls | consistent; cerebrospinal fluid (see eTable 26) |
| TNF-α | Non-septic controls | Orhun 2019 | Delirium vs coma (within SIBD/sepsis); SIBD vs healthy controls | other; Non-septic controls | consistent |
| TNF-α | Non-septic controls | Ritter 2014 | Delirium vs non-delirium (mixed ICU population) | Non-septic controls | consistent |
| sTNFR-1 | Non-septic controls | Brummel 2024 | Delirium vs non-delirium (mixed ICU population) | Non-septic controls | consistent |
| sTNFR-1 | Non-septic controls | Ritter 2014 | Delirium vs non-delirium (mixed ICU population) | Non-septic controls | consistent |
| CRP | Septic controls | Erikson 2018 | Delirium vs non-delirium (within sepsis) | Septic controls | consistent |
| CRP | Septic controls | Pfister 2008 | Delirium vs non-delirium (within sepsis) | Septic controls | consistent |
| IFN-γ | Septic controls | Jorge-Ripper 2016 | Delirium vs Control; Delirium vs non-delirium (within sepsis); Sepsis vs non-sepsis (within delirium) | Non-septic controls; Septic controls | consistent |
| IFN-γ | Septic controls | Yu 2025 | SAE vs no SAE (within sepsis) | Septic controls | consistent |
| IL-10 | Septic controls | Jorge-Ripper 2016 | Delirium vs Control; Delirium vs non-delirium (within sepsis); Sepsis vs non-sepsis (within delirium) | Non-septic controls; Septic controls | consistent |
| IL-10 | Septic controls | Lei 2022 | Delirium vs Non Delirium (within sepsis); Sepsis vs non-septic | Non-septic controls; Septic controls | consistent |
| IL-10 | Septic controls | Li 2022 | SAE vs no SAE (within sepsis) | Septic controls | consistent |
| IL-10 | Septic controls | Yu 2025 | SAE vs no SAE (within sepsis) | Septic controls | consistent |
| IL-6 | Septic controls | Erikson 2018 | Delirium vs non-delirium (within sepsis) | Septic controls | consistent |
| IL-6 | Septic controls | Feng 2017 | SAE vs non-SAE (within sepsis) | Septic controls | consistent |
| IL-6 | Septic controls | Guo 2022 | SAE vs non-SAE (within sepsis) | Septic controls | consistent |
| IL-6 | Septic controls | Jorge-Ripper 2016 | Delirium vs Control; Delirium vs non-delirium (within sepsis); Sepsis vs non-sepsis (within delirium) | Non-septic controls; Septic controls | consistent |
| IL-6 | Septic controls | Lei 2022 | Delirium vs Non Delirium (within sepsis); Sepsis vs non-septic | Non-septic controls; Septic controls | consistent |
| IL-6 | Septic controls | Li 2022 | SAE vs no SAE (within sepsis) | Septic controls | consistent |
| IL-6 | Septic controls | Nakamura 2003 | SAE vs no SAE (within sepsis); Secondary: septic controls vs non-septic controls; Secondary: septic encephalopathy vs non-septic controls | other; Non-septic controls; Septic controls | consistent |
| IL-6 | Septic controls | Pfister 2008 | Delirium vs non-delirium (within sepsis) | Septic controls | consistent |
| IL-6 | Septic controls | Wang 2022 | SAE vs no SAE (within sepsis) | Septic controls | consistent |
| IL-6 | Septic controls | Wei 2025 | SAE vs no SAE (within sepsis) | Septic controls | consistent |
| IL-6 | Septic controls | Wu 2020 | SAE vs no SAE (within sepsis) | Septic controls | consistent |

| Biomarker | Framework plotted | Study | Workbook comparison types | Frameworks supported | Verdict |
|---|---|---|---|---|---|
| IL-6 | Septic controls | Yu 2025 | SAE vs no SAE (within sepsis) | Septic controls | consistent |
| NSE | Septic controls | Erikson 2018 | Delirium vs non-delirium (within sepsis) | Septic controls | consistent |
| NSE | Septic controls | Feng 2017 | SAE vs non-SAE (within sepsis) | Septic controls | consistent |
| NSE | Septic controls | Guo 2022 | SAE vs non-SAE (within sepsis) | Septic controls | consistent |
| NSE | Septic controls | Li 2022 | SAE vs no SAE (within sepsis) | Septic controls | consistent |
| NSE | Septic controls | Wang 2022 | SAE vs no SAE (within sepsis) | Septic controls | consistent |
| S100B | Septic controls | Erikson 2018 | Delirium vs non-delirium (within sepsis) | Septic controls | consistent |
| S100B | Septic controls | Feng 2017 | SAE vs non-SAE (within sepsis) | Septic controls | consistent |
| S100B | Septic controls | Guo 2022 | SAE vs non-SAE (within sepsis) | Septic controls | consistent |
| S100B | Septic controls | Jorge-Ripper 2016 | Delirium vs Control; Delirium vs non-delirium (within sepsis); Sepsis vs non-sepsis (within delirium) | Non-septic controls; Septic controls | consistent |
| S100B | Septic controls | Li 2022 | SAE vs no SAE (within sepsis) | Septic controls | consistent |
| S100B | Septic controls | Pfister 2008 | Delirium vs non-delirium (within sepsis) | Septic controls | consistent |
| S100B | Septic controls | Wang 2022 | SAE vs no SAE (within sepsis) | Septic controls | consistent |
| S100B | Septic controls | Wu 2020 | SAE vs no SAE (within sepsis) | Septic controls | consistent |
| TNF-α | Septic controls | Erikson 2018 | Delirium vs non-delirium (within sepsis) | Septic controls | consistent |
| TNF-α | Septic controls | Jorge-Ripper 2016 | Delirium vs Control; Delirium vs non-delirium (within sepsis); Sepsis vs non-sepsis (within delirium) | Non-septic controls; Septic controls | consistent |
| TNF-α | Septic controls | Lei 2022 | Delirium vs Non Delirium (within sepsis); Sepsis vs non-septic | Non-septic controls; Septic controls | consistent |
| TNF-α | Septic controls | Li 2022 | SAE vs no SAE (within sepsis) | Septic controls | consistent |

Workbook comparison type of every plotted row, the framework it supports, and the verdict. SIBD = sepsis-induced brain dysfunction.

#### eTable 19. Reproduction of the pooled odds ratios from study-level log odds

| Biomarker | Framework | k | OR, refit | 95% CI low | 95% CI high | $\tau^2$ | $I^2$, % | OR, printed | Low, printed | High, printed | $I^2$, printed | Maximum relative deviation |
|---|---|---|---|---|---|---|---|---|---|---|---|---|
| IFN-γ | Association (OR) | 2 | 0.9 | 0.805 | 1.005 | 0.0 | 0.0 | 0.9 | 0.8 | 1.01 | 0 | 0.006 |
| IL-1β | Association (OR) | 2 | 1.02 | 1.007 | 1.032 | 0.0 | 0.0 | 1.02 | 1.01 | 1.03 | 0 | 0.0026 |
| IL-6 | Association (OR) | 4 | 1.853 | 1.5 | 2.29 | 0.016 | 34.2 | 1.85 | 1.5 | 2.29 | 34 | 0.0017 |
| S100B | Association (OR) | 2 | 3.491 | 0.222 | 54.986 | 3.201 | 79.0 | 3.49 | 0.22 | 54.98 | 79 | 0.0076 |

Reproduction of the four pooled odds ratios of eFigure 3 from the study-level log odds ratios plotted in those panels.

#### eTable 20. Deviations from the registered protocol

| # | Registered plan | What was done, and why | Effect on eligibility, outcomes or conclusions |
|---|---|---|---|
| 1 | Certainty of evidence rated in GRADE working tables prepared during the original analysis | Certainty was re-derived from the pooled estimates using the mechanical rules of eAppendix 2, section 2.9, because the domain judgments in the original working tables could not be reconciled with their own recorded downgrades | None on eligibility or outcomes. Certainty is now reproducible from the reported estimates by a stated rule |
| 2 | Small-study effects reported for the two comparisons tabulated in the analysis output | Reported for one comparison. The second Egger and trim-and-fill block is labeled as the non-septic-control framework but is fitted on 11 comparisons and could not be reconciled with any analysis in that framework; it is not reported | Narrows one sensitivity analysis. No pooled estimate and no conclusion depends on the unreconciled block |
| 3 | Vote counting on statistical significance | Vote counting implemented on the direction of the point estimate, following the Cochrane Handbook; a correction to the analysis as executed, not a change of question | Changes the vote-count table only, in the direction the Handbook prescribes |
| 4 | Not in the registered analysis plan | A 95% prediction interval was added, because the observed heterogeneity makes a confidence interval alone an inadequate summary. It was computed for all 14 comparisons with three or more studies from a study-level refit that reproduces every pooled estimate to within 0.01 (eAppendix 2; eTable 9) | Addition only; reported alongside, not instead of, the confidence interval |
| 5 | Not in the registered analysis plan | Implied discrimination (area under the curve) was added, because a candidate biomarker is judged clinically on how well it separates patients rather than on a standardized difference. The transformation uses no data beyond the pooled estimates already reported | Addition only; no new data and no change to any pooled estimate |
| 6 | Eligible biomarkers measured in blood or cerebrospinal fluid | Both compartments were eligible; one included study (Mao 2023) reported cerebrospinal fluid only. Its comparisons enter the non-septic-control framework, and the prespecified compartment analysis is reported as a sensitivity analysis (eTable 26) | None on eligibility; no conclusion changes without cerebrospinal fluid data |
| 7 | Subgroup analysis by timing of sampling (early vs late sepsis) | Not performed: sampling time was unreported or not comparable across studies (admission, diagnosis, day 1 to day 5), so no study-level classification was reproducible | Timing remains an unexplored source of heterogeneity (Discussion) |
| 8 | Meta-regression when at least 10 studies are available (compartment, timing, sepsis phenotype, outcome definition, assay, baseline severity) | Not performed: only IL-6 versus septic controls reached 10 studies, and the candidate moderators were unreported or invariant across them; the instrument and phenotype contrasts were examined by subgroup analysis instead (eTables 5, 11 and 12) | Effect modification assessed by subgroup rather than regression; interpretation unchanged |
| 9 | Analyses in R (meta or metafor) | Pooled analyses were run in Review Manager; small-study effects, prediction intervals, sensitivity analyses and the prioritization index were computed from the arm-level data of eTable 16 with the same DerSimonian-Laird model | None; the refit reproduces every pooled estimate to within 0.01 |
| 10 | Pooling of correlation coefficients, dose-response analysis across biomarker categories, and adjusted-versus-unadjusted association sensitivity | Not performed: fewer than two studies reported comparable correlations with the neurological outcome or categorical dose-response data, and the four pooled odds ratios came from models too few to separate adjusted from unadjusted estimates | The association framework is reported as pooled odds ratios only |
| 11 | Secondary outcomes: severity scores, ICU length of stay, mortality, duration of ventilation and cognitive outcomes | Not synthesized: the review question was narrowed to the neurological phenotype, and these outcomes were not reported by neurological group in a poolable form | Stated as a limitation; no bearing on the primary comparisons |
| 12 | Th17-related markers (IL-17F, IL-22, IL-23, RORγt, Th17 cell frequency) and Th2 and Th9 comparators (IL-4, IL-5, IL-13, IL-9) | None was reported by neurological group in a poolable form by two or more included studies, so none entered the synthesis | The Th17 hypothesis is tested through IL-17A only (Discussion) |
| 13 | Evidence map of animal and mechanistic studies for translational purposes | Not constructed; mechanistic evidence is cited narratively in the Introduction and Discussion | None on the quantitative synthesis |

The registered plan, what was done instead and why, and the effect of each deviation on the eligibility criteria, the outcome definitions and the conclusions; PRISMA 2020 item 24c.

#### eTable 21. Direction of evidence by vote counting

| Biomarker | vs non-septic controls (higher / no difference / lower) | vs septic controls (higher / no difference / lower) | All comparisons (higher / no difference / lower) | Higher in SAE/delirium |
|---|---|---|---|---|
| IL-17 | 3 / 0 / 0 (k = 3) | 1 / 0 / 0 (k = 1) | 4 / 0 / 0 (k = 4) | 100% |
| IL-6 | 6 / 0 / 1 (k = 7) | 10 / 0 / 2 (k = 12) | 16 / 0 / 3 (k = 19) | 84% |
| IL-1β | 4 / 0 / 1 (k = 5) | - | 4 / 0 / 1 (k = 5) | 80% |
| IFN-γ | 4 / 0 / 0 (k = 4) | 2 / 0 / 0 (k = 2) | 6 / 0 / 0 (k = 6) | 100% |
| IL-12 | 2 / 0 / 2 (k = 4) | - | 2 / 0 / 2 (k = 4) | 50% |
| TNF-α | 3 / 0 / 3 (k = 6) | 4 / 0 / 0 (k = 4) | 7 / 0 / 3 (k = 10) | 70% |
| IL-8 | 4 / 0 / 0 (k = 4) | - | 4 / 0 / 0 (k = 4) | 100% |
| CRP | 1 / 0 / 0 (k = 1) | 1 / 0 / 1 (k = 2) | 2 / 0 / 1 (k = 3) | 67% |
| sTNFR-1 | 2 / 0 / 0 (k = 2) | - | 2 / 0 / 0 (k = 2) | 100% |
| IL-10 | 5 / 1 / 1 (k = 7) | 4 / 0 / 0 (k = 4) | 9 / 1 / 1 (k = 11) | 82% |
| S100B | 2 / 0 / 0 (k = 2) | 7 / 0 / 1 (k = 8) | 9 / 0 / 1 (k = 10) | 90% |
| NSE | 1 / 0 / 0 (k = 1) | 4 / 0 / 1 (k = 5) | 5 / 0 / 1 (k = 6) | 83% |

Each comparison is classified by the sign of its point estimate; statistical significance is not used. "No difference" is reserved for an exactly null point estimate, and comparisons without an estimable direction are excluded. Provenance of the non-septic-control counts follows eTable 16.

#### eTable 22. Harmonized pooled estimates across the three analysis frameworks

| Framework | Biomarker | k | Participants, n | Effect estimate | 95% CI | p | I² | 95% CI excludes the null |
|---|---|---|---|---|---|---|---|---|
| SAE/delirium vs non-septic controls | IL-17 | 3 | 277 | 0.37 | 0.01 to 0.73 | 0.05 | 48% | Yes |
| SAE/delirium vs non-septic controls | IL-6 | 7 | 1472 | 0.54 | 0.25 to 0.82 | <0.001 | 72% | Yes |
| SAE/delirium vs non-septic controls | IL-1β | 5 | 1346 | 0.24 | 0.03 to 0.45 | 0.02 | 43% | Yes |
| SAE/delirium vs non-septic controls | IFN-γ | 4 | 1223 | 0.15 | -0.09 to 0.39 | 0.22 | 44% | No |
| SAE/delirium vs non-septic controls | IL-12 | 4 | 1268 | -0.08 | -0.46 to 0.30 | 0.68 | 79% | No |
| SAE/delirium vs non-septic controls | TNF-α | 6 | 1353 | -0.02 | -0.56 to 0.52 | 0.95 | 91% | No |
| SAE/delirium vs non-septic controls | IL-8 | 4 | 1268 | 0.47 | 0.12 to 0.82 | 0.009 | 75% | Yes |
| SAE/delirium vs non-septic controls | sTNFR-1 | 2 | 1069 | 0.37 | -0.15 to 0.89 | 0.17 | 79% | No |
| SAE/delirium vs non-septic controls | IL-10 | 7 | 1472 | 0.16 | -0.09 to 0.41 | 0.20 | 66% | No |
| SAE/delirium vs septic controls | IL-6 | 12 | 921 | 1.05 | 0.51 to 1.59 | <0.001 | 92% | Yes |
| SAE/delirium vs septic controls | IFN-γ | 2 | 222 | 0.55 | 0.28 to 0.83 | <0.001 | 0% | Yes |
| SAE/delirium vs septic controls | TNF-α | 4 | 197 | 0.58 | 0.08 to 1.08 | 0.02 | 61% | Yes |
| SAE/delirium vs septic controls | CRP | 2 | 38 | 0.25 | -1.33 to 1.82 | 0.76 | 78% | No |
| SAE/delirium vs septic controls | IL-10 | 4 | 315 | 0.97 | 0.05 to 1.88 | 0.04 | 92% | Yes |
| SAE/delirium vs septic controls | S100B | 8 | 484 | 1.39 | 0.54 to 2.25 | 0.001 | 93% | Yes |
| SAE/delirium vs septic controls | NSE | 5 | 282 | 1.45 | 0.33 to 2.57 | 0.01 | 93% | Yes |
| Association with SAE/delirium (OR) | IL-6 | 4 | - | 1.85 | 1.50 to 2.29 | <0.00001 | 34% | Yes |
| Association with SAE/delirium (OR) | IFN-γ | 2 | - | 0.90 | 0.80 to 1.01 | 0.06 | 0% | No |
| Association with SAE/delirium (OR) | IL-1β | 2 | - | 1.02 | 1.01 to 1.03 | 0.002 | 0% | Yes |
| Association with SAE/delirium (OR) | S100B | 2 | - | 3.49 | 0.22 to 54.98 | 0.37 | 79% | No |

Estimates are random-effects standardized mean differences for the two contrast frameworks and odds ratios for the association framework; the scales are not interchangeable and are never pooled across frameworks. CRP versus non-septic controls (one study) and IL-17 versus septic controls (one study) are not pooled.

#### eTable 23. Implied discrimination of each pooled biomarker difference

| Framework | Biomarker | k | Pooled SMD | 95% CI (SMD) | Implied AUC | 95% CI (AUC) | $I^2$ |
|---|---|---|---|---|---|---|---|
| vs septic controls | NSE | 5 | 1.45 | 0.33 to 2.57 | 0.85 | 0.59 to 0.97 | 93% |
| vs septic controls | S100B | 8 | 1.39 | 0.54 to 2.25 | 0.84 | 0.65 to 0.94 | 93% |
| vs septic controls | IL-6 | 12 | 1.05 | 0.51 to 1.59 | 0.77 | 0.64 to 0.87 | 92% |
| vs septic controls | IL-10 | 4 | 0.97 | 0.05 to 1.88 | 0.75 | 0.52 to 0.91 | 92% |
| vs septic controls | TNF-α | 4 | 0.58 | 0.08 to 1.08 | 0.66 | 0.52 to 0.78 | 61% |
| vs septic controls | IFN-γ | 2 | 0.55 | 0.28 to 0.83 | 0.65 | 0.58 to 0.72 | 0% |
| vs septic controls | CRP | 2 | 0.25 | -1.33 to 1.82 | 0.57 | 0.17 to 0.90 | 78% |
| vs non-septic controls | IL-6 | 7 | 0.54 | 0.25 to 0.82 | 0.65 | 0.57 to 0.72 | 72% |
| vs non-septic controls | IL-8 | 4 | 0.47 | 0.12 to 0.82 | 0.63 | 0.53 to 0.72 | 75% |
| vs non-septic controls | sTNFR-1 | 2 | 0.37 | -0.15 to 0.89 | 0.60 | 0.46 to 0.74 | 79% |
| vs non-septic controls | IL-17 | 3 | 0.37 | 0.01 to 0.73 | 0.60 | 0.50 to 0.70 | 48% |
| vs non-septic controls | IL-1β | 5 | 0.24 | 0.03 to 0.45 | 0.57 | 0.51 to 0.62 | 43% |
| vs non-septic controls | IL-10 | 7 | 0.16 | -0.09 to 0.41 | 0.55 | 0.48 to 0.61 | 66% |
| vs non-septic controls | IFN-γ | 4 | 0.15 | -0.09 to 0.39 | 0.54 | 0.47 to 0.61 | 44% |
| vs non-septic controls | TNF-α | 6 | -0.02 | -0.56 to 0.52 | 0.49 | 0.35 to 0.64 | 91% |
| vs non-septic controls | IL-12 | 4 | -0.08 | -0.46 to 0.30 | 0.48 | 0.37 to 0.58 | 79% |

Implied AUC = $\Phi(\text{SMD}/\sqrt{2})$, applied to the point estimate and both confidence limits. Values are implied, not measured, accuracies.

#### eTable 24. Biomarker prioritization index and its rank stability

| Framework | Biomarker | Evidence quantity | Consistency | Effect magnitude | Association | BPI (base weights) | Rank | Rank range across the five weighting scenarios |
|---|---|---|---|---|---|---|---|---|
| vs septic controls | IL-6 | 100.0 | 83.3 | 72.4 | 100.0 | 88.9 | 1 | 1 |
| vs septic controls | S100B | 66.7 | 87.5 | 95.9 | 0.0 | 62.5 | 2 | 2 |
| vs septic controls | NSE | 41.7 | 80.0 | 100.0 | 0.0 | 55.4 | 3 | 3 |
| vs septic controls | IL-10 | 33.3 | 100.0 | 66.9 | 0.0 | 50.1 | 4 | 4 |
| vs septic controls | TNF-α | 33.3 | 100.0 | 40.0 | 0.0 | 43.3 | 5 | 5 |
| vs septic controls | IFN-γ | 16.7 | 100.0 | 37.9 | 0.0 | 38.6 | 6 | 6 |
| vs septic controls | CRP | 16.7 | 50.0 | 17.2 | 0.0 | 21.0 | 7 | 7 |
| vs non-septic controls | IL-6 | 100.0 | 85.7 | 100.0 | 100.0 | 96.4 | 1 | 1 |
| vs non-septic controls | IL-1β | 71.4 | 80.0 | 44.4 | 100.0 | 74.0 | 2 | 2-6 |
| vs non-septic controls | IL-8 | 57.1 | 100.0 | 87.0 | 0.0 | 61.0 | 3 | 2-3 |
| vs non-septic controls | IL-17 | 42.9 | 100.0 | 68.5 | 0.0 | 52.8 | 4 | 3-5 |
| vs non-septic controls | IL-10 | 100.0 | 71.4 | 29.6 | 0.0 | 50.3 | 5 | 4-6 |
| vs non-septic controls | sTNFR-1 | 28.6 | 100.0 | 68.5 | 0.0 | 49.3 | 6 | 5-7 |
| vs non-septic controls | IFN-γ | 57.1 | 100.0 | 27.8 | 0.0 | 46.2 | 7 | 6-8 |
| vs non-septic controls | TNF-α | 85.7 | 50.0 | 3.7 | 0.0 | 34.9 | 8 | 7-8 |
| vs non-septic controls | IL-12 | 57.1 | 50.0 | 14.8 | 0.0 | 30.5 | 9 | 9 |

Each component is rescaled to 0–100 within its framework; scores are comparable within but not between frameworks. Consistency is the proportion of comparisons pointing to higher concentrations in SAE/SAD (eTable 21). Base weights equal; the four sensitivity scenarios remove the association component, remove the consistency component, and double the weight on effect magnitude and on evidence quantity, respectively.

#### eTable 25. Leave-one-out sensitivity analysis for every pooled comparison with three or more studies

| Framework | Biomarker | Pooled estimate, all studies (95% CI) | Study omitted | k | Estimate without the study (95% CI) | $I^2$, % | Significance changes |
|---|---|---|---|---|---|---|---|
| Non-septic controls | IL-6 | 0.54 (0.25 to 0.82) | Mao 2023 | 6 | 0.52 (0.21 to 0.83) | 76 | No |
| Non-septic controls | IL-6 | 0.54 (0.25 to 0.82) | Orhun 2019 | 6 | 0.46 (0.19 to 0.73) | 64 | No |
| Non-septic controls | IL-6 | 0.54 (0.25 to 0.82) | Brummel 2024 | 6 | 0.62 (0.33 to 0.90) | 52 | No |
| Non-septic controls | IL-6 | 0.54 (0.25 to 0.82) | Jorge-Ripper 2016 | 6 | 0.49 (0.19 to 0.79) | 73 | No |
| Non-septic controls | IL-6 | 0.54 (0.25 to 0.82) | Lei 2022 | 6 | 0.62 (0.32 to 0.92) | 74 | No |
| Non-septic controls | IL-6 | 0.54 (0.25 to 0.82) | Ritter 2014 | 6 | 0.56 (0.23 to 0.89) | 77 | No |
| Non-septic controls | IL-6 | 0.54 (0.25 to 0.82) | Smith 2022 | 6 | 0.49 (0.18 to 0.80) | 71 | No |
| Non-septic controls | IL-8 | 0.47 (0.12 to 0.82) | Mao 2023 | 3 | 0.38 (0.03 to 0.74) | 75 | No |
| Non-septic controls | IL-8 | 0.47 (0.12 to 0.82) | Orhun 2019 | 3 | 0.46 (0.01 to 0.92) | 79 | No |
| Non-septic controls | IL-8 | 0.47 (0.12 to 0.82) | Brummel 2024 | 3 | 0.62 (0.37 to 0.88) | 0 | No |
| Non-septic controls | IL-8 | 0.47 (0.12 to 0.82) | Smith 2022 | 3 | 0.43 (0.00 to 0.86) | 75 | No |
| Non-septic controls | IFN-γ | 0.15 (-0.09 to 0.39) | Mao 2023 | 3 | 0.20 (-0.11 to 0.51) | 63 | No |
| Non-septic controls | IFN-γ | 0.15 (-0.09 to 0.39) | Orhun 2019 | 3 | 0.15 (-0.19 to 0.49) | 56 | No |
| Non-septic controls | IFN-γ | 0.15 (-0.09 to 0.39) | Brummel 2024 | 3 | 0.29 (0.02 to 0.57) | 0 | Yes |
| Non-septic controls | IFN-γ | 0.15 (-0.09 to 0.39) | Jorge-Ripper 2016 | 3 | 0.03 (-0.10 to 0.15) | 0 | No |
| Non-septic controls | IL-12 | -0.08 (-0.46 to 0.30) | Mao 2023 | 3 | -0.11 (-0.58 to 0.35) | 86 | No |
| Non-septic controls | IL-12 | -0.08 (-0.46 to 0.30) | Orhun 2019 | 3 | 0.09 (-0.22 to 0.41) | 61 | No |
| Non-septic controls | IL-12 | -0.08 (-0.46 to 0.30) | Brummel 2024 | 3 | -0.08 (-0.78 to 0.62) | 86 | No |
| Non-septic controls | IL-12 | -0.08 (-0.46 to 0.30) | Smith 2022 | 3 | -0.24 (-0.66 to 0.18) | 75 | No |
| Non-septic controls | TNF-α | -0.02 (-0.56 to 0.52) | Mao 2023 | 5 | -0.00 (-0.63 to 0.63) | 93 | No |
| Non-septic controls | TNF-α | -0.02 (-0.56 to 0.52) | Orhun 2019 | 5 | 0.25 (-0.11 to 0.60) | 73 | No |
| Non-septic controls | TNF-α | -0.02 (-0.56 to 0.52) | Brummel 2024 | 5 | -0.01 (-0.85 to 0.84) | 93 | No |
| Non-septic controls | TNF-α | -0.02 (-0.56 to 0.52) | Jorge-Ripper 2016 | 5 | -0.16 (-0.75 to 0.44) | 91 | No |
| Non-septic controls | TNF-α | -0.02 (-0.56 to 0.52) | Lei 2022 | 5 | -0.17 (-0.75 to 0.41) | 91 | No |
| Non-septic controls | TNF-α | -0.02 (-0.56 to 0.52) | Ritter 2014 | 5 | -0.02 (-0.69 to 0.64) | 93 | No |
| Non-septic controls | IL-1β | 0.24 (0.03 to 0.45) | Mao 2023 | 4 | 0.28 (0.04 to 0.52) | 54 | No |
| Non-septic controls | IL-1β | 0.24 (0.03 to 0.45) | Orhun 2019 | 4 | 0.26 (-0.02 to 0.53) | 57 | Yes |
| Non-septic controls | IL-1β | 0.24 (0.03 to 0.45) | Brummel 2024 | 4 | 0.31 (0.00 to 0.61) | 45 | No |
| Non-septic controls | IL-1β | 0.24 (0.03 to 0.45) | Ritter 2014 | 4 | 0.15 (0.03 to 0.27) | 0 | No |
| Non-septic controls | IL-1β | 0.24 (0.03 to 0.45) | Smith 2022 | 4 | 0.27 (-0.02 to 0.55) | 57 | Yes |
| Non-septic controls | IL-10 | 0.16 (-0.09 to 0.41) | Jorge-Ripper 2016 | 6 | 0.14 (-0.15 to 0.42) | 69 | No |
| Non-septic controls | IL-10 | 0.16 (-0.09 to 0.41) | Mao 2023 | 6 | 0.18 (-0.10 to 0.46) | 71 | No |
| Non-septic controls | IL-10 | 0.16 (-0.09 to 0.41) | Orhun 2019 | 6 | 0.12 (-0.16 to 0.40) | 67 | No |
| Non-septic controls | IL-10 | 0.16 (-0.09 to 0.41) | Brummel 2024 | 6 | 0.20 (-0.13 to 0.53) | 65 | No |
| Non-septic controls | IL-10 | 0.16 (-0.09 to 0.41) | Lei 2022 | 6 | 0.16 (-0.12 to 0.44) | 71 | No |
| Non-septic controls | IL-10 | 0.16 (-0.09 to 0.41) | Ritter 2014 | 6 | 0.25 (0.02 to 0.49) | 56 | Yes |
| Non-septic controls | IL-10 | 0.16 (-0.09 to 0.41) | Smith 2022 | 6 | 0.08 (-0.15 to 0.30) | 49 | No |
| Non-septic controls | IL-17 | 0.37 (0.01 to 0.73) | Mao 2023 | 2 | 0.48 (0.10 to 0.87) | 47 | No |
| Non-septic controls | IL-17 | 0.37 (0.01 to 0.73) | Orhun 2019 | 2 | 0.38 (-0.26 to 1.03) | 70 | Yes |
| Non-septic controls | IL-17 | 0.37 (0.01 to 0.73) | Smith 2022 | 2 | 0.20 (-0.14 to 0.53) | 0 | Yes |
| Septic controls | IL-6 | 1.05 (0.51 to 1.59) | Feng 2017 | 11 | 1.10 (0.51 to 1.69) | 92 | No |

| Framework | Biomarker | Pooled estimate, all studies (95% CI) | Study omitted | k | Estimate without the study (95% CI) | I², % | Significance changes |
|---|---|---|---|---|---|---|---|
| Septic controls | IL-6 | 1.05 (0.51 to 1.59) | Guo 2022 | 11 | 1.01 (0.42 to 1.61) | 92 | No |
| Septic controls | IL-6 | 1.05 (0.51 to 1.59) | Li 2022 | 11 | 0.92 (0.37 to 1.46) | 92 | No |
| Septic controls | IL-6 | 1.05 (0.51 to 1.59) | Nakamura 2003 | 11 | 0.92 (0.37 to 1.47) | 92 | No |
| Septic controls | IL-6 | 1.05 (0.51 to 1.59) | Pfister 2008 | 11 | 1.09 (0.52 to 1.65) | 93 | No |
| Septic controls | IL-6 | 1.05 (0.51 to 1.59) | Wang 2022 | 11 | 1.09 (0.51 to 1.67) | 92 | No |
| Septic controls | IL-6 | 1.05 (0.51 to 1.59) | Wei 2025 | 11 | 0.90 (0.44 to 1.36) | 86 | No |
| Septic controls | IL-6 | 1.05 (0.51 to 1.59) | Wu 2020 | 11 | 1.10 (0.50 to 1.70) | 92 | No |
| Septic controls | IL-6 | 1.05 (0.51 to 1.59) | Yu 2025 | 11 | 1.05 (0.43 to 1.68) | 93 | No |
| Septic controls | IL-6 | 1.05 (0.51 to 1.59) | Erikson 2018 | 11 | 1.10 (0.53 to 1.67) | 92 | No |
| Septic controls | IL-6 | 1.05 (0.51 to 1.59) | Jorge-Ripper 2016 | 11 | 1.15 (0.61 to 1.70) | 91 | No |
| Septic controls | IL-6 | 1.05 (0.51 to 1.59) | Lei 2022 | 11 | 1.18 (0.64 to 1.72) | 91 | No |
| Septic controls | IL-10 | 0.97 (0.05 to 1.88) | Li 2022 | 3 | 0.37 (-0.10 to 0.85) | 71 | Yes |
| Septic controls | IL-10 | 0.97 (0.05 to 1.88) | Yu 2025 | 3 | 1.10 (-0.42 to 2.61) | 94 | Yes |
| Septic controls | IL-10 | 0.97 (0.05 to 1.88) | Jorge-Ripper 2016 | 3 | 1.28 (-0.07 to 2.63) | 94 | Yes |
| Septic controls | IL-10 | 0.97 (0.05 to 1.88) | Lei 2022 | 3 | 1.31 (0.13 to 2.49) | 94 | No |
| Septic controls | NSE | 1.45 (0.33 to 2.57) | Feng 2017 | 4 | 1.83 (0.27 to 3.38) | 94 | No |
| Septic controls | NSE | 1.45 (0.33 to 2.57) | Guo 2022 | 4 | 1.75 (0.07 to 3.42) | 95 | No |
| Septic controls | NSE | 1.45 (0.33 to 2.57) | Li 2022 | 4 | 0.59 (0.24 to 0.93) | 30 | No |
| Septic controls | NSE | 1.45 (0.33 to 2.57) | Wang 2022 | 4 | 1.74 (0.26 to 3.22) | 95 | No |
| Septic controls | NSE | 1.45 (0.33 to 2.57) | Erikson 2018 | 4 | 1.86 (0.54 to 3.19) | 94 | No |
| Septic controls | S100B | 1.39 (0.54 to 2.25) | Feng 2017 | 7 | 1.65 (0.67 to 2.63) | 94 | No |
| Septic controls | S100B | 1.39 (0.54 to 2.25) | Guo 2022 | 7 | 1.28 (0.39 to 2.18) | 92 | No |
| Septic controls | S100B | 1.39 (0.54 to 2.25) | Li 2022 | 7 | 0.75 (0.18 to 1.32) | 86 | No |
| Septic controls | S100B | 1.39 (0.54 to 2.25) | Pfister 2008 | 7 | 1.51 (0.58 to 2.44) | 94 | No |
| Septic controls | S100B | 1.39 (0.54 to 2.25) | Wang 2022 | 7 | 1.53 (0.55 to 2.51) | 94 | No |
| Septic controls | S100B | 1.39 (0.54 to 2.25) | Wu 2020 | 7 | 1.60 (0.53 to 2.67) | 94 | No |
| Septic controls | S100B | 1.39 (0.54 to 2.25) | Erikson 2018 | 7 | 1.52 (0.57 to 2.47) | 94 | No |
| Septic controls | S100B | 1.39 (0.54 to 2.25) | Jorge-Ripper 2016 | 7 | 1.66 (0.70 to 2.63) | 93 | No |
| Septic controls | TNF-α | 0.58 (0.08 to 1.08) | Li 2022 | 3 | 0.32 (-0.00 to 0.65) | 0 | Yes |
| Septic controls | TNF-α | 0.58 (0.08 to 1.08) | Erikson 2018 | 3 | 0.65 (0.01 to 1.28) | 74 | No |
| Septic controls | TNF-α | 0.58 (0.08 to 1.08) | Jorge-Ripper 2016 | 3 | 0.76 (0.16 to 1.35) | 54 | No |
| Septic controls | TNF-α | 0.58 (0.08 to 1.08) | Lei 2022 | 3 | 0.62 (-0.11 to 1.36) | 74 | Yes |

Each study was omitted in turn and the random-effects model refitted. "Significance changes" marks omissions after which the 95% CI crosses, or ceases to cross, the null. Comparisons with two studies (sTNFR-1 against non-septic controls; CRP and IFN-γ against septic controls) are not eligible.

#### eTable 26. Sensitivity analyses excluding studies at high risk of bias and cerebrospinal fluid measurements

| Analysis | Framework | Biomarker | Studies excluded | Pooled estimate, all studies (95% CI) | k | Estimate after exclusion (95% CI) | I², % | Significance changes |
|---|---|---|---|---|---|---|---|---|
| Excluding high risk of bias | Non-septic controls | IL-6 | Mao 2023 | 0.54 (0.25 to 0.82) | 6 | 0.52 (0.21 to 0.83) | 76 | No |
| Excluding high risk of bias | Non-septic controls | IL-8 | Mao 2023 | 0.47 (0.12 to 0.82) | 3 | 0.38 (0.03 to 0.74) | 75 | No |
| Excluding high risk of bias | Non-septic controls | IFN-γ | Mao 2023 | 0.15 (-0.09 to 0.39) | 3 | 0.20 (-0.11 to 0.51) | 63 | No |
| Excluding high risk of bias | Non-septic controls | IL-12 | Mao 2023 | -0.08 (-0.46 to 0.30) | 3 | -0.11 (-0.58 to 0.35) | 86 | No |
| Excluding high risk of bias | Non-septic controls | TNF-α | Mao 2023 | -0.02 (-0.56 to 0.52) | 5 | -0.00 (-0.63 to 0.63) | 93 | No |
| Excluding high risk of bias | Non-septic controls | IL-1β | Mao 2023 | 0.24 (0.03 to 0.45) | 4 | 0.28 (0.04 to 0.52) | 54 | No |
| Excluding high risk of bias | Non-septic controls | IL-10 | Mao 2023 | 0.16 (-0.09 to 0.41) | 6 | 0.18 (-0.10 to 0.46) | 71 | No |
| Excluding high risk of bias | Non-septic controls | IL-17 | Mao 2023 | 0.37 (0.01 to 0.73) | 2 | 0.48 (0.10 to 0.87) | 47 | No |
| Excluding high risk of bias | Septic controls | IL-6 | Feng 2017 | 1.05 (0.51 to 1.59) | 11 | 1.10 (0.51 to 1.69) | 92 | No |
| Excluding high risk of bias | Septic controls | NSE | Feng 2017 | 1.45 (0.33 to 2.57) | 4 | 1.83 (0.27 to 3.38) | 94 | No |
| Excluding high risk of bias | Septic controls | S100B | Feng 2017 | 1.39 (0.54 to 2.25) | 7 | 1.65 (0.67 to 2.63) | 94 | No |
| Blood only (excluding cerebrospinal fluid) | Non-septic controls | IL-6 | Mao 2023 | 0.54 (0.25 to 0.82) | 6 | 0.52 (0.21 to 0.83) | 76 | No |
| Blood only (excluding cerebrospinal fluid) | Non-septic controls | IL-8 | Mao 2023 | 0.47 (0.12 to 0.82) | 3 | 0.38 (0.03 to 0.74) | 75 | No |
| Blood only (excluding cerebrospinal fluid) | Non-septic controls | IFN-γ | Mao 2023 | 0.15 (-0.09 to 0.39) | 3 | 0.20 (-0.11 to 0.51) | 63 | No |
| Blood only (excluding cerebrospinal fluid) | Non-septic controls | IL-12 | Mao 2023 | -0.08 (-0.46 to 0.30) | 3 | -0.11 (-0.58 to 0.35) | 86 | No |
| Blood only (excluding cerebrospinal fluid) | Non-septic controls | TNF-α | Mao 2023 | -0.02 (-0.56 to 0.52) | 5 | -0.00 (-0.63 to 0.63) | 93 | No |
| Blood only (excluding cerebrospinal fluid) | Non-septic controls | IL-1β | Mao 2023 | 0.24 (0.03 to 0.45) | 4 | 0.28 (0.04 to 0.52) | 54 | No |
| Blood only (excluding cerebrospinal fluid) | Non-septic controls | IL-10 | Mao 2023 | 0.16 (-0.09 to 0.41) | 6 | 0.18 (-0.10 to 0.46) | 71 | No |
| Blood only (excluding cerebrospinal fluid) | Non-septic controls | IL-17 | Mao 2023 | 0.37 (0.01 to 0.73) | 2 | 0.48 (0.10 to 0.87) | 47 | No |

Studies at high risk of bias on the Newcastle-Ottawa Scale (eTable 2): Feng 2017 and Mao 2023. Mao 2023 is also the only study reporting cerebrospinal fluid concentrations, so the prespecified compartment analysis coincides with its exclusion. No pooled estimate changed significance.

#### eFigure 1A. Forest plot versus non-septic controls — IL-6

| | SAE/Delirium | | | Control (non-sepsis) | | | | |
|---|---|---|---|---|---|---|---|---|
| **Study or Subgroup** | **Mean** | **SD** | **Total** | **Mean** | **SD** | **Total** | **Weight** | **Std. Mean Difference IV, Random, 95% CI** |
| **2.1.1 SAE** | | | | | | | | |
| Mao 2023 | 9.67 | 14.21 | 19 | 3.15 | 2.65 | 24 | 10.7% | 0.67 [0.05, 1.28] |
| Orhun 2019 | 54.22 | 66.32 | 82 | 0 | 0.02 | 33 | 14.7% | 0.96 [0.54, 1.38] |
| **Subtotal (95% CI)** | | | **101** | | | **57** | | **0.87 [0.52, 1.22]** |
| **2.1.2 Delirium only** | | | | | | | | |
| Brummel 2024 | 3.56 | 1.6 | 691 | 3.17 | 1.68 | 300 | 20.8% | 0.24 [0.11, 0.38] |
| Jorge-Ripper 2016 | 31.5 | 42.51 | 47 | 2.9 | 6.75 | 27 | 13.2% | 0.83 [0.33, 1.32] |
| Lei 2022 | 43.17 | 51.98 | 17 | 50.13 | 74.4 | 35 | 11.4% | -0.10 [-0.68, 0.48] |
| Ritter 2014 | 332.33 | 97.77 | 31 | 237.33 | 288.88 | 47 | 13.9% | 0.40 [-0.05, 0.86] |
| Smith 2022 | 426.84 | 418.61 | 41 | 181.89 | 238.74 | 78 | 15.4% | 0.78 [0.39, 1.17] |
| **Subtotal (95% CI)** | | | **827** | | | **487** | | **0.43 [0.13, 0.73]** |
| **Total (95% CI)** | | | **928** | | | **544** | **100.0%** | **0.54 [0.25, 0.82]** |

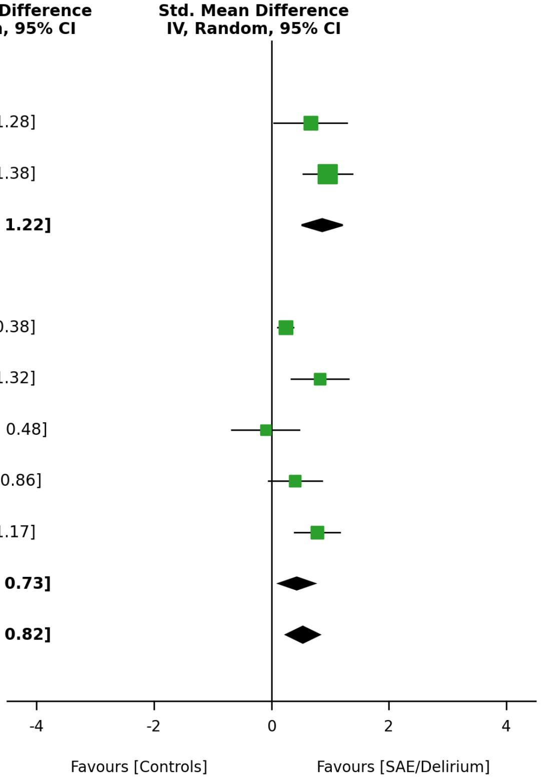


Heterogeneity: $Tau^2 = 0.10$; $Chi^2 = 21.77$, df = 6; $I^2 = 72\%$

Test for overall effect: Z = 3.70 (P < 0.001)

Test for subgroup differences: $Chi^2 = 3.40$, df = 1 (P = 0.07)

Forest plots of every biomarker pooled against non-septic controls. Random-effects standardized mean differences (inverse variance, DerSimonian-Laird) with 95% CIs, one panel per biomarker, with subtotals by neurological phenotype (SAE stratum, delirium-only stratum) where both strata contribute. Erikson 2018 does not appear in this framework: its only comparison is delirium against no delirium within sepsis, which belongs to the septic-control framework (eTable 18). Arm-level values are in eTable 16.

#### eFigure 1B. Forest plot versus non-septic controls — IL-1β

| Study or Subgroup | SAE/Delirium Mean | SAE/Delirium SD | SAE/Delirium Total | Control (non-sepsis) Mean | Control (non-sepsis) SD | Control (non-sepsis) Total | Weight | Std. Mean Difference IV, Random, 95% CI |
|---|---|---|---|---|---|---|---|---|
| **2.2.1 SAE** | | | | | | | | |
| Mao 2023 | 2.9 | 3.23 | 19 | 3 | 5.73 | 24 | 9.7% | -0.02 [-0.62, 0.58] |
| Orhun 2019 | 15.63 | 78.09 | 82 | 0 | 0.1 | 33 | 17.2% | 0.23 [-0.17, 0.64] |
| **Subtotal (95% CI)** | | | **101** | | | **57** | | **0.16 [-0.18, 0.49]** |
| **2.2.2 Delirium only** | | | | | | | | |
| Brummel 2024 | -0.85 | 1.06 | 691 | -0.987 | 0.756 | 300 | 40.3% | 0.14 [0.00, 0.28] |
| Ritter 2014 | 76.33 | 82.22 | 31 | 34.33 | 17.78 | 47 | 14.1% | 0.78 [0.31, 1.25] |
| Smith 2022 | 9.81 | 11.65 | 41 | 7.4 | 12.67 | 78 | 18.7% | 0.19 [-0.18, 0.57] |
| **Subtotal (95% CI)** | | | **763** | | | **425** | | **0.31 [-0.02, 0.65]** |
| **Total (95% CI)** | | | **864** | | | **482** | **100.0%** | **0.24 [0.03, 0.45]** |

Heterogeneity: $Tau^2 = 0.02$; $Chi^2 = 7.03$, df = 4; $I^2 = 43\%$

Test for overall effect: Z = 2.25 (P = 0.024)

Test for subgroup differences: $Chi^2 = 0.43$, df = 1 (P = 0.51)

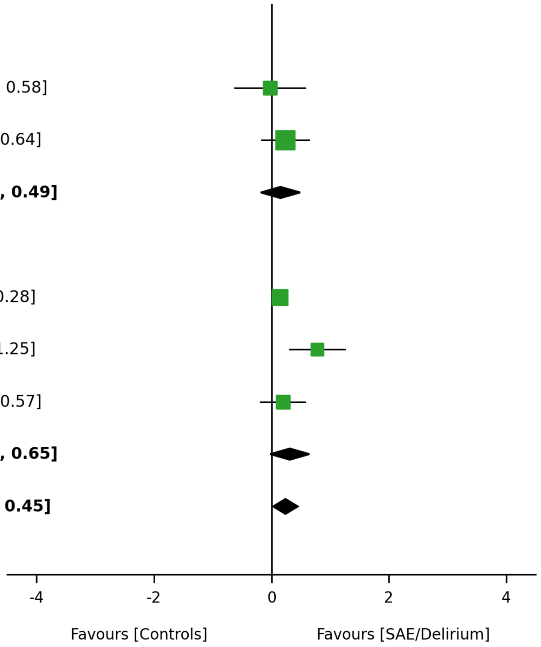

#### eFigure 1C. Forest plot versus non-septic controls — IL-10

| Study or Subgroup | SAE/Delirium Mean | SAE/Delirium SD | SAE/Delirium Total | Control (non-sepsis) Mean | Control (non-sepsis) SD | Control (non-sepsis) Total | Weight | Std. Mean Difference IV, Random, 95% CI |
|---|---|---|---|---|---|---|---|---|
| **2.3.1 SAE** | | | | | | | | |
| Mao 2023 | 2.57 | 0.349 | 19 | 2.57 | 0.294 | 24 | 10.1% | 0.00 [-0.60, 0.60] |
| Orhun 2019 | 12.1 | 35.2 | 82 | 0 | 0.01 | 33 | 14.8% | 0.40 [-0.00, 0.81] |
| **Subtotal (95% CI)** | | | **101** | | | **57** | | **0.26 [-0.11, 0.64]** |
| **2.3.2 Delirium only** | | | | | | | | |
| Jorge-Ripper 2016 | 9.9 | 35.65 | 47 | 0 | 0 | 27 | 12.9% | 0.34 [-0.13, 0.82] |
| Brummel 2024 | 2.26 | 1.26 | 691 | 2.23 | 1.41 | 300 | 22.7% | 0.02 [-0.12, 0.16] |
| Lei 2022 | 23.97 | 27.8 | 17 | 18.67 | 23.29 | 35 | 10.6% | 0.21 [-0.37, 0.79] |
| Ritter 2014 | 5.77 | 1.11 | 31 | 24.77 | 51.11 | 47 | 13.4% | -0.47 [-0.93, -0.01] |
| Smith 2022 | 50.79 | 63.46 | 41 | 21.99 | 35.01 | 78 | 15.4% | 0.61 [0.23, 1.00] |
| **Subtotal (95% CI)** | | | **827** | | | **487** | | **0.14 [-0.18, 0.46]** |
| **Total (95% CI)** | | | **928** | | | **544** | **100.0%** | **0.16 [-0.09, 0.41]** |

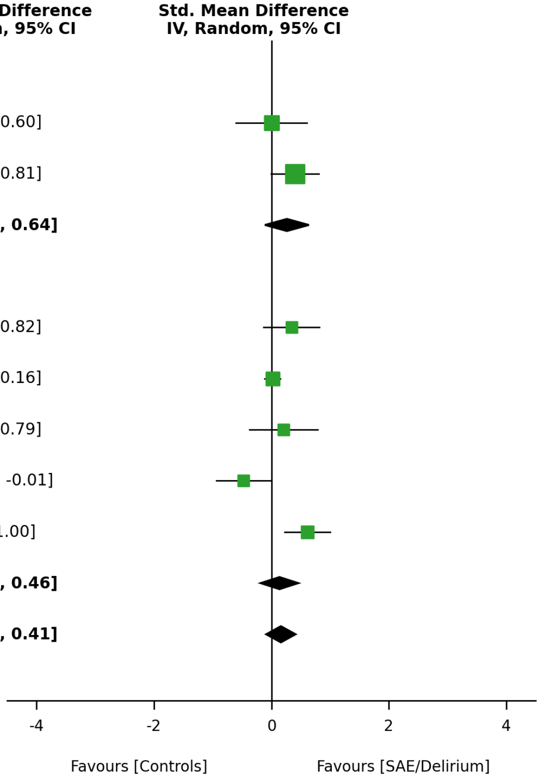


Heterogeneity: $Tau^2 = 0.07$; $Chi^2 = 17.41$, df = 6; $I^2 = 66\%$

Test for overall effect: Z = 1.27 (P = 0.205)

Test for subgroup differences: $Chi^2 = 0.25$, df = 1 (P = 0.62)

#### eFigure 1D. Forest plot versus non-septic controls — TNF-α

| | SAE/Delirium | | | Control (non-sepsis) | | | | |
|---|---|---|---|---|---|---|---|---|
| **Study or Subgroup** | **Mean** | **SD** | **Total** | **Mean** | **SD** | **Total** | **Weight** | **Std. Mean Difference IV, Random, 95% CI** |
| **2.4.1 SAE** | | | | | | | | |
| Mao 2023 | 2.6 | 0.697 | 19 | 2.68 | 0.931 | 24 | 15.4% | -0.09 [-0.70, 0.51] |
| Orhun 2019 | 37.92 | 51.62 | 82 | 105.04 | 27.5 | 33 | 16.9% | -1.45 [-1.89, -1.00] |
| **Subtotal (95% CI)** | | | **101** | | | **57** | | **-0.79 [-2.11, 0.54]** |
| **2.4.2 Delirium only** | | | | | | | | |
| Brummel 2024 | 2.09 | 0.719 | 691 | 2.12 | 0.77 | 300 | 18.9% | -0.03 [-0.17, 0.10] |
| Jorge-Ripper 2016 | 18 | 28.11 | 47 | 2.17 | 4.16 | 27 | 16.5% | 0.69 [0.21, 1.18] |
| Lei 2022 | 14.17 | 6 | 17 | 10.89 | 2.37 | 35 | 15.4% | 0.82 [0.22, 1.43] |
| Ritter 2014 | 225.33 | 382.96 | 31 | 214.67 | 298.52 | 47 | 16.8% | 0.03 [-0.42, 0.49] |
| **Subtotal (95% CI)** | | | **786** | | | **409** | | **0.32 [-0.10, 0.75]** |
| **Total (95% CI)** | | | **887** | | | **466** | **100.0%** | **-0.02 [-0.56, 0.52]** |

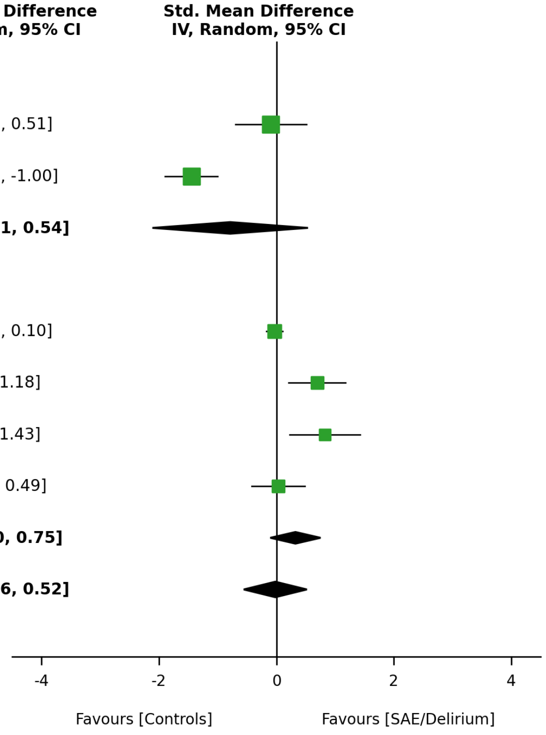


Heterogeneity: $Tau^2 = 0.39$; $Chi^2 = 54.82$, df = 5; $I^2 = 91\%$

Test for overall effect: Z = 0.06 (P = 0.948)

Test for subgroup differences: $Chi^2 = 2.44$, df = 1 (P = 0.12)

### eFigure 1E. Forest plot versus non-septic controls — IFN-γ

| | SAE/Delirium | | | Control (non-sepsis) | | | | |
|---|---|---|---|---|---|---|---|---|
| **Study or Subgroup** | **Mean** | **SD** | **Total** | **Mean** | **SD** | **Total** | **Weight** | **Std. Mean Difference IV, Random, 95% CI** |
| **2.5.1 SAE** | | | | | | | | |
| Mao 2023 | 3.24 | 9.76 | 19 | 3.05 | 7.84 | 24 | 12.6% | 0.02 [-0.58, 0.62] |
| Orhun 2019 | 7.25 | 18.02 | 82 | 3.65 | 0.91 | 33 | 21.9% | 0.23 [-0.17, 0.64] |
| **Subtotal (95% CI)** | | | **101** | | | **57** | | **0.17 [-0.17, 0.50]** |
| **2.5.2 Delirium only** | | | | | | | | |
| Brummel 2024 | 0.0767 | 1.34 | 691 | 0.07 | 1.41 | 300 | 48.1% | 0.00 [-0.13, 0.14] |
| Jorge-Ripper 2016 | 22.8 | 43.19 | 47 | 3.45 | 5.98 | 27 | 17.4% | 0.55 [0.07, 1.03] |
| **Subtotal (95% CI)** | | | **738** | | | **327** | | **0.23 [-0.30, 0.75]** |
| **Total (95% CI)** | | | **839** | | | **384** | **100.0%** | **0.15 [-0.09, 0.39]** |

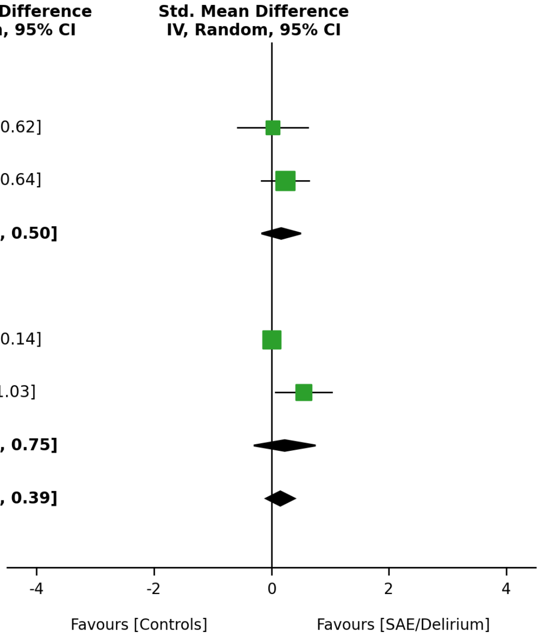


Heterogeneity: Tau² = 0.03; Chi² = 5.36, df = 3; I² = 44%

Test for overall effect: Z = 1.24 (P = 0.217)

Test for subgroup differences: Chi² = 0.03, df = 1 (P = 0.85)

#### eFigure 1F. Forest plot versus non-septic controls — IL-17

| | SAE/Delirium | | | Control (non-sepsis) | | | | | |
|---|---|---|---|---|---|---|---|---|---|
| **Study or Subgroup** | **Mean** | **SD** | **Total** | **Mean** | **SD** | **Total** | **Weight** | **Std. Mean Difference IV, Random, 95% CI** | **Std. Mean Difference IV, Random, 95% CI** |
| **2.6.1 SAE** | | | | | | | | | |
| Mao 2023 | 12 | 13.08 | 19 | 11.87 | 13.03 | 24 | 24.0% | 0.01 [-0.59, 0.61] | |
| Orhun 2019 | 5.03 | 18.33 | 82 | 0.62 | 0.155 | 33 | 37.3% | 0.28 [-0.12, 0.69] | |
| **Subtotal (95% CI)** | | | **101** | | | **57** | | **0.20 [-0.14, 0.53]** | |
| **2.6.2 Delirium only** | | | | | | | | | |
| Smith 2022 (e) | 0.31 | 0.689 | 41 | 0.0333 | 0.0741 | 78 | 38.8% | 0.67 [0.29, 1.06] | |
| **Subtotal (95% CI)** | | | **41** | | | **78** | | **0.67 [0.29, 1.06]** | |
| **Total (95% CI)** | | | **142** | | | **135** | **100.0%** | **0.37 [0.01, 0.73]** | |

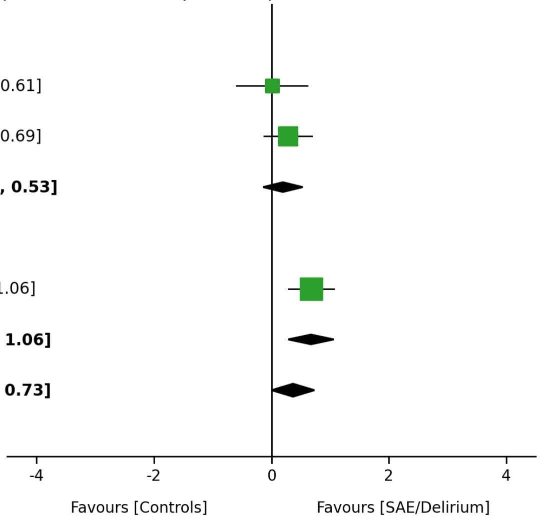


Heterogeneity: $Tau^2 = 0.05$; $Chi^2 = 3.87$, df = 2; $I^2 = 48\%$

Test for overall effect: Z = 1.99 (P = 0.047)

Test for subgroup differences: $Chi^2 = 3.33$, df = 1 (P = 0.07)

(e) Smith 2022: control group reported as 0 (0–0); an SD of 0.074 was imputed to allow estimation.

#### eFigure 1G. Forest plot versus non-septic controls — IL-12

| | SAE/Delirium | | | Control (non-sepsis) | | | | |
|---|---|---|---|---|---|---|---|---|
| **Study or Subgroup** | **Mean** | **SD** | **Total** | **Mean** | **SD** | **Total** | **Weight** | **Std. Mean Difference IV, Random, 95% CI** |
| **2.7.1 SAE** | | | | | | | | |
| Mao 2023 | 2.6 | 0.741 | 19 | 2.56 | 0.539 | 24 | 18.2% | 0.06 [-0.54, 0.66] |
| Orhun 2019 | 10.75 | 9.77 | 82 | 16.62 | 4.16 | 33 | 24.1% | -0.68 [-1.09, -0.27] |
| **Subtotal (95% CI)** | | | **101** | | | **57** | | **-0.34 [-1.07, 0.38]** |
| **2.7.2 Delirium only** | | | | | | | | |
| Brummel 2024 | 0.15 | 1.5 | 691 | 0.263 | 1.59 | 300 | 32.5% | -0.07 [-0.21, 0.06] |
| Smith 2022 | 0.137 | 0.304 | 41 | 0.0567 | 0.126 | 78 | 25.2% | 0.39 [0.01, 0.77] |
| **Subtotal (95% CI)** | | | **732** | | | **378** | | **0.12 [-0.33, 0.57]** |
| **Total (95% CI)** | | | **833** | | | **435** | **100.0%** | **-0.08 [-0.46, 0.30]** |

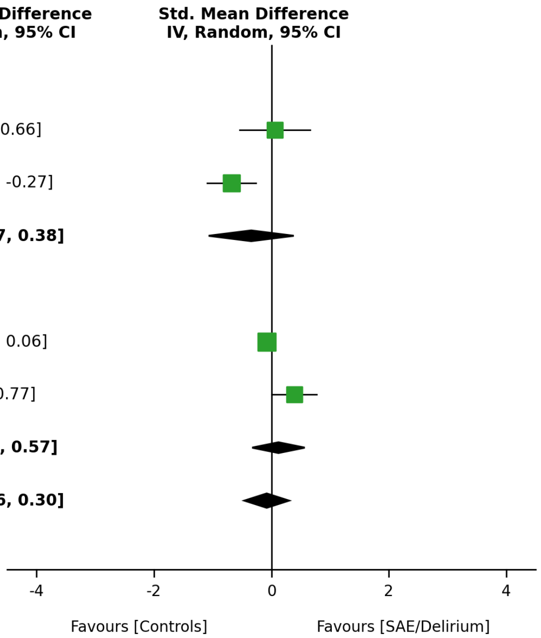


Heterogeneity: Tau² = 0.11; Chi² = 14.10, df = 3; I² = 79%

Test for overall effect: Z = 0.41 (P = 0.682)

Test for subgroup differences: Chi² = 1.14, df = 1 (P = 0.29)

#### eFigure 1H. Forest plot versus non-septic controls — CRP

| | SAE/Delirium | | | Control (non-sepsis) | | | | |
|---|---|---|---|---|---|---|---|---|
| **Study or Subgroup** | **Mean** | **SD** | **Total** | **Mean** | **SD** | **Total** | **Weight** | **Std. Mean Difference IV, Random, 95% CI** |
| **2.8.1 SAE** | | | | | | | | |
| *Not estimable (no comparison)* | | | | | | | | |
| **2.8.2 Delirium only** | | | | | | | | |
| Brummel 2024 | 11.66 | 0.719 | 691 | 11.55 | 1.01 | 300 | 100.0% | 0.13 [-0.01, 0.27] |
| **Subtotal (95% CI)** | | | **691** | | | **300** | | **0.13 [-0.01, 0.27]** |
| **Total (95% CI)** | | | **691** | | | **300** | **100.0%** | **0.13 [-0.01, 0.27]** |

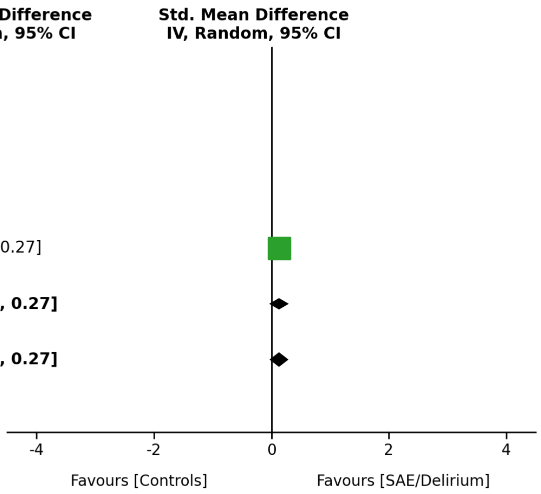


Heterogeneity: Not applicable

Test for overall effect: Z = 1.88 (P = 0.060)

Test for subgroup differences: Not applicable

#### eFigure 1I. Forest plot versus non-septic controls — sTNFR-1

| | SAE/Delirium | | | Control (non-sepsis) | | | | |
|---|---|---|---|---|---|---|---|---|
| **Study or Subgroup** | **Mean** | **SD** | **Total** | **Mean** | **SD** | **Total** | **Weight** | **Std. Mean Difference IV, Random, 95% CI** |
| **2.9.1 SAE** | | | | | | | | |
| *Not estimable (no comparison)* | | | | | | | | |
| **2.9.2 Delirium only** | | | | | | | | |
| Brummel 2024 | 8.99 | 1.04 | 691 | 8.84 | 0.955 | 300 | 58.8% | 0.15 [0.01, 0.28] |
| Ritter 2014 | 4306 | 2,171 | 31 | 2839 | 2,061 | 47 | 41.2% | 0.69 [0.22, 1.16] |
| **Subtotal (95% CI)** | | | **722** | | | **347** | | **0.37 [-0.15, 0.89]** |
| **Total (95% CI)** | | | **722** | | | **347** | **100.0%** | **0.37 [-0.15, 0.89]** |

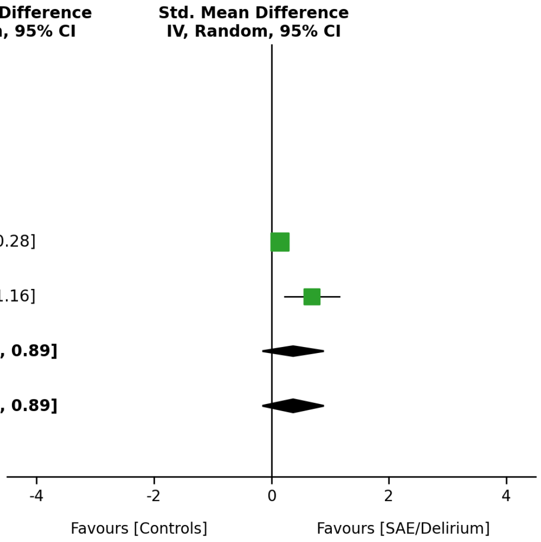


Heterogeneity: $Tau^2 = 0.12$; $Chi^2 = 4.79$, df = 1; $I^2 = 79\%$

Test for overall effect: Z = 1.39 (P = 0.165)

Test for subgroup differences: Not applicable

#### eFigure 1J. Forest plot versus non-septic controls — IL-8

| | SAE/Delirium | | | Control (non-sepsis) | | | | |
|---|---|---|---|---|---|---|---|---|
| **Study or Subgroup** | **Mean** | **SD** | **Total** | **Mean** | **SD** | **Total** | **Weight** | **Std. Mean Difference IV, Random, 95% CI** |
| **2.10.1 SAE** | | | | | | | | |
| Mao 2023 | 203.91 | 290.33 | 19 | 34.22 | 22.65 | 24 | 16.6% | 0.86 [0.23, 1.49] |
| Orhun 2019 | 152.81 | 333.83 | 82 | 0 | 0.05 | 33 | 24.1% | 0.54 [0.13, 0.95] |
| **Subtotal (95% CI)** | | | **101** | | | **57** | | **0.63 [0.29, 0.98]** |
| **2.10.2 Delirium only** | | | | | | | | |
| Brummel 2024 | 1.94 | 0.941 | 691 | 1.81 | 1.01 | 300 | 34.1% | 0.13 [-0.01, 0.26] |
| Smith 2022 | 53.35 | 50.6 | 41 | 32.3 | 21.53 | 78 | 25.1% | 0.61 [0.22, 1.00] |
| **Subtotal (95% CI)** | | | **732** | | | **378** | | **0.33 [-0.13, 0.80]** |
| **Total (95% CI)** | | | **833** | | | **435** | **100.0%** | **0.47 [0.12, 0.82]** |

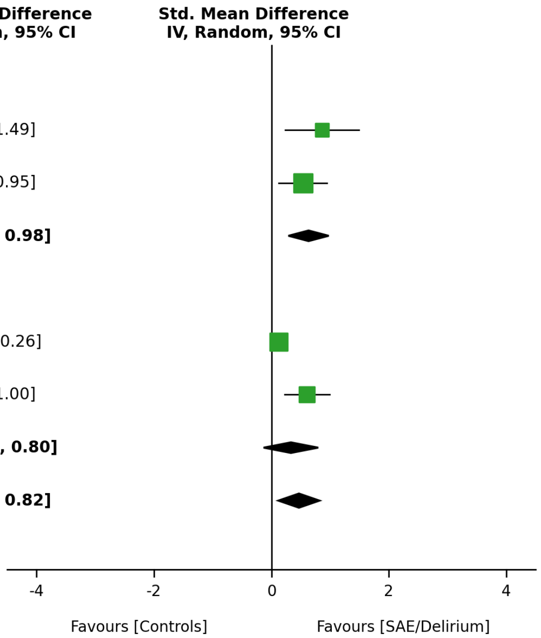


Heterogeneity: Tau² = 0.09; Chi² = 11.86, df = 3; I² = 75%

Test for overall effect: Z = 2.62 (P = 0.009)

Test for subgroup differences: Chi² = 1.03, df = 1 (P = 0.31)

#### eFigure 2A. Forest plot versus septic controls without encephalopathy — IL-6

| | SAE/Delirium | | | Control (within sepsis) | | | | | |
|---|---|---|---|---|---|---|---|---|---|
| **Study or Subgroup** | **Mean** | **SD** | **Total** | **Mean** | **SD** | **Total** | **Weight** | **Std. Mean Difference IV, Random, 95% CI** | **Std. Mean Difference IV, Random, 95% CI** |
| **1.1.1 SAE** | | | | | | | | | |
| Feng 2017 | 1,919 | 3,643 | 36 | 299.86 | 448.84 | 23 | 8.8% | 0.56 [0.02, 1.09] | |
| Guo 2022 | 229.15 | 29.61 | 30 | 196.71 | 18.62 | 90 | 9.0% | 1.48 [1.02, 1.93] | |
| Li 2022 | 237 | 16 | 21 | 191 | 18 | 20 | 7.7% | 2.65 [1.79, 3.51] | |
| Nakamura 2003 | 6050 | 1100 | 16 | 2750 | 1150 | 10 | 6.7% | 2.86 [1.70, 4.01] | |
| Pfister 2008 | 409.63 | 599.18 | 12 | 79.9 | 51.62 | 4 | 6.7% | 0.59 [-0.57, 1.74] | |
| Wang 2022 | 1,999 | 3,545 | 20 | 306.67 | 392.59 | 20 | 8.4% | 0.66 [0.02, 1.30] | |
| Wei 2025 | 928.71 | 163.74 | 105 | 528.71 | 176.51 | 114 | 9.2% | 2.34 [1.99, 2.68] | |
| Wu 2020 | 1876 | 3,635 | 59 | 197.87 | 236.44 | 45 | 9.1% | 0.61 [0.21, 1.00] | |
| Yu 2025 | 13,371 | 17,953 | 62 | 423.03 | 716.13 | 78 | 9.2% | 1.08 [0.72, 1.43] | |
| **Subtotal (95% CI)** | | | **361** | | | **404** | | **1.38 [0.83, 1.94]** | |
| **1.1.2 Delirium only** | | | | | | | | | |
| Erikson 2018 | 250.86 | 293.11 | 10 | 126.1 | 199.03 | 12 | 7.7% | 0.49 [-0.37, 1.34] | |
| Jorge-Ripper 2016 | 31.5 | 42.51 | 47 | 31.7 | 36.68 | 35 | 9.0% | -0.00 [-0.44, 0.43] | |
| Lei 2022 | 43.17 | 51.98 | 17 | 63.83 | 74.46 | 35 | 8.6% | -0.30 [-0.88, 0.28] | |
| **Subtotal (95% CI)** | | | **74** | | | **82** | | **-0.02 [-0.37, 0.33]** | |
| **Total (95% CI)** | | | **435** | | | **486** | **100.0%** | **1.05 [0.51, 1.59]** | |

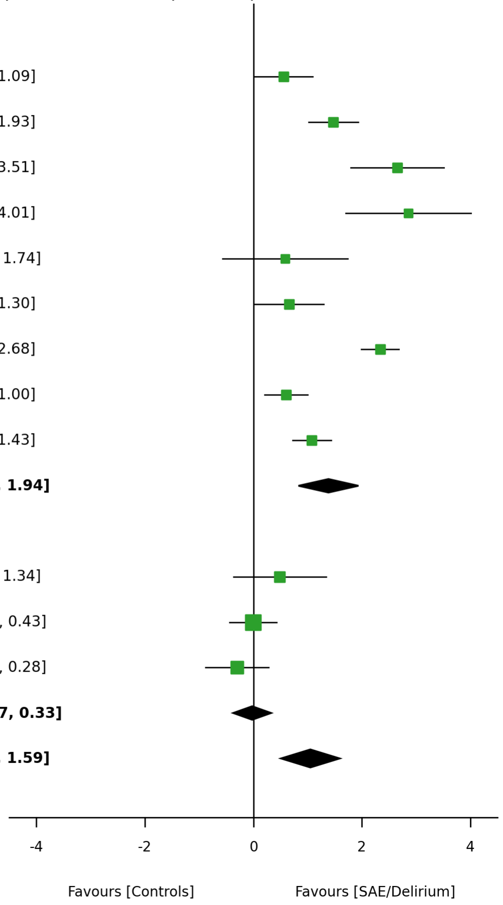


Heterogeneity: Tau² = 0.80; Chi² = 134.32, df = 11; I² = 92%

Test for overall effect: Z = 3.80 (P < 0.001)

Test for subgroup differences: Chi² = 17.72, df = 1 (P = 0.00)

Forest plots of every biomarker pooled against septic patients without encephalopathy. Conventions as in eFigure 1. Index and comparator groups are both septic, so a difference in this framework is attributable to the neurological phenotype rather than to sepsis itself.

#### eFigure 2B. Forest plot versus septic controls without encephalopathy — S100B

| Study or Subgroup | SAE/Delirium Mean | SAE/Delirium SD | SAE/Delirium Total | Control (within sepsis) Mean | Control (within sepsis) SD | Control (within sepsis) Total | Weight | Std. Mean Difference IV, Random, 95% CI |
|---|---|---|---|---|---|---|---|---|
| **1.2.1 SAE** | | | | | | | | |
| Feng 2017 | 0.34 | 0.289 | 36 | 0.307 | 0.274 | 23 | 13.9% | 0.12 [-0.41, 0.64] |
| Guo 2022 | 0.27 | 0.06 | 30 | 0.18 | 0.04 | 90 | 14.0% | 1.96 [1.47, 2.44] |
| Li 2022 | 2.38 | 0.21 | 21 | 0.35 | 0.12 | 20 | 5.9% | 11.56 [8.86, 14.27] |
| Pfister 2008 | 0.276 | 0.318 | 12 | 0.076 | 0.037 | 4 | 11.5% | 0.67 [-0.49, 1.83] |
| Wang 2022 | 892.85 | 383.93 | 20 | 625 | 178.57 | 20 | 13.5% | 0.88 [0.22, 1.53] |
| Wu 2020 | 0.311 | 0.219 | 59 | 0.155 | 0.094 | 45 | 14.2% | 0.88 [0.47, 1.28] |
| **Subtotal (95% CI)** | | | **178** | | | **202** | | **1.87 [0.76, 2.97]** |
| **1.2.2 Delirium only** | | | | | | | | |
| Erikson 2018 | 0.36 | 0.29 | 10 | 0.173 | 0.17 | 12 | 12.7% | 0.78 [-0.10, 1.65] |
| Jorge-Ripper 2016 | 26.3 | 52.1 | 47 | 26.6 | 38.45 | 35 | 14.1% | -0.01 [-0.44, 0.43] |
| **Subtotal (95% CI)** | | | **57** | | | **47** | | **0.29 [-0.45, 1.03]** |
| **Total (95% CI)** | | | **235** | | | **249** | **100.0%** | **1.39 [0.54, 2.25]** |

Heterogeneity: Tau² = 1.29; Chi² = 102.67, df = 7; I² = 93%

Test for overall effect: Z = 3.21 (P = 0.001)

Test for subgroup differences: Chi² = 5.40, df = 1 (P = 0.02)

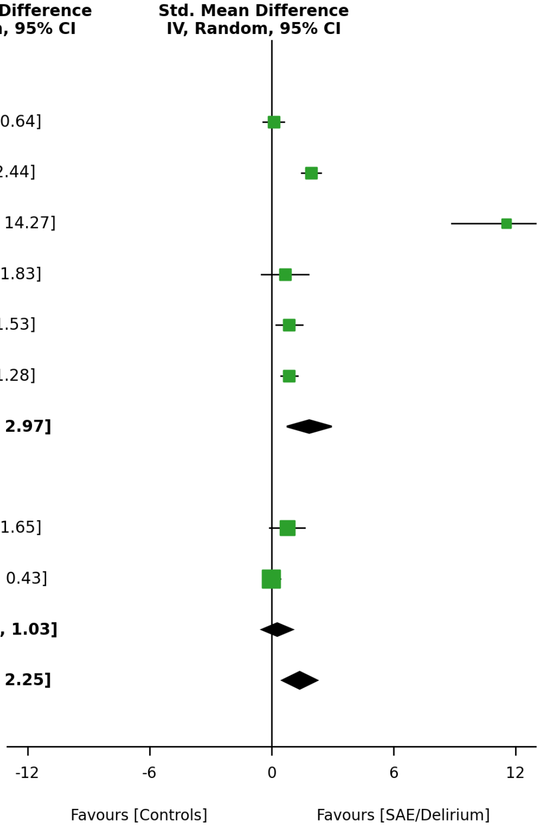

#### eFigure 2C. Forest plot versus septic controls without encephalopathy — TNF-α

| | SAE/Delirium | | | Control (within sepsis) | | | | |
|---|---|---|---|---|---|---|---|---|
| **Study or Subgroup** | **Mean** | **SD** | **Total** | **Mean** | **SD** | **Total** | **Weight** | **Std. Mean Difference IV, Random, 95% CI** |
| **1.3.1 SAE** | | | | | | | | |
| Li 2022 | 17 | 1.6 | 21 | 15.1 | 1.1 | 20 | 23.3% | 1.35 [0.67, 2.04] |
| **Subtotal (95% CI)** | | | **21** | | | **20** | | **1.35 [0.67, 2.04]** |
| **1.3.2 Delirium only** | | | | | | | | |
| Erikson 2018 | 43 | 33.19 | 10 | 33.17 | 17.33 | 12 | 18.9% | 0.37 [-0.48, 1.22] |
| Jorge-Ripper 2016 | 18 | 28.11 | 47 | 12.5 | 24.26 | 35 | 31.5% | 0.21 [-0.23, 0.64] |
| Lei 2022 | 14.17 | 6 | 17 | 12.02 | 2.96 | 35 | 26.4% | 0.51 [-0.08, 1.09] |
| **Subtotal (95% CI)** | | | **74** | | | **82** | | **0.32 [-0.00, 0.65]** |
| **Total (95% CI)** | | | **95** | | | **102** | **100.0%** | **0.58 [0.08, 1.08]** |

Heterogeneity: Tau² = 0.15; Chi² = 7.75, df = 3; I² = 61%

Test for overall effect: Z = 2.29 (P = 0.022)

Test for subgroup differences: Chi² = 7.08, df = 1 (P = 0.01)

**Std. Mean Difference IV, Random, 95% CI**

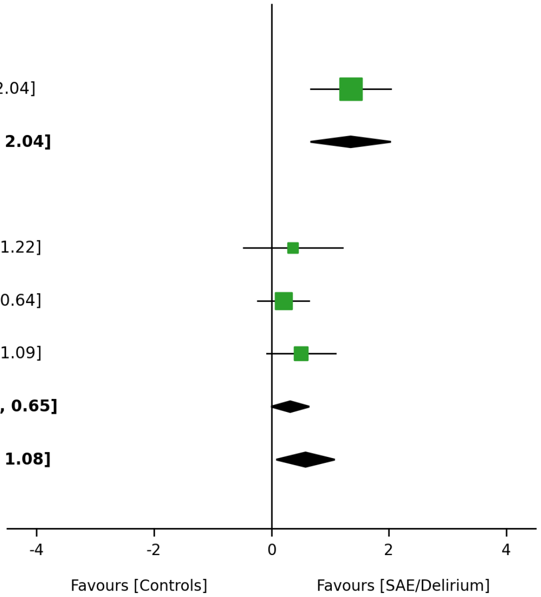

#### eFigure 2D. Forest plot versus septic controls without encephalopathy — IL-10

| | SAE/Delirium | | | Control (within sepsis) | | | | |
|---|---|---|---|---|---|---|---|---|
| **Study or Subgroup** | **Mean** | **SD** | **Total** | **Mean** | **SD** | **Total** | **Weight** | **Std. Mean Difference IV, Random, 95% CI** |
| **1.4.1 SAE** | | | | | | | | |
| Li 2022 | 30.3 | 1.8 | 21 | 25.2 | 1.2 | 20 | 21.4% | 3.25 [2.29, 4.21] |
| Yu 2025 | 1,344 | 2,338 | 62 | 121.44 | 26.3 | 78 | 27.0% | 0.78 [0.44, 1.13] |
| **Subtotal (95% CI)** | | | **83** | | | **98** | | **1.98 [-0.45, 4.40]** |
| **1.4.2 Delirium only** | | | | | | | | |
| Jorge-Ripper 2016 | 9.9 | 35.65 | 47 | 3.8 | 15.38 | 35 | 26.4% | 0.21 [-0.23, 0.65] |
| Lei 2022 | 23.97 | 27.8 | 17 | 23.57 | 24.48 | 35 | 25.2% | 0.02 [-0.56, 0.59] |
| **Subtotal (95% CI)** | | | **64** | | | **70** | | **0.14 [-0.21, 0.49]** |
| **Total (95% CI)** | | | **147** | | | **168** | **100.0%** | **0.97 [0.05, 1.88]** |

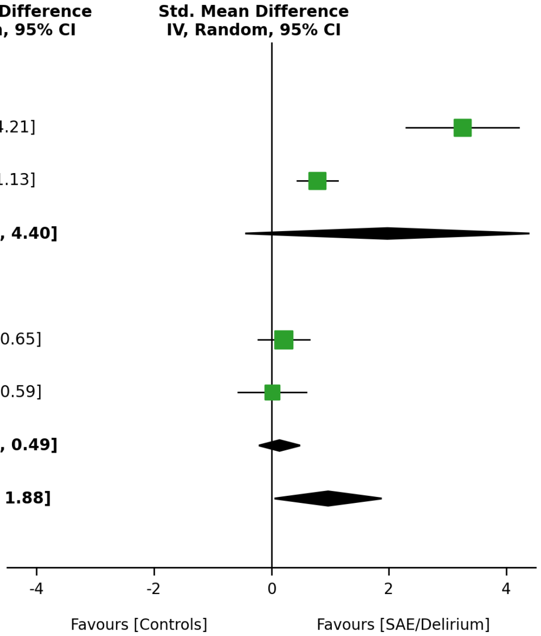


Heterogeneity: Tau² = 0.77; Chi² = 37.21, df = 3; I² = 92%

Test for overall effect: Z = 2.08 (P = 0.038)

Test for subgroup differences: Chi² = 2.17, df = 1 (P = 0.14)

#### eFigure 2E. Forest plot versus septic controls without encephalopathy — NSE

| | SAE/Delirium | | | Control (within sepsis) | | | | |
|---|---|---|---|---|---|---|---|---|
| **Study or Subgroup** | **Mean** | **SD** | **Total** | **Mean** | **SD** | **Total** | **Weight** | **Std. Mean Difference IV, Random, 95% CI** |
| **1.5.1 SAE** | | | | | | | | |
| Feng 2017 | 20.93 | 12.98 | 36 | 15.4 | 10.69 | 23 | 21.6% | 0.45 [-0.08, 0.98] |
| Guo 2022 | 10.16 | 2.11 | 30 | 8.62 | 1.62 | 90 | 22.0% | 0.87 [0.44, 1.30] |
| Li 2022 | 18.4 | 2.2 | 21 | 6.3 | 1.1 | 20 | 15.1% | 6.77 [5.11, 8.43] |
| Wang 2022 | 13.33 | 5.36 | 20 | 10.23 | 2.98 | 20 | 21.1% | 0.70 [0.06, 1.34] |
| **Subtotal (95% CI)** | | | **107** | | | **153** | | **1.86 [0.54, 3.19]** |
| **1.5.2 Delirium only** | | | | | | | | |
| Erikson 2018 | 21.4 | 10.96 | 10 | 22.2 | 15.19 | 12 | 20.1% | -0.06 [-0.90, 0.78] |
| **Subtotal (95% CI)** | | | **10** | | | **12** | | **-0.06 [-0.90, 0.78]** |
| **Total (95% CI)** | | | **117** | | | **165** | **100.0%** | **1.45 [0.33, 2.57]** |

Heterogeneity: $Tau^2 = 1.43$; $Chi^2 = 55.67$, df = 4; $I^2 = 93\%$

Test for overall effect: Z = 2.55 (P = 0.011)

Test for subgroup differences: $Chi^2 = 5.76$, df = 1 (P = 0.02)

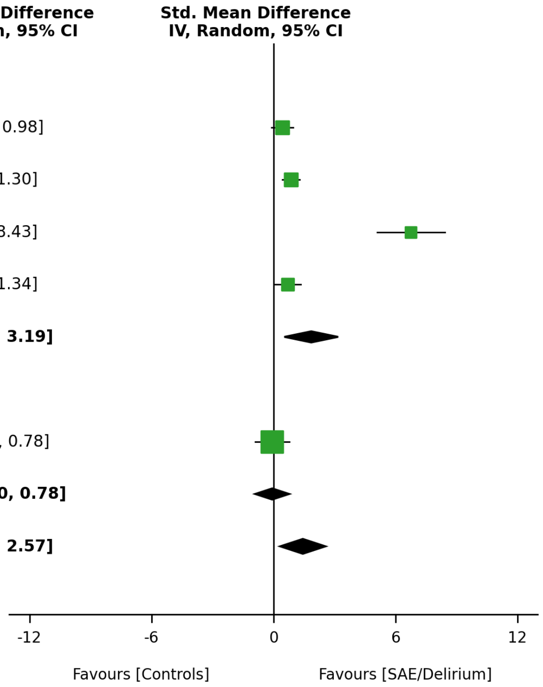

#### eFigure 2F. Forest plot versus septic controls without encephalopathy — IFN-γ

| | SAE/Delirium | | | Control (within sepsis) | | | | | |
|---|---|---|---|---|---|---|---|---|---|
| **Study or Subgroup** | **Mean** | **SD** | **Total** | **Mean** | **SD** | **Total** | **Weight** | **Std. Mean Difference IV, Random, 95% CI** | **Std. Mean Difference IV, Random, 95% CI** |
| **1.6.1 SAE** | | | | | | | | | |
| Yu 2025 | 106.97 | 127.65 | 62 | 46.91 | 49.9 | 78 | 62.5% | 0.64 [0.30, 0.99] | |
| **Subtotal (95% CI)** | | | **62** | | | **78** | | **0.64 [0.30, 0.99]** | |
| **1.6.2 Delirium only** | | | | | | | | | |
| Jorge-Ripper 2016 | 22.8 | 43.19 | 47 | 9.1 | 10.65 | 35 | 37.5% | 0.41 [-0.04, 0.85] | |
| **Subtotal (95% CI)** | | | **47** | | | **35** | | **0.41 [-0.04, 0.85]** | |
| **Total (95% CI)** | | | **109** | | | **113** | **100.0%** | **0.55 [0.28, 0.83]** | |

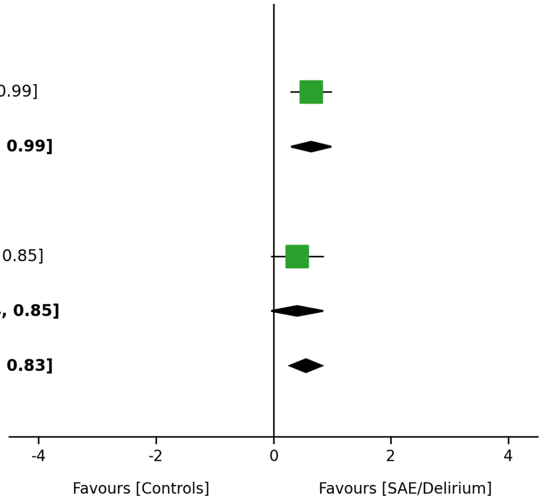


Heterogeneity: Tau² = 0.00; Chi² = 0.70, df = 1; I² = 0%

Test for overall effect: Z = 4.02 (P < 0.001)

Test for subgroup differences: Chi² = 0.70, df = 1 (P = 0.40)

#### eFigure 2G. Forest plot versus septic controls without encephalopathy — CRP

| | SAE/Delirium | | | Control (within sepsis) | | | | |
|---|---|---|---|---|---|---|---|---|
| **Study or Subgroup** | **Mean** | **SD** | **Total** | **Mean** | **SD** | **Total** | **Weight** | **Std. Mean Difference IV, Random, 95% CI** |
| **1.7.1 SAE** | | | | | | | | |
| Pfister 2008 | 318 | 162.96 | 12 | 130 | 148.15 | 4 | 46.2% | 1.11 [-0.10, 2.33] |
| **Subtotal (95% CI)** | | | **12** | | | **4** | | **1.11 [-0.10, 2.33]** |
| **1.7.2 Delirium only** | | | | | | | | |
| Erikson 2018 | 185.33 | 88.15 | 10 | 236 | 105.19 | 12 | 53.8% | -0.50 [-1.35, 0.36] |
| **Subtotal (95% CI)** | | | **10** | | | **12** | | **-0.50 [-1.35, 0.36]** |
| **Total (95% CI)** | | | **22** | | | **16** | **100.0%** | **0.25 [-1.33, 1.82]** |

**Std. Mean Difference IV, Random, 95% CI**

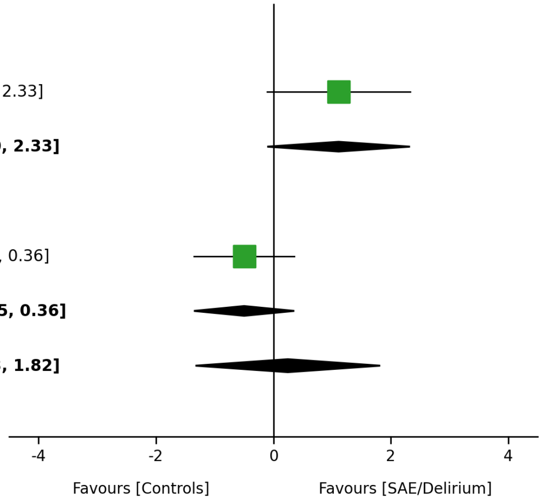


Heterogeneity: $Tau^2 = 1.01$; $Chi^2 = 4.51$, df = 1; $I^2 = 78\%$

Test for overall effect: Z = 0.31 (P = 0.759)

Test for subgroup differences: $Chi^2 = 4.51$, df = 1 (P = 0.03)

#### eFigure 3. Forest plots of the pooled odds ratios for association with the neurological outcome

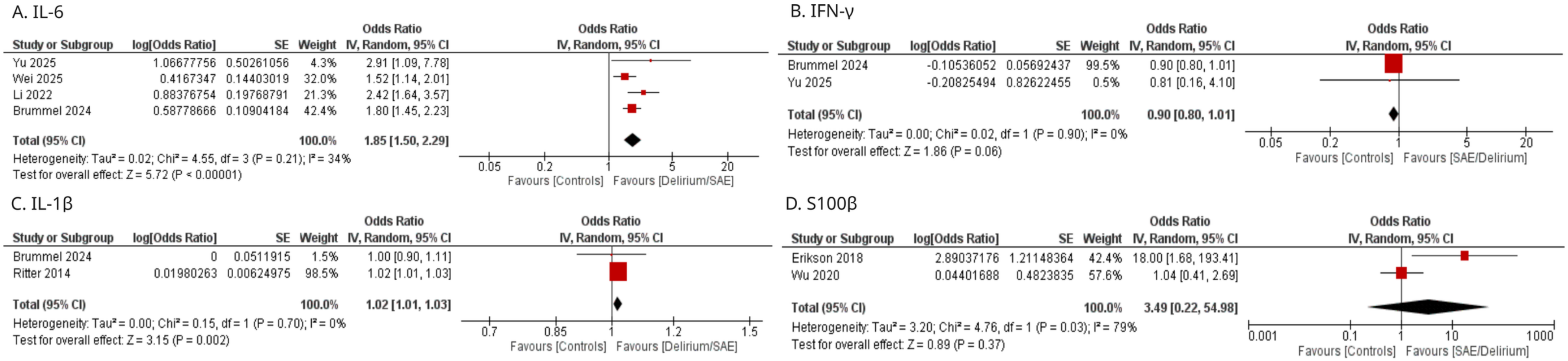


Forest plots of the pooled odds ratios for the association between biomarker concentration and the neurological outcome. The reproduction of each pooled value from the plotted rows is in eTable 19.

**eFigure 4. Pooled effect by whether the study states an ascertainment instrument**

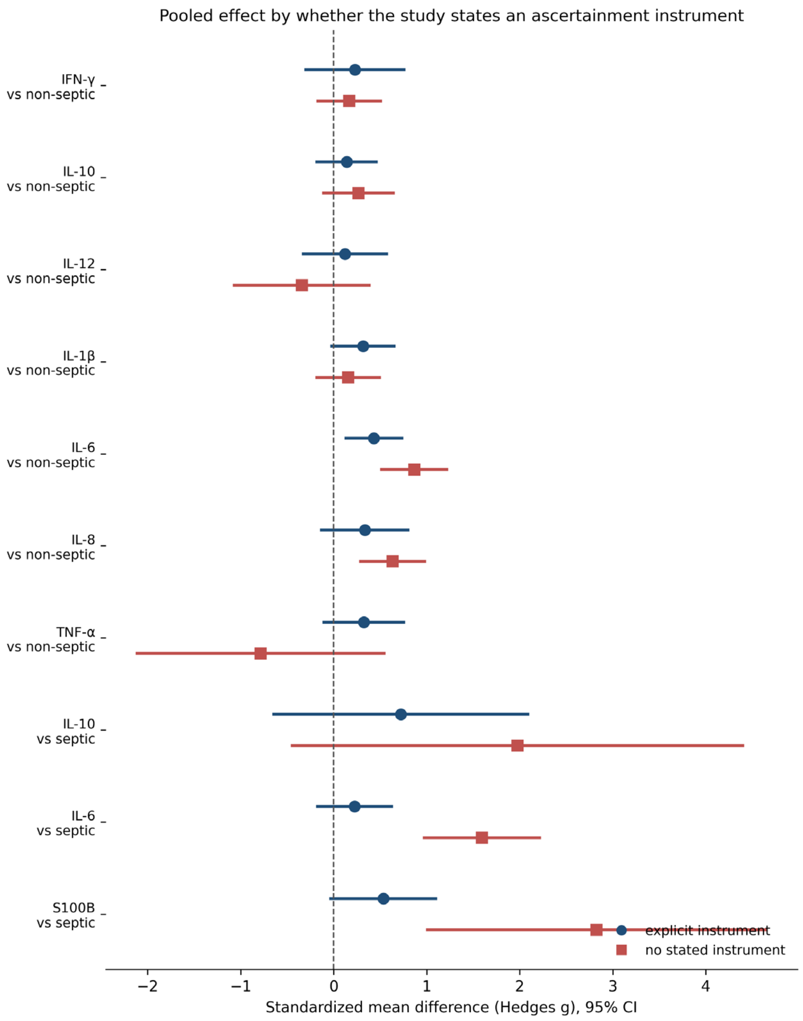


Post hoc subgroup analysis by whether the study states an ascertainment instrument for the neurological case. Pooled standardized mean difference with 95% CI for studies with an explicit instrument and for studies stating none, by biomarker and framework, for every comparison in which both subgroups contain at least two studies. Interaction tests are in eTable 12. This is a descriptive contrast of reporting quality, not a test of a biological hypothesis.
