## Supplementary material for "The Th17/IL-17 Axis and Systemic Inflammatory Biomarkers in Sepsis-Associated Encephalopathy and Delirium: A Systematic Review and Meta-Analysis": Table 1

**Table 1. Characteristics of the 18 Included Studies**

| **Study (Reference)** | **Country** | **Design** | **Participants, n** | **Septic Patients, n** | **SAE/Delirium Arm, n** | **NOS Score** | **Risk of Bias** | **Contributed to a Pooled Comparison** |
| --- | --- | --- | --- | --- | --- | --- | --- | --- |
| Nakamura et al, 2003 (50) | Japan | Prospective interventional | 46 | 26 | 16 | 5/9 | Moderate | Yes |
| Pfister et al, 2008 (51) | Switzerland | Prospective cohort | 16 | 16 | 12 | 8/9 | Low | Yes |
| Ritter et al, 2014 (52) | Brazil | Prospective cohort | 78 | 39 | 31 | 8/9 | Low | Yes |
| Jorge-Ripper et al, 2016 (53) | Spain | Observational cohort | 119 | 83 | 83 | 8/9 | Low | Yes |
| Feng et al, 2017 (54) | China | Retrospective cohort | 59 | 59 | 36 | 4/9 | High | Yes |
| Kuswardhani et al, 2017 (55) | Indonesia | Observational study | 60 | 44 | 60 | - | Not assessed | No |
| Erikson et al, 2018 (56) | Finland | Prospective cohort | 22 | 22 | 10 | 8/9 | Low | Yes |
| Orhun et al, 2019 (57) | Turkey | Prospective cohort | 119 | 86 | 86 | 5/9 | Moderate | Yes |
| Wu et al, 2020 (58) | China | Prospective cohort | 104 | 104 | 59 | 7/9 | Low | Yes |
| Guo et al, 2022 (59) | China | Observational cohort | 120 | 120 | 30 | 7/9 | Low | Yes |
| Lei et al, 2022 (60) | China | Prospective longitudinal | 52 | 28 | 17 | 8/9 | Low | Yes |
| Li et al, 2022 (61) | China | Retrospective cohort | 41 | 41 | 21 | 7/9 | Low | Yes |
| Smith et al, 2022 (62, 68) | United States | Prospective cohort | 119 | 66 | 41 | 8/9 | Low | Yes |
| Wang et al, 2022 (63) | China | Prospective cohort | 40 | 40 | 20 | 7/9 | Low | Yes |
| Mao et al, 2023 (64)ᵃ | China | Case-control | 43 | 43 | 19 | 3/9 | High | Yes |
| Brummel et al, 2024 (65) | United States | Prospective cohort | 991 | 687 | 691 | 8/9 | Low | Yes |
| Wei et al, 2025 (66) | China | Retrospective cohort | 219 | 219 | 105 | 7/9 | Low | Yes |
| Yu et al, 2025 (67) | China | Prospective cohort | 140 | 140 | 78 | 8/9 | Low | Yes |
| **Total** |  |  | 2,388 | 1,863 | 1,415 |  |  | 17 |

NOS = Newcastle-Ottawa Scale (maximum 9 points; low risk ≥ 7, moderate 5–6, high ≤ 4), SAE = sepsis-associated encephalopathy. The SAE/delirium arm column is an arm total and not a prevalence denominator: in several studies the encephalopathy arm was sampled separately from the septic comparison arm, so arm totals do not partition the septic column. One study (Kuswardhani et al, 2017) was included in the review, but no risk-of-bias assessment was supplied and it contributed to no pooled comparison. Studies are labeled by the year used in the analysis file; for Guo et al the indexed publication year is 2021 (reference 59). Smith et al (2022) is reported in two publications (references 62 and 68). ᵃ Biomarkers measured in cerebrospinal fluid; all other studies measured serum or plasma.
