## Supplementary material for "The Th17/IL-17 Axis and Systemic Inflammatory Biomarkers in Sepsis-Associated Encephalopathy and Delirium: A Systematic Review and Meta-Analysis": Table 2

**Table 2. Evidence Map: Volume of Evidence and Pooled Estimates by Biomarker**

| **Biomarker** | **Immunological Domain** | **Pooled Comparisons, k** | **Participants, n** | **SMD vs Non-Septic Controls (95% CI)** | **SMD vs Septic Controls (95% CI)** | **OR for Association With SAE/Delirium (95% CI)** |
| --- | --- | --- | --- | --- | --- | --- |
| IL-6 | Th17-related inflammation | 19 | 2,393 | 0.54 (0.25 to 0.82) | 1.05 (0.51 to 1.59) | 1.85 (1.50 to 2.29) |
| IL-17 | Th17 axis | 3 | 277 | 0.37 (0.01 to 0.73) | Single study, not pooled | Not assessed |
| TNF-α | Innate inflammation | 10 | 1,550 | -0.02 (-0.56 to 0.52) | 0.58 (0.08 to 1.08) | Not assessed |
| IL-1β | Th17-related inflammation | 5 | 1,346 | 0.24 (0.03 to 0.45) | Not pooled | 1.02 (1.01 to 1.03) |
| IL-8 | Innate inflammation | 4 | 1,268 | 0.47 (0.12 to 0.82) | Not pooled | Not assessed |
| IL-10 | Regulatory | 11 | 1,787 | 0.16 (-0.09 to 0.41) | 0.97 (0.05 to 1.88) | Not assessed |
| IFN-γ | Th1 | 6 | 1,445 | 0.15 (-0.09 to 0.39) | 0.55 (0.28 to 0.83) | 0.90 (0.80 to 1.01) |
| S100B | Astroglial injury/BBB dysfunction | 8 | 484 | Not pooled | 1.39 (0.54 to 2.25) | 3.49 (0.22 to 54.98) |
| NSE | Neuronal injury | 5 | 282 | Not pooled | 1.45 (0.33 to 2.57) | Not assessed |
| CRP | Acute phase/innate inflammation | 2 | 38 | Single study, not pooled | 0.25 (-1.33 to 1.82) | Not assessed |
| sTNFR-1 | TNF pathway | 2 | 1,069 | 0.37 (-0.15 to 0.89) | Not pooled | Not assessed |
| IL-12 | Th1/Th17-related | 4 | 1,268 | -0.08 (-0.46 to 0.30) | Not pooled | Not assessed |

BBB = blood-brain barrier, CRP = C-reactive protein, IFN = interferon, IL = interleukin, NSE = neuron-specific enolase, OR = odds ratio, SAE = sepsis-associated encephalopathy, SMD = standardized mean difference, sTNFR-1 = soluble tumor necrosis factor receptor-1, TNF = tumor necrosis factor. Non-septic controls comprise healthy controls, non-septic hospitalized patients and hospitalized patients without delirium. Pooled comparisons and participants are summed across the two contrast frameworks; a study entering both frameworks is counted in each. SMDs are random-effects (DerSimonian-Laird) estimates; a positive SMD indicates a higher concentration in the SAE/delirium group. ORs are pooled from the association framework and are not on the SMD scale. Per-study arm values are in eTable 16 and sensitivity analyses in eTables 25 and 26.
